# Immunogenicity and safety of a recombinant adenovirus type-5 COVID-19 vaccine in adults: data from a randomised, double-blind, placebo-controlled, single-dose, phase 3 trial in Russia

**DOI:** 10.1101/2022.03.01.22271507

**Authors:** Dmitry Lioznov, Irina Amosova, Savely A. Sheetikov, Ksenia V. Zornikova, Yana Serdyuk, Grigory A. Efimov, Mikhail Tsyferov, Mikhail Khmelevskii, Andrei Afanasiev, Nadezhda Khomyakova, Dmitry Zubkov, Anton Tikhonov, Tao Zhu, Luis Barreto, Vitalina Dzutseva

## Abstract

**Background:** To determine the immunogenicity, efficacy, reactogenicity, and safety of a single dose of recombinant adenovirus type-5 vectored COVID-19 vaccine (Ad5-nCoV, 5 × 10^10^ viral particles per 0.5 mL dose), we conducted a single-dose, randomised, double-blind, placebo-controlled, parallel group (3:1 Ad5-nCoV:placebo), phase 3 trial (Prometheus).

**Methods:** From 11-September-2020 to 05-May-2021, across six sites in the Russian Federation, 496 participants were injected with either placebo or Ad5-nCoV expressing the full-length spike (S) protein from the severe acute respiratory syndrome coronavirus 2 (SARS-CoV-2).

**Results:** Seroconversion (the primary endpoint) rates of 78.5% (95% CI: 73.9; 82.6) against receptor binding domain (RBD), 90.6% (95% CI: 87.2; 93.4) against S protein and 59% (95% CI: 53.3; 64.6) against neutralising SARS-CoV2 antibodies 28 days post-vaccination. Geometric mean titres (GMTs) were also elevated for antibodies against the RBD (405.32 [95% CI: 361.58; 454.46]) and S protein (678.86 [95% CI: 607.44; 754.40]) compared to the GMT of neutralising antibodies against SARS-CoV-2 (16.73 [95% CI: 15.36; 18.22]). Using an IFN-γ ELISpot assay after stimulating the cells with full-length S protein we showed that the Ad5-nCoV vaccine induced the most robust cellular immune response on Days 14 and 28. Up to Day 28, the primary and all secondary endpoints of the Ad5-nCoV vaccine were statistically superior to the placebo (р <0.001). Systemic reactions were reported in 113 of 496 (22.8%) participants (Ad5-nCoV, 26.9%; Placebo, 10.5%), and local reactions were reported in 108 (21.8%) participants (Ad5-nCoV, 28.5%; Placebo, 1.6%). These were generally mild and resolved within 7 days after vaccination. Of the six serious adverse events reported, none of the events were vaccine related. There were no deaths or premature withdrawals.

**Conclusion:** A single-dose of Ad5-nCoV vaccine induced a marked specific humoral and cellular immune response with a favourable safety profile.

**Trial Registration:** ClinicalTrials.gov: NCT04540419

## INTRODUCTION

The ongoing COVID-19 pandemic caused by severe acute respiratory syndrome coronavirus 2 (SARS-CoV-2) has resulted in morbidity and mortality unseen since the Spanish flu outbreak more than a century ago [1]. Quarantine measures, a traditional public health method of infection control, have been only partially effective in the face of this highly transmissible virus [2]. Vaccination has the potential to reduce disease severity and transmission but requires expedited development and global administration. In response to this enormous task, many potential vaccine candidates are in development, with several now approved for full or limited use being actively administered [3–6]. A range of technologies have been used in the development of SARS-CoV-2 vaccines, including replicating or non-replicating viral vectors, inactivated viruses and mRNA, DNA or autologous cell-based vaccines. It is not currently known which approach provides the most effective immunity for different recipient risk groups with an acceptable safety profile, all at acceptable levels of cost with ease of manufacturing and distribution [7].

A candidate COVID-19 vaccine that initially showed a capacity to induce a significant antibody and cellular immune response is the adenovirus type 5 (Ad5)-nCoV vaccine developed by CanSino Biologics Inc., Tianjin, and the Beijing Institute of Biotechnology, People’s Republic of China. It consists of a replication-defective Ad5 vector expressing the SARS-CoV-2 spike (S) protein, including its receptor-binding domain (RBD). This protein serves as the main antigen in SARS-CoV-2 vaccines [8]. The Ad5-nCoV vaccine was one of the first to enter phase 1 and 2 clinical trials in China, 2 months after the identification of the virus genotype and the results showed the vaccine to be safe and immunogenic after a single dose [9, 10]. Its potential advantages are single-dose immunization and its proven technology, both of which were used previously for the Ebola vaccine (Ad5-EBOV), and its stability that permits it to be stored in a standard refrigerator at 2–8°C, enabling the ease of worldwide distribution.

A collaborative development project between CanSino Biologics and the Russian pharmaceutical company NPO Petrovax Pharm LLC provided the basis for a new phase 3 study, Prometheus Rus. This multicentre, randomised, double-blind, placebo-controlled clinical trial examines the immunogenicity, reactogenicity, efficacy and safety of the Ad5-nCoV COVID-19 vaccine compared with placebo, in a mostly white, Russian population. The study population was recruited at centres in the western part of the Russian federation (Moscow, St. Petersburg and Yaroslavl) and therefore provides the first clinical data available for Ad5-nCoV in a white European population; previously published data were based on the phase 1 and 2 clinical trials conducted in China. We present the final analysis of the 496 participants in this phase 3 trial.

## MATERIALS AND METHODS

The protocol for this trial and supporting CONSORT checklist are available as supporting information; see S1 Protocol and S1 Checklist, respectively.

### Ethical Conduct of the Study

The study proceeded in accordance with the principles of the Declaration of Helsinki and Good Clinical Practice. The trial protocol was reviewed and approved by the Independent Ethics Committees of the involved sites and the Ethics Council of the Ministry of Health of the Russian Federation. The study design and methodology have been developed in line with the FDA [11], EMA [12] and EAEU [13] guidelines, as well as the regulatory documents of the Russian Federation [14]. The trial is registered with ClinicalTrials.gov, NCT04540419 [15].

To be included, participants needed to be able to understand the content of the informed consent documents and be willing to sign the informed consent form. Written informed consent was obtained from each participant before eligibility screening.

### Study Design and Participants

Prometheus is a multicentre, randomised, double-blind, placebo-controlled, clinical trial being conducted in six centres in the Russian Federation. The study seeks to evaluate the immunogenicity, efficacy, reactogenicity and safety of a single dose of the Ad5-nCoV COVID-19 vaccine compared with placebo in adults up to 6 months after vaccination. Competitive recruitment of the planned sample size of 500 eligible participants in six centres was completed in November 2020 and follow-up observations were completed on 05-May-2021. A planned interim analysis of data collected in 200 participants up to 28 days after a single injection of Ad5-nCoV or placebo was completed on 21-December-2020. Herein, we present the results of final analysis of data collected in 496 participants up to 6 months after a single injection of Ad5-nCoV or placebo.

Participation was sought through online recruitment advertising and patient databases of the trial sites. All participants underwent detailed screening 1–10 days before vaccination with Ad5-nCoV or placebo (Day 0). Screening included the detection of SARS-CoV-2 RNA using real-time polymerase chain reaction (PCR) via a swab, and SARS-CoV-2 immunoglobulin M (IgM) and immunoglobulin G (IgG) antibody testing to ensure negative results, as well as testing for human immunodeficiency virus (HIV), syphilis, hepatitis B and hepatitis C viruses via blood serum. A detailed medical history for each participant was taken and records included if the participant experienced any COVID-19 symptoms and if the participant was in close contact with people suspected or proven to have SARS-CoV-2 infection. Participants underwent physical examination (including a neurological examination, vital signs, and body temperature), haematological, biochemical and coagulation testing, urinalysis, electrocardiogram, and when applicable, pregnancy testing as pregnancy (and breastfeeding) was an exclusion criterion.

Men and women aged 18–85 years with a body mass index (BMI) between 18.5 and 30.0 kg/m^2^ were selected to participate if they had no indication of a current or previous SARS-CoV-2 infection (e.g., respiratory infection in last 14 days, axillary temperature ≥37.0 °C) or close contact with a suspected or confirmed case of SARS-CoV-2 infection. Participants were considered to be eligible if they were in general good health as established by medical history and screening. Those with a range of chronic illnesses, including mental disorders, a history of allergies, recipients of concurrent medication, those with addictions, and those for whom there were concerns over adherence to study protocol were also excluded.

### Randomisation and Masking

The investigational vaccine, Ad5-nCoV, and the placebo were provided by NPO Petrovax Pharm LLC (Moscow, Russia). Both vaccine and placebo were developed by CanSino Biologics Inc. (Tianjin, China) and the Beijing Institute of Biotechnology (Beijing, China). The vaccine was administered with the optimal dose of 5 × 10^10^ viral particles per 0.5 mL dose, as determined in a previous study [9, 10]; placebo contained vaccine excipients only. The appearance of Ad5-nCoV and placebo syringes and packaging was identical.

Eligible participants were randomly allocated to the Ad5-nCoV group or the Placebo group, in a 3:1 ratio, by an independent statistician using a validated system including a pseudorandom number generator with a seed value; allocation used block randomisation and stratification by study site. Neither the investigators nor participants were aware of the group assignment. Investigators were trained to use the centralised interactive web response system that was used for randomisation.

Randomisation codes were kept by authorised personnel from the responsible contracted organisation.

### Procedures

A single dose (0.5 mL) of Ad5-nCoV or placebo was administered by intramuscular (IM) injection to the upper arm on Day 0. Participants were requested to remain at the site for 2 h after vaccination for study staff to monitor for any systemic or local reactions to vaccination. Following the administration of the vaccine or placebo, participants attended clinic visits on Day 2, Day 7, Day 14, Day 28 and after Month 6. Between Day 28 and Month 6 there were phone calls at Months 2, 3, 4 and 5.

Determination of serum antibodies against the S protein and RBD of SARS-CoV-2 and the presence of neutralising antibodies against SARS-CoV-2 were conducted on Day 0, Day 14, Day 28 and after Month 6; neutralising antibodies against Ad5 were assessed on Day 0, Day 28 and after Month 6. Immunoglobulin G antibodies to the S protein and RBD were determined using indirect enzyme-linked immunosorbent assay kits that involved incubation of serially diluted serum samples with the recombinant antigen (either RBD [SARS-CoV-2-IgG-EIA; XEMA Co. Ltd] or S protein [SARS-CoV-2-IgG-EIA-BEST; Vector-Best]) immobilised to the surface of a 96-well plate. The assays were performed according to the manufacturer’s instructions. Horseradish-peroxidase conjugated mouse monoclonal anti-human IgG antibody was used to detect antibodies, visualised with tetramethylbenzidine substrate solution. The detection limit for antibodies against the S protein and RBD of SARS-CoV-2 was 1:100.

Anti-coronavirus neutralising antibodies were determined with a microneutralisation assay in which Vero cell (#ССL-81, American Type Culture Collection) monolayers were incubated in 96-well plates with 2-fold serial dilutions (1:10 to 1:1280) of participant serum. Recently thawed and diluted SARS-CoV-2 virus ([GISAID HCoV-19/st_petersburg-3524S/2020] was obtained from the clinical material collection at Smorodintsev Research Institute of Influenza, St. Petersburg, Russia) was added to the wells, and plates were incubated for 1 h at 37±0.5°C in a humidified incubator. Medium was removed from wells, replaced with a mixture of the most highly diluted (1:1280) serum and virus, and plates were incubated at 37±0.5°C and 5% CO_2_ for 4 days. Anti-adenovirus neutralising antibodies were also determined using microneutralisation assay; A549 cell monolayers were incubated in 96-well plates with serial dilutions (1:10 to 1:1280) of participant serum and working dilutions of Ad5 (Adenovir; Smorodintsev Research Institute of Influenza). Plates were incubated for 2 h at 36±0.5°C and 5% CO_2_. Medium was removed from wells, replaced with a mixture of diluted serums and working virus dilution, and plates were incubated at 36±0.5°C and 5% CO_2_ for 72 h. For both microneutralisation assays, results were assessed by visual inspection of cytopathic effects. Serum titres were determined as the maximum dilution at which complete inhibition of viral reproduction was detected as the result of interaction between virus and specific antibodies; the detection limit was 1:10. Antibody titres undetectable in serum were assigned values of half of the detection limits for calculation.

Sequence encoding ΔFurin variant of SARS-CoV-2 S protein (amino acids 1–1213) containing a C-terminal Gly-Gly-6xHis tag was subcloned into the pMCAG-2T vector using the GeneArt Type IIs Assembly Kit, *Bbs*I (Thermo Fisher Scientific), according to the manufacturer’s instructions. Recombinant SARS-CoV-2 S protein was expressed in Expi293F cells (Thermo Fisher Scientific) as previously described [16]. Five days following transfection, cells were harvested via centrifugation (15,600 x *g*), the supernatant was concentrated and diafiltered using the ÄKTATM flux tangential flow filtration system (Cytiva) into buffer A (10 mM phosphate buffer, 2.7 mM KCl, 500 mM NaCl, pH 8.0). The His-tagged S protein was further purified using Ni-NTA agarose resin (Qiagen), washed with buffer A containing 30 mM imidazole and eluted in buffer A with 200 mM imidazole. Using a Slide-A-Lyzer Dialysis Cassette (20K MWCO, Thermo Fisher Scientific), the eluate was dialysed against PBS (10 mM phosphate buffer, 2.7 mM KCl, 137 mM NaCl, pH 7.5) before use in subsequent experiments.

To measure the cellular immune response from T cells (specifically CD8^+^ and CD4^+^ T cells), peripheral blood mononuclear cells (PBMCs) from participants were isolated and tested as described by Shomuradova et al. [16]. In brief, 30 mL of venous blood was collected from the participants and centrifuged via a density gradient (Ficoll; PanEco) for 400 x *g* for 30 min to isolate PBMCs, which were washed with PBS containing 2 mM EDTA. For the enzyme-linked immunospot (ELISpot) assay, PBMCs (3 x 10^5^ cell/well) were plated into 96-well nitrocellulose plate that was pre-coated with human IFN-γ capture antibody (ImmunoSpot kit Human IFN γ Single-Color ELISpot kit, Cellular Technology Limited) in serum-free test medium (Cellular Technology Limited) containing 1 mM GlutaMAX (Gibco) in a final volume of 200 µL/well as previously described [16]. To stimulate the cells, cells were pulsed in duplicates with the S protein at a final concentration of 10 µg/mL or with a pool of overlapping peptides covering the human S protein (PepTivator SARS-CoV-2 Prot S [Cat. No. 130-126-701; Miltenyi Biotec]) at a final concentration of 1 µM. Plates were incubated at 37°C in 5% CO_2_ for 18 h, and then the assays were performed according to manufacturer’s instructions. Plates were washed twice with PBS, washed twice with PBS + 0.05% Tween-20, and then incubated at room temperature with biotinylated anti-human IFN-γ detection antibody for 2 h.

Plates were then washed three times with PBS + 0.05% Tween-20, and then incubated at room temperature with streptavidin-AP for 30 min. After at least two washes, the colorimetric reaction was initiated by adding the substrate components for 15 min at room temperature. The reaction was halted by gently rinsing the plate with distilled water. Spots were counted with the ImmunoSpot Analyzer using the ImmunoSpot software (Cellular Technology Limited). Samples were designated as positive for a T cell response when the mean number of spots in two replicas minus the number of spots in the negative control was ≥10.

Evidence of local and general reactogenicity (frequency and nature of systemic and local immunisation reactions on the day of vaccination and within 7 days after vaccination) was sought on Day 0, Day 2 and Day 7. Adverse events (AEs) were monitored from the day of vaccination (Day 0) onwards, during a scheduled phone call that evening, and at all subsequent clinic visits. A physical examination, including neurologic examination, and vital signs, including body temperature, was conducted at all visits (screening, Day 0, Day 2, Day 7, Day 14, Day 28 and Month 6). Participants received telephone calls after 2, 3, 4 and 5 months, and were asked to answer questions to assess safety and determine if they presented with signs of an acute respiratory infection due to COVID-19. Haematology, clinical biochemistry, coagulation testing and urinalysis were conducted at screening, day 2 and Day 28; electrocardiography was performed at screening and on day 2. Immunoglobulin E (IgE) levels were assessed to determine any allergic effects of vaccine components at screening and on Day 28.

### Outcomes

The primary endpoint was the seroconversion rate, specifically the percentage of individuals with a 4-fold or higher increase in antibody titres to the RBD of the SARS-CoV-2 S protein, 28 days after vaccination. Secondary endpoints included the assessment of four outcomes: 1) examining the seroconversion rates in response to RBD, S protein and neutralising SARS-CoV-3 antibodies on Day 14, Day 28 (except for RBD) and at Month 6; 2) geometric mean titres (GMTs) of serum antibodies against the RBD, S protein and neutralising antibodies against SARS-CoV-2 on Day 14, Day 28 and Month 6 post-vaccination; 3) GMTs of neutralising antibodies against the Ad5 vector on Day 28 and Month 6 post-vaccination; and 4) the cellular immune response as indicated by the secretion of IFN-γ on Day 14, Day 28 and Month 6 post-vaccination.

Exploratory endpoints were the frequency of laboratory-confirmed COVID-19 cases, severe COVID-19 cases, hospitalisations due to COVID-19 and COVID-19-related deaths (Day 14 to Month 6 post-vaccination). The safety endpoints were reactogenicity (Day 0 to day 7), the frequency and nature of AEs (Day 0 to the end of the study [Month 6 Visit]), and the results of physical and laboratory examinations (i.e., haematological tests, urinalysis, serum IgE concentration).

Of note, two definitions were used to define seroconversion, one quantitative and one qualitative. The primary and secondary endpoints used the quantitative definition: the proportion of participants with at least a 4-fold increase in antibody titres against SARS-CoV-2 S protein and/or its RBD, specifically. The qualitative analysis was defined as an antibody titre above the lower limit of quantification (LLOQ) post-vaccination (if the baseline titre was below the LLOQ), or a 4-fold increase over baseline post-vaccination (if the baseline titre was above the LLOQ).

### Statistical Methods

Statistical analysis was performed using SPSS Statistics, Version 26.0 (IBM Corp.). Initially, as part of the interim analysis and to ensure 90% power for the between-group comparison of the primary variable, 180 subjects were to be included. This was based on the assumptions of a 20% seroconversion rate in the Placebo group (based on quantitative analysis), superiority of the vaccine group of ≥30% (conservative assumption, odds ratio = 4), corrected two-sided significance level of 0.02616 (one-sided level of 0.01308) and the randomisation ratio of 3:1. Considering potential dropouts from the study during the initial observation period of 28 days (10% of subjects), the total number of randomised participants was increased to 200. To provide more detailed safety and efficacy data (including age subgroups) and to descriptively present the frequency of confirmed COVID-19 cases that occurred within 6 months post-vaccination (except for COVID-19 cases that developed during the first 14 days after the vaccination), a sample size of 500 was selected.

An unblinded interim analysis was originally planned and conducted in this clinical trial when obtaining partial information to evaluate the primary endpoint, gathered from the first 200 randomised volunteers and based on the results collected up to Day 28 (Visit 5), including the data for those who left the trial before Visit 5. Subsequently, an alpha spending function (that is, an increasing function of the proportion of the maximum sample size) of the Pocock type was employed to adjust the level of significance due to the multiple comparisons of the primary variable in the planned interim and final analysis sets. Analyses were performed with bilateral alpha levels of 0.02616 and 0.03039 on the interim and final analysis sets, respectively (with a total bilateral significance level of 5%).

Variables representing the seroconversion rate (the proportion of participants with at least a 4-fold increase in antibody titres) were tabulated by evaluation time-points and treatment groups, and two-sided Clopper-Pearson 95% confidence intervals (CIs) presented. Comparative analysis of the primary endpoint (seroconversion, quantitative definition) was performed using the Cochran-Mantel-Haenszel test and the chi-squared test (or Fisher’s exact test).

Antibody GMTs and neutralising antibody GMTs were presented and compared by evaluation time points based on the calculated 95% CIs using two-way analysis of variance (ANOVA) following logarithmic transformation. Mean log-transformed differences between the study groups (vaccine and placebo) were evaluated with the corresponding 95% CI. The point estimates of the mean differences and the corresponding CIs were back-transformed.

Geometric mean fold-increases in antibody and neutralising antibody titres for each treatment group and each antibody GMT evaluation time point were assessed with the corresponding two-sided 95% CIs following log-transformation, point estimation of the difference and CIs, and back-transformation of obtained values. Between-group comparisons were performed using ANOVA following the logarithmic transformation. Results from the ELISpot assay to assess differences in cellular immunity were analysed using Mann-Whitney test with Bonferroni adjustment of p-values.

Statistical significance was accepted at p<0.05. Results for immunogenicity analyses are presented for the full analysis set ([FAS], which included all eligible participants who received a dose of vaccine and provided at least one immunogenicity assessment result). Results for the per protocol set ([PPS], which included members of the FAS who received a dose of vaccine according to the randomisation and study scheme and provided data for immunogenicity assessment before and after vaccination in line with the study scheme, did not receive a prohibited therapy, had no significant protocol deviations that could have impacted efficacy assessment and did not develop COVID-19 within the first 14 days post-vaccination) were similar to those of the FAS and are therefore not presented. Cellular immunity results were analysed in participants from the FAS that provided at least one cellular immunity assessment result. Prophylactic efficacy results are presented for the PPS (for efficacy analysis). The safety analysis set included all randomized volunteers who received a dose of the vaccine.

### Patient and Public Involvement

Participants were not directly involved in the development, implementation, or interpretation of this study due to the requirement for a quick response to the rapidly-evolving coronavirus pandemic.

### Data Sharing

The Authors commit to making the relevant anonymised participant level data available upon reasonable request for 3 years following publication of this study. Requests should be directed to the corresponding author.

## RESULTS

Out of the 783 participants who were screened, this analysis included 500 eligible participants intended for vaccination at six locations in the Russian Federation between 11 September 2020 and 11 November 2020. Of these, 374 participants were randomised to the Ad5-nCoV group and 126 to the Placebo group (Fig. 1). The safety analysis set included 496 participants, the FAS for immunogenicity analysis included 495 participants, and the PPS for efficacy analysis included 481 participants.

**Fig. 1.**
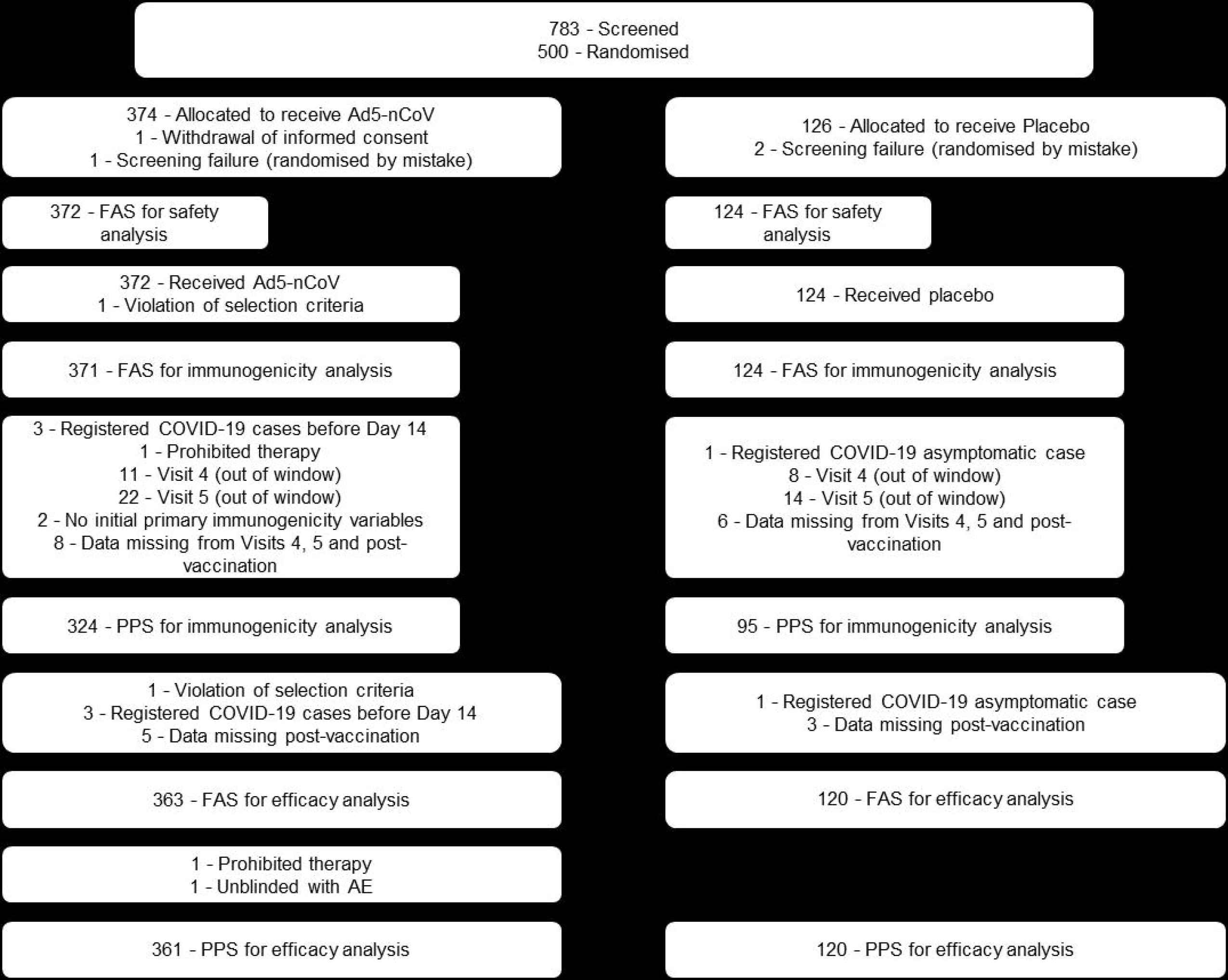
Participant flow diagram. AE, adverse event; FAS, Full Analysis Set; PPS, Per-Protocol Set.

The mean age of participants was 41.2 years (range 18–79 years), with 300 (60.5%) participants aged 18–44 years, 161 (32.5%) aged 45–59 years, and 35 (7.1%) aged 60 years or older (Table 1). There were more males (297 out of 496 [59.9%]) than females. The vast majority of the participants (99.4%) were white race and 3 participants were Asian. Baseline characteristics were largely similar across groups.

**Table 1.** Participant Demographics.

|  | **Ad5-nCoV**<br>**N=372** | **Placebo**<br>**N=124** | **Total**<br>**N=496** |
| --- | --- | --- | --- |
| **Age (years[%])** |  |  |  |
| 18–44 years | 223 (59.9) | 77 (62.1) | 300 (60.5) |
| 45–59 years | 122 (32.8) | 39 (31.5) | 161 (32.5) |
| ≥60 years | 27 (7.3) | 8 (6.5) | 35 (7.1) |
| Mean | 41.2 | 41.0 | 41.2 |
| **Sex (n[%])** |  |  |  |
| Male | 151 (40.6) | 48 (38.7) | 199 (40.1) |
| Female | 221 (59.4) | 76 (61.3) | 297 (59.9) |
| **Race (n[%])** |  |  |  |
| White | 371 (99.7) | 122 (98.4) | 493 (99.4) |
| Asian | 1 (0.3) | 2 (1.6) | 3 (0.6) |
| **Country (n[%])** |  |  |  |
| Russia | 372 (100%) | 124 (100%) | 496 (100%) |
| **Body Mass Index (kg/m<sup>2</sup>)** |  |  |  |
| Mean | 25.03 | 24.7 | 24.94 |
| Minimum | 18.6 | 18.6 | 18.5 |
| Maximum | 30.0 | 29.9 | 30.0 |
| **Underlying Disease (n[%])** |  |  |  |
| Yes | 34 (22.8) | 9 (18.0) | 43 (21.6) |
| No | 115 (77.2) | 41 (82.0) | 156 (78.4) |
| **Prior Disease (n[%])** |  |  |  |
| Yes | 176 (47.3) | 56 (45.2) | 232 (46.8) |
| No | 93 (62.4) | 36 (72.0) | 129 (64.8) |

### Immunogenicity and efficacy results

Baseline antibody titres of the participants in the FAS (Ad5-nCoV; placebo) were similar for anti-RBD antibodies (50.1; 50.0), anti-S protein (50.3; 50.3), neutralising anti-SARS-CoV-2 antibodies (5.0; 5.0), as well as neutralising antibodies against the Ad5 vector (11.3; 8.8). Administration of the Ad5-nCoV vaccine led to a marked increase in anti-RBD antibody response compared with placebo across the FAS, with GMTs of 138.61 (95% CI: 125.60; 152.97) on Day 14 and 405.32 (95% CI: 361.58; 454.46) on Day 28; differences compared with placebo achieved statistical significance on days 14 and 28 (both p<0.001; Fig. 2). Six months following vaccination, antibodies levels in the vaccinated group remained high with a mean GMT of 153.07 (95% CI: 133.08; 176.08). Interestingly, a notable increase in anti-RBD antibodies at Month 6 was also observed in the Placebo group, which could be attributed to an asymptomatic infection (GMT of 115.50 [95% CI: 88.84; 150.18], p=0.064).

**Fig. 2.**
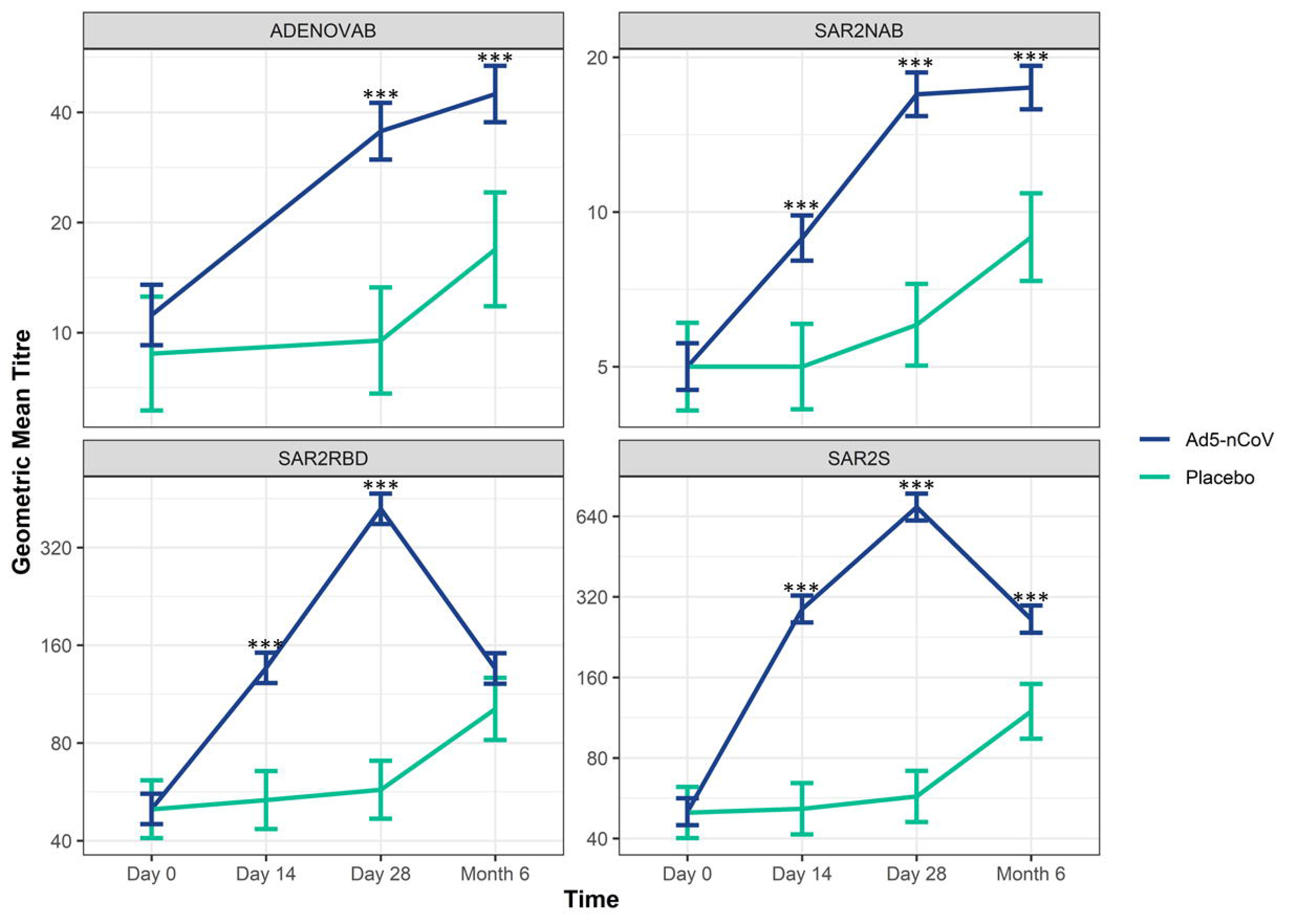
Geometric mean titre (GMT) of serum antibodies against the RBD and SARS-CoV-2 S protein on Day 0, Day 14, Day 28 and Month 6 after vaccination. The GMTs with 95% CI are shown for serum antibodies against Ad5 (ADENOVAB), SARS-CoV-2 neutralising antibodies (SAR2NAB), RBD (SAR2RBD) and S protein (SAR2S) (***, p<0.001). Ad5, adenovirus type-5; CI, confidence interval; RBD, receptor binding domain; S, spike; SARS-CoV-2, severe acute respiratory syndrome coronavirus 2.

In the primary analysis on Day 28, 285 (78.5%, 95% CI: 73.9; 82.6) of 363 participants in the Ad5-nCoV group showed seroconversion of RBD-specific antibodies compared with 7 (5.9%; 95% CI: 2.4, 11.7) of 119 participants in the Placebo group, which was indicative of a significant treatment difference (Ad5-nCoV – placebo was 72.6% [95% CI: 65.7, 78.1; p<0.001]). On Day 14, 152/359 (42.3%) participants from the Ad5-nCoV group showed seroconversion of RBD-specific antibodies compared with the Placebo group (p☐001). However, at Month 6, 140/353 (39.7%) participants in the Ad5-nCoV group showed a slight reduction in the seroconversion rate of RBD-specific antibodies compared with the Placebo group (35/101 [34.7%]; p=0.354).

The vaccine dose also induced a significant increase in anti-S protein antibody responses, with a GMT of 288.60 on Day 14 (95% CI: 258.05; 322.78) compared with the Placebo group (51.49 [95% CI: 42.34; 62.62; p☐0.001]); moreover, the significant increase in anti-S protein was also observed on Day 28, with a GMT of 676.87 (95% CI: 607.44; 754.40) compared with the Placebo group (61.31 [95% CI: 50.71; 74.11; p[0.001]). This increasing trend in anti-S protein antibody responses continued even at Month 6, with a GMT of 293.22 (95% CI: 252.12; 341.11) compared with the Placebo group (145.85 [95% CI: 109.85; 193.64; p 0.001]) (Fig. 2). Similarly, 239 (66.6%, 95% CI: 61.4; 71.4) of 359 participants had seroconverted with S protein-specific antibodies by Day 14 compared with 1 (0/8%, 95% CI: 0.0; 4.6) of 118 the Placebo group (p[0.001). Additionally, 329 (90.6%, 95% CI: 87.2; 93.4) of 363 participants also had significantly elevated levels of seroconversion with S protein-specific antibodies by Day 28 compared with the Placebo group (6.7% [95% CI: 2.9; 12.8; p 0.001]). Treatment difference on Day 28 was 65.9% (95% CI; 59.6; 71.3) compared with the Placebo group (p<0.001). At Month 6, 222 (62.9% (95% CI: 57.6; 67.9) of 353 participants had seroconverted with S protein-specific antibodies compared to the Placebo group (41 [40.6%] of 101 participants; p☐001).

Administration of the Ad5-nCoV vaccine induced significant greater neutralising antibody responses to SARS-CoV-2 than placebo (p<0.001), with GMTs of 8.78 on Day 14 (95% CI: 8.19; 9.41) and 16.73 on Day 28 (95% CI: 15.36; 18.22). At Month 6, significantly greater neutralising antibody responses to SARS-CoV-2 were still present, with a GMT of 19.50 (95% CI: 16.97; 22.41) compared with a GMT of 8.94 (95% CI: 6.63; 12.04) in the Placebo group (p[0.001) (Fig. 3). Seroconversion for neutralising antibodies to SARS-CoV-2 occurred in 75 (24.4%, 95% CI: 19.7; 29.5) of 308 participants on Day 14. No increase in neutralising antibody from baseline was observed in the Placebo group. On Day 28, seroconversion for neutralising antibodies to SARS-CoV-2 occurred in 183 (59.0%, 95% CI: 53.3; 64.6) of 320 participants; notably, greater proportions were observed in the Ad5-nCoV group on days 14 and 28 compared with the Placebo group (p<0.001). At Month 6, 149 (48.9%, 95% CI 43.1; 54.6) of 305 participants had a 4-fold or greater increase of neutralising antibodies compared with the Placebo group (32/93 [34.4%, 95% CI: 24.9; 45.0; p=0.012]).

**Fig. 3.**
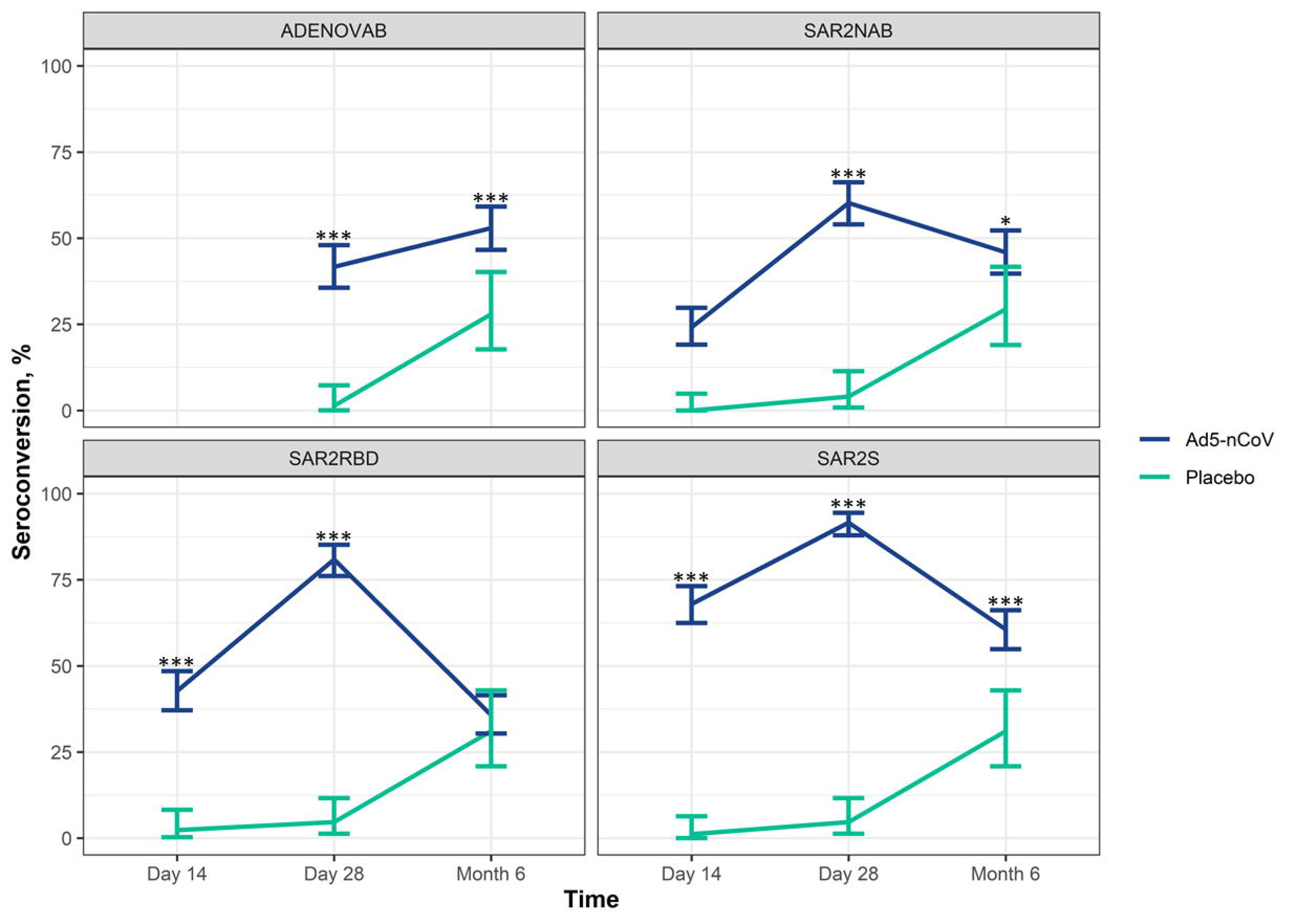
Seroconversion rates of the neutralising antibodies against SARS-CoV-2 and the seroconversion rate against the RBD and SARS-CoV-2 S protein on Day 0, Day 14, Day 28 and Month 6 after vaccination. The seroconversion rates with 95% CI are shown for serum antibodies against Ad5 (ADENOVAB), SARS-CoV-2 neutralising antibodies (SAR2NAB), RBD (SAR2RBD) and S protein (SAR2S) (***, p<0.001; *, p<0.05). Ad5, adenovirus type-5; CI, confidence interval; RBD, receptor binding domain; S, spike; SARS-CoV-2, severe acute respiratory syndrome coronavirus 2.

Almost all participants had neutralising antibodies against the Ad5 vector at baseline: 317/372 (85.2%) participants who received Ad5-nCoV and 106/124 (85.5%) who received placebo. For most participants, the antibody titre to Ad5 was low (≤1:200) (Fig. 4); very few participants had high (>1:200) levels of pre-existing antibodies to Ad5 (7/424, 1.7%). Geometric mean anti-Ad5 antibody titre in participants with low pre-existing anti-Ad5 antibodies increased from 11.3 at baseline (n=317) to 36.5 at Day 28 (n=353), which then increased further to 48.3 (n=355) at Month 6.

**Fig. 4.**
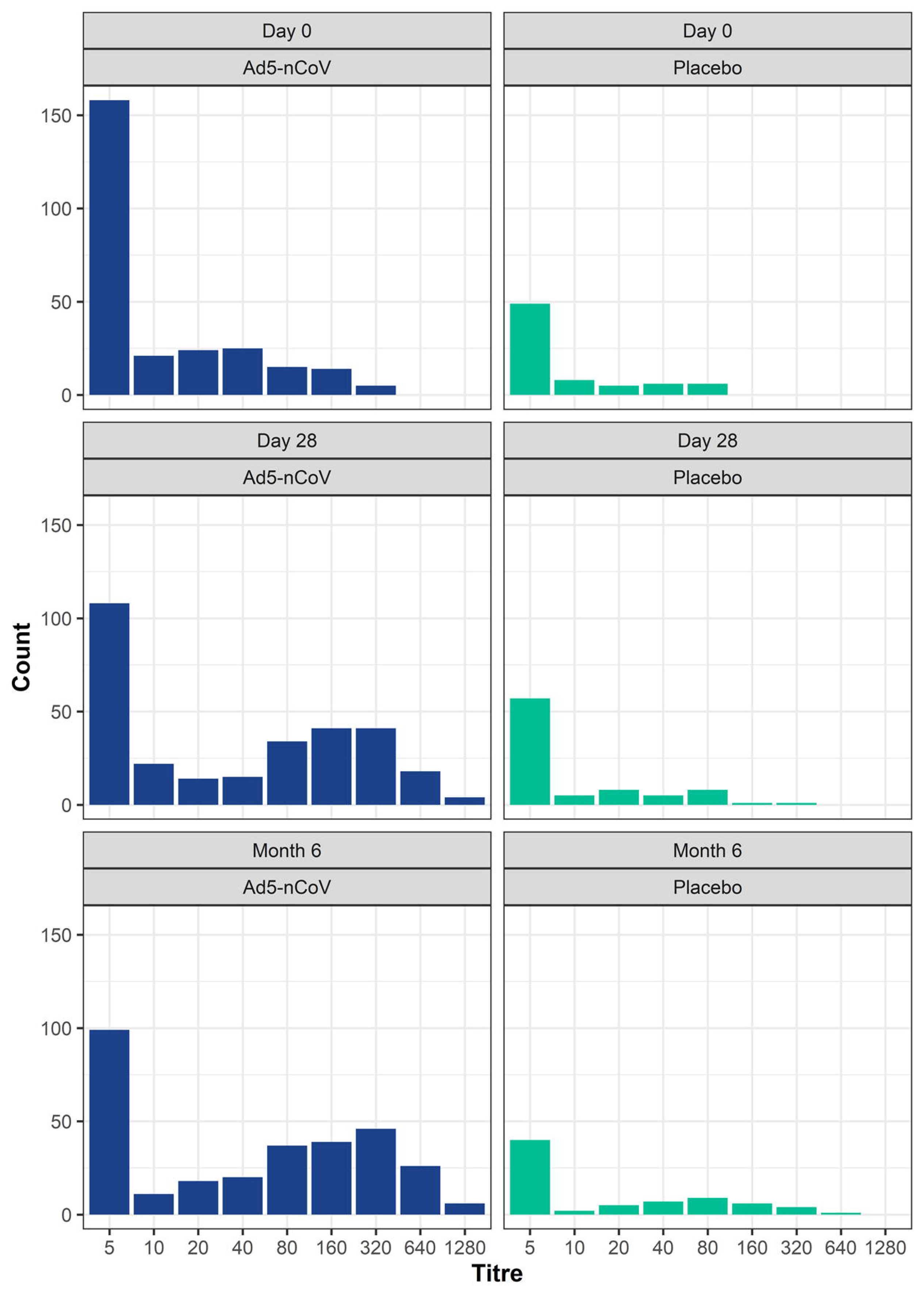
Titre distribution of anti-Ad5 antibodies on Day 0, Day 28 and Month 6 after vaccination in those from either the Ad5-nCoV or Placebo groups.

Corresponding values for the very few participants with pre-existing immunity to Ad5 >1:200 showed an increase from 320.0 at baseline (n=6) to 359.2 at Day 28 (n=6) and a further increase to 557.2 at Month 6 (n=6). The GMT of neutralising antibodies against the Ad5 vector in all participants who received Ad5-nCoV was 11.28 on Day 0 (95% CI: 9.95; 12.78), 36.48 on Day 28 (95% CI: 30.72; 43.30) and 48.34 at Month 6 (95% CI: 40.25; 58.05), with significant levels of neutralising antibodies against Ad5 present at Day 28 and Month 6, compared with the Placebo group (p☐001).

The effect of pre-existing immunity to Ad5 on the immunological responses to the vaccine was studied. As presented in Fig. 5, the increase of GMTs to anti-RBD antibodies after vaccination in the Ad5-nCoV group was comparable between patients with high (>1:200) and low (≤1:200) baseline titres of antibodies to Ad5 up to Day 28, although this observation may be attributed to the small number of patients with pre-existing high Ad5 titres (n=7).

**Fig. 5.**
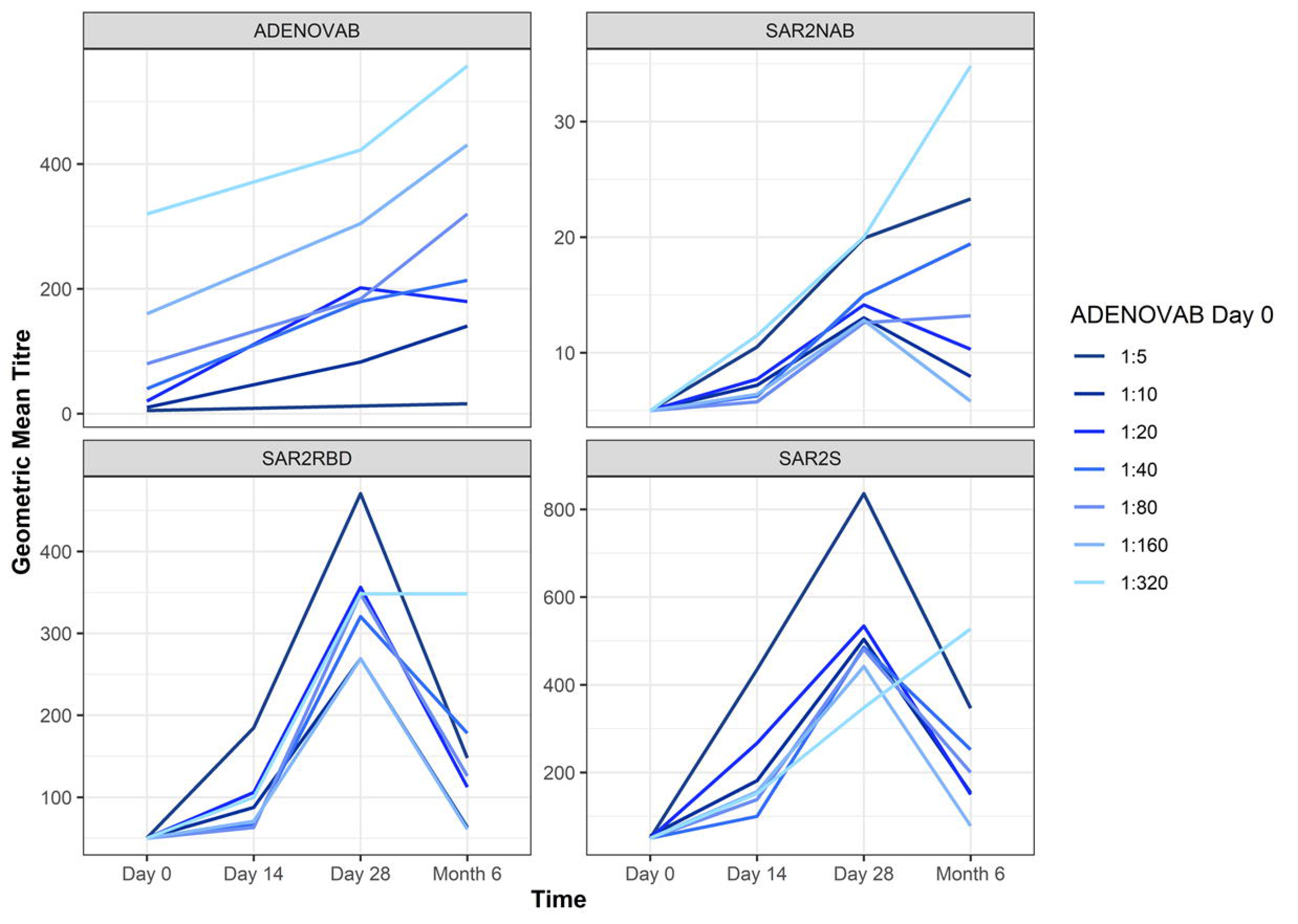
Antibody responses to SARS-CoV-2 vaccine in those with pre-existing immunity to Ad5. Geometric mean titre (GMT) of titres in response to anti-Ad5 antibody (ADENOVAB), anti-RBD (SAR2RBD) and S protein (SAR2S) antibodies as well as neutralising SARS-CoV-2 antibodies (SAR2NAB) on Day 0, Day 14, Day 28 and Month 6 after vaccination. Volunteers are divided into seven (from 1:5 to 1:320) cohorts based on their pre-existing anti-Ad5 antibody levels. Ad5, adenovirus type-5; CI, confidence interval; RBD, receptor binding domain; S, spike; SARS-CoV 2, severe acute respiratory syndrome coronavirus 2.

Pearson’s correlation coefficients between baseline GMTs to Ad5, GMTs to RBD and S protein antibodies as well as neutralising SARS-CoV-2 antibodies were calculated. There was a negative correlation of -0.36 between baseline GMT to Ad5 and GMT to RBD on Day 14 that weakened to -0.05 at 6 months post-vaccination. The correlations between baseline GMT and GMTs to S protein and neutralising SARS-CoV-2 antibodies decreased by Month 6 (Fig. 6). The decrease in correlation was less pronounced between GMTs to RBD, indicating that the relationship between levels of baseline Ad5 GMTs to the COVID-19 humoral immune response weakens over time following vaccination.

**Fig. 6.**
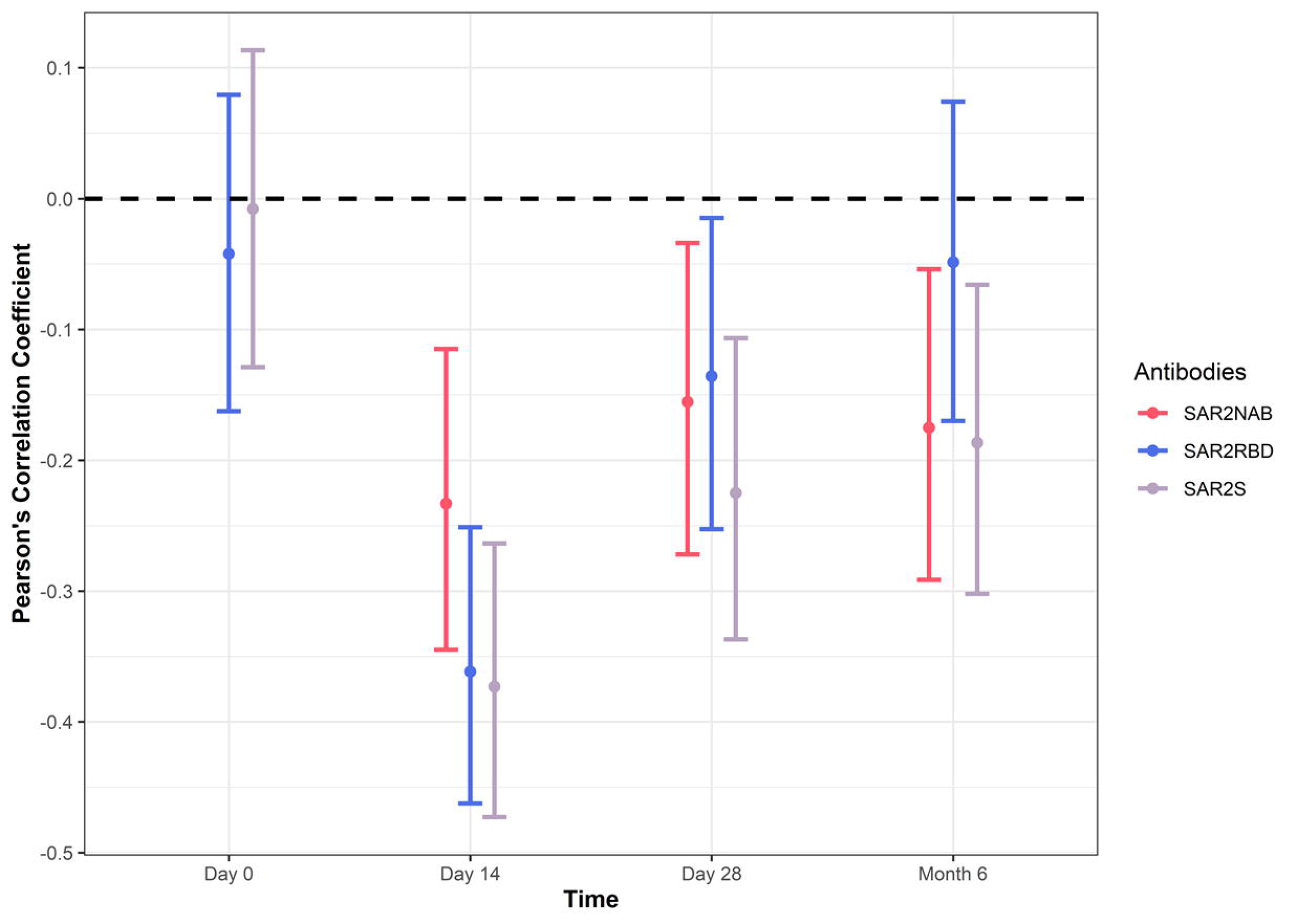
Pearson’s correlation coefficient between GMTs versus anti-Ad5 antibodies (ADENOVAB) in response to anti-RBD (SAR2RBD) and S protein (SAR2S) antibodies as well as neutralising SARS-CoV-2 antibodies (SAR2NAB) on Day 0, Day 14, Day 28 and Month 6 after vaccination. The correlation coefficients with 95% CI are shown. Ad5, adenovirus type-5; CI, confidence interval; RBD, receptor binding domain; S, spike; SARS-CoV 2, severe acute respiratory syndrome coronavirus 2.

The effect of the vaccination on cellular immune responses was studied in a subgroup of 69 participants recruited by Moscow research centres (Ad5-nCoV group, n=50 volunteers; placebo group, n=19). The presence of SARS-CoV-2 specific CD8^+^ and CD4^+^ T cells in venous blood samples was assessed by counting the number of spot-forming cells (SFCs; i.e., IFN-γ secreting cells) in an ELISpot assay in which isolated PBMCs were stimulated with either full-length S protein or a pool of overlapping peptides covering the S protein (i.e., peptide pool). The most pronounced response occurred on Day 14 after the Ad5-nCoV vaccination in which the median number of SFCs was 32.8 (Quartile [Q]1: 19.0, Q3: 78.9) when stimulated with the peptide pool and 32.2 (Q1: 16.0, Q3: 99.9) when stimulated with full-length S protein (Fig. 7A and 7B, respectively). For cells stimulated with the peptide pool, the median number of SFCs decreased to 5.5 (Q1: 1.5, Q3: 12.2) on Day 28 and further still to 1.5 (Q1: 0, Q3: 8.5) at Month 6. When stimulated with full-length S protein, the number of SFCs increased to 4.5 (Q1: 3.0, Q3: 12.5) on Day 28 and 5.5 (Q1: 0.75, Q3: 13.8) by 6 months post-vaccination.

**Fig. 7.**
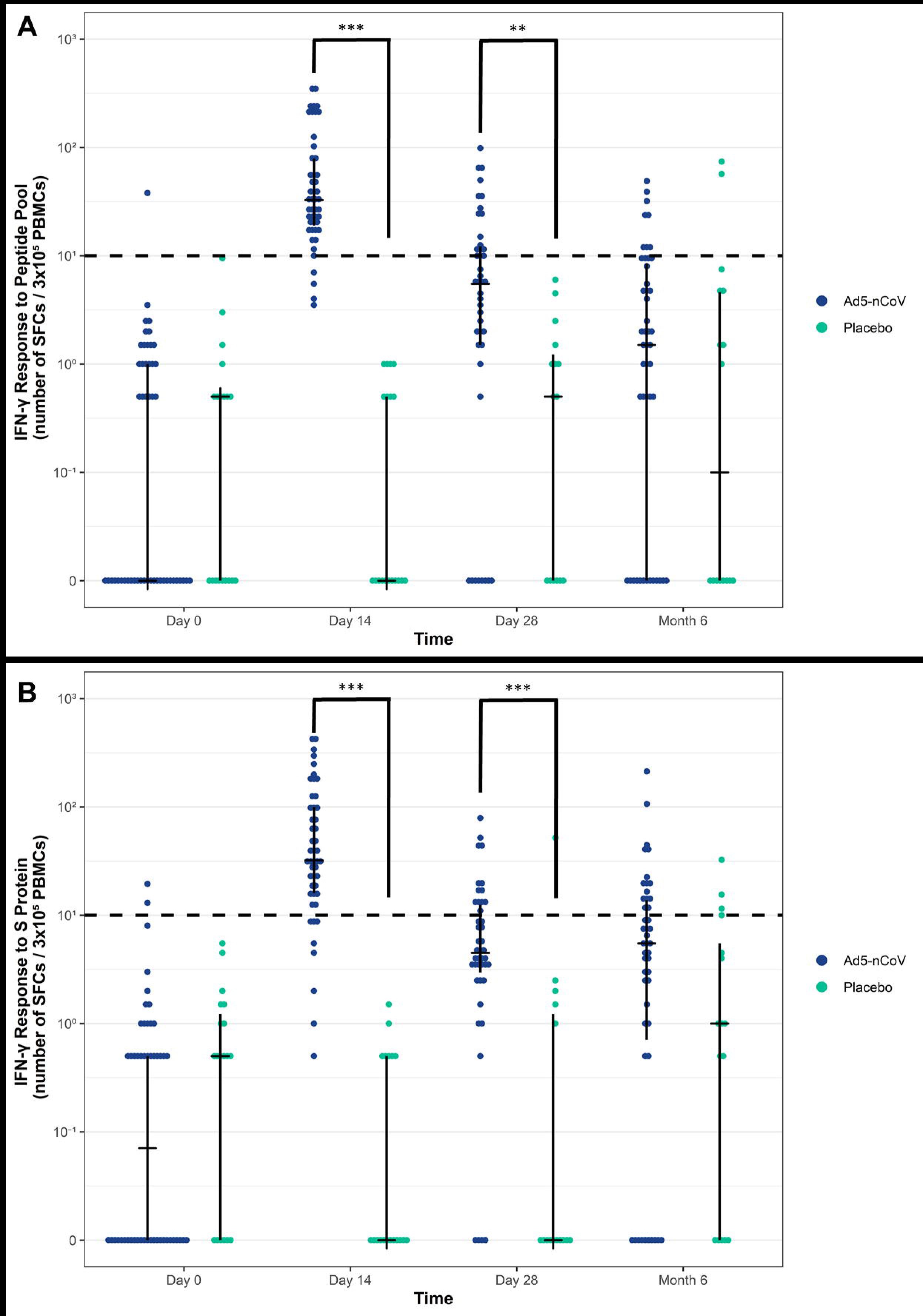
Assessing the cellular immune response to SARS-CoV 2 vaccine using an enzyme-linked immunospot (ELISpot) assay. Peripheral blood mononuclear cells (PBMCs) were isolated from blood samples taken from participants from either the Placebo or Ad5-nCoV group. PBMCs isolated on Day 0, Day 14, Day 28 and Month 6 after vaccination were stimulated with (**A**) a pool of peptides that span the S protein (i.e., peptide pool) or (**B**) the full-length S protein. The response of T cells was determined by counting IFN-γ-positive spot-forming cells (SFCs). Spots were quantified via an automated digital image. The median with quartiles is the plotted, the dotted line indicates the positivity threshold (***, p<0.001; **, p<0.01). Ad5, adenovirus type-5; S, spike; SARS-CoV-2, severe acute respiratory syndrome coronavirus 2.

Compared with the response observed in the Placebo group, those in the Ad5-nCoV group displayed significantly higher numbers of IFN-γ-positive T cells, regardless of how the cells were stimulated, on Days 14 (p<0.001) and 28 (p<0.01). However, 6 months post-vaccination, the differences between the groups were not statistically significant when cells were stimulated with the full-length S protein (p=0.6) or with the peptide pool (p=1.0).

The percentage of participants who had an IFN-γ response above the threshold following stimulation with the peptide pool on Day 14 in the Ad5-nCoV group was 91.7% (95% CI: 80.4; 96.7), whereas no participants were positive in the Placebo group. During the follow-up period, the response rate decreased for those in the Ad5-nCoV group and the percentage of patients that displayed an IFN-γ response on Day 28 was 37.2% (95% CI: 24.4; 52.1), whereas no participants were positive in the Placebo group. At Month 6, the percentage of patients displayed an IFN-γ response was 21.2% and 12.5% in the Ad5-nCoV and Placebo groups, respectively. The trend of an IFN-γ response for cells stimulated with full-length S protein was similar to the trend observed when cells were stimulated with the peptide pool. The percentage of participants who displayed an IFN-γ response following stimulation with full-length S protein in the Ad5-nCoV group on Day 14 was 83.3% (95% CI: 70.4; 91.3), whereas no participants were positive in the Placebo group. During the follow-up period, the response rate markedly decreased for those in the Ad5-nCoV group and the percentage of patients that displayed an IFN-γ response on Day 28 was 30.2 % (95% CI: 18.6; 45.1), whereas there was one participant who had an IFN-γ response in the Placebo group. At Month 6, 31% of patients (95% CI: 20.4; 46.2) displayed an IFN-γ response that was comparable to 25% of patients in the Placebo group following stimulation with the full-length S protein.

Of note, 31 of the participants were confirmed to have COVID-19 within 14 days post-vaccination. Thirteen participants were from the Placebo group (13/31 [11.32%]), whereas 18/31 (4.97%) participants were from the Ad5-nCov group (p=0.023). Cases of severe COVID-19 (with the exception of COVID-19 cases that occurred during the first 14 days post-vaccination) were registered in only 2 (1.7%) participants, both of whom were from the Placebo group. Both of these severe COVID-19 cases required hospitalization. No deaths from COVID-19 were registered.

### Safety evaluation

#### Systemic (general) immunisation reactions

A total of 113 (22.8%) of the 372 participants who received the Ad5-nCoV vaccine reported systemic reactions (Table 2). The incidence of systemic reactions was significantly higher than in the Placebo group, where 13 (10.5%) of the 124 participants reported systemic reactions.

**Table 2.** Summary of adverse events (AEs) by safety analysis set.

|  | **Ad5-nCoV**<br>**N = 372** | **Placebo**<br>**N = 124** | **Total**<br>**N = 496** |
| --- | --- | --- | --- |
| **Systemic (general) immunisation reactions** | **100 (26.9%)/162** | **13 (10.5%)/20** | **113 (22.8%)/182** |
| Grade 1 | 78 (21.0%)/131 | 10 (8.1%)/16 | 88 (17.7%)/147 |
| Grade 2 | 17 (4.6%)/26 | 3 (2.4%)/4 | 20 (4.0%)/30 |
| Grade 3 | 5 (1.3%) | 0 | 5 (1.0%) |
| **Local immunisation reactions** | **106 (28.5%)/180** | **2 (1.6%)/4** | **108 (21.8%)/184** |
| Grade 1 | 91 (24.5%)/154 | 2 (1.6%)/4 | 93 (18.8%)/158 |
| Grade 2 | 15 (4.0%)/26 | 0 | 15 (3.0%)/26 |
| Grade 3 | 0 | 0 | 0 |
| **AEs (except for immunisation reactions)** | **152 (40.9%)/365** | **38 (30.6%)/75** | **190 (38.3%)/440** |
| **AEs related to vaccination** | **130 (34.9%)/280** | **25 (20.2%)/43** | **155 (31.3%)/323** |
| Grade 1 | 106 (28.5%)/246 | 20 (16.1%)/36 | 126 (25.4%)/282 |
| Grade 2 | 21 (5.6%)/30 | 5 (4.0%)/7 | 26 (5.2%)/37 |
| Grade 3 | 3 (0.8%)/4 | 0 | 3 (0.6%)/4 |
| **AEs related to vaccination and registered during the first 7 days after vaccination** | **123 (33.1%)/244** | **25 (20.2%)/42** | **148 (29.8%)/286** |
| Serious AEs | 1 (0.3%)/1 | 5 (4.0%)/5 | 6(1.2%)/6 |
| Death | 0 | 0 | 0 |
| Early discontinuation due to AE | 0 | 0 | 0 |

Data presented as the number of participants with adverse events per category (%) / number of AE records.

The most commonly reported reactions in the Ad5-nCoV group included increase in body temperature (20.2%), headache (5.9%), fatigue (5.4%), myalgia (4.8%) and arthralgia (1.9%; S1 Table). For most participants, general immunisation reactions were mild (Grade 1, 21.0%). Moderate (Grade 2) reactions occurred in 4.6% of participants and included increase in body temperature, headache, myalgia, arthralgia, and fatigue. Severe (Grade 3) reactions, including an increase in body temperature (39.0–40.0 °C), occurred in 4 participants, and myalgia occurred in only 1 participant (Table 2 and S2 Table).

In the Placebo group, reactions included increase in body temperature (6.5%), headache (4.8%), fatigue (5.4%) and diarrhoea (0.8%). These AEs were mild or moderate; no severe reactions were reported (Table 2, S1 Table and S2 Table).

There were no fatal outcomes. During the first 7 days after vaccination, a total of six serious AEs (SAEs) were reported (Table 2): one SAE in 1 (0.3%) participant from the Ad5-nCoV group and five SAEs in 5 (4.0%) participants from the Placebo group. Of note, two of the participants from the Placebo group were hospitalised due to COVID-19. For the only SAE event experienced by a participant from the Ad5-nCoV group, the event was found to have no connection with the study drug. The SAEs are described in S3 Table.

#### Injection site reactions

Injection site reactions occurred in 106 participants (28.5%) who received the Ad5-nCoV vaccine. By comparison, only 2 participants in the Placebo group (1.6%) experienced an injection site reaction including swelling and pain or induration (S4 Table), all of which were Grade 1 in severity (S5 Table). The most common reactions included pain (16.9%), erythema (14.8%) and induration at the injection site (4.8%) (S4 Table). As listed in S5 Table, for most participants, local injection site reactions were mild (24.5%). Fifteen participants (4.0%) who received the Ad5-nCoV vaccine had a moderate reaction that included pain, erythema, swelling or induration at the injection site. No severe reactions were reported.

#### Other adverse events (excluding immunisation reactions)

A total of 190 participants (38.3%) reported other AEs during the 6 months after vaccination; 152/365 (40.9%) participants from the Ad5-nCoV group and 38/75 (3.6%) participants from the Placebo group. As listed in Table 2, these AEs were assessed by the investigators as related to the vaccine (Ad5-nCoV: 34.9%; Placebo: 20.2%). The majority of vaccine-related AEs were reported during the first 7 days after vaccination (Ad5-nCoV: 33.1%; Placebo: 20.2%). In both groups, the most common AEs were out-of-range laboratory measurements: 34.4% reported by participants in the Ad5-nCoV group and 16.9% from the Placebo group. The most pronounced differences in AE incidence in the Ad5-nCoV group compared with the Placebo group were: increases in C-reactive protein (17.6% vs. 2.4%), monocytes (4.0% vs. 0%) and aspartate aminotransferase (1.6% vs. 1.6%) as well as a decrease in neutrophils (6.2% vs 2.4%) (S6 Table).

For most participants, AEs were mild, reported in 28.5% of the Ad5-nCoV group and 16.1% of the Placebo group. Moderate events were registered in 5.6% of participants who received the Ad5-nCoV vaccine and 4.0% who received the placebo. By comparison, SAEs occurred in 0.8% of participants in the Ad5-nCoV group, but none in the Placebo group; these included increases in C-reactive protein (0.5%) and blood fibrinogen levels (0.5%).

For most participants, general and injection site reactions and AEs resolved within 7 days after vaccination.

Results of the analysis of laboratory parameters demonstrated a trend towards an increase in C-reactive protein levels, an increase in the red cell sedimentation rate, an increase in the mean percentage of monocytes, and a decrease in the mean percentage of neutrophils after administration of Ad5-nCoV. The changes in the examined laboratory parameters were pronounced the day after vaccination, but had largely resolved by Day 28.

The proportion of participants in the Ad5-nCoV group with elevated IgE at Day 28 showed no changes from screening: 73/372 (19.6%) to 76/365 (20.8%).

## DISCUSSION

In this multicentre, randomised, double-blind, placebo-controlled, phase 3 trial including 500 adult participants aged 18–85 years (mean age: 41.2 years), the immunogenicity, efficacy and safety of the Ad5-nCoV COVID-19 vaccine was assessed up to 6 months after vaccination. Both study groups (Ad5-nCoV and placebo) had similar baseline characteristics. A single immunisation with the Ad5-nCoV vaccine led to a marked immune response. The primary endpoint (seroconversion rate of anti-RBD antibodies on Day 28 after vaccination) and all secondary endpoints on Day 28 after vaccination showed statistically significant superiority of the Ad5-nCoV vaccine compared with placebo (p<0.001).

The seroconversion rate of antibodies against the RBD of the SARS-CoV-2 S protein (78.5%) and of neutralising antibodies against SARS-CoV-2 (59%) on Day 28 were similar to the results obtained during the interim analysis of a phase 2 clinical study conducted in China. The seroconversion rate of anti-RBD antibodies in the Ad5-nCoV group was 97% (95% CI: 92; 99), and the seroconversion rate of neutralising antibodies against live SARS-CoV-2 was 47% (95% CI: 39; 56) [9].

The GMTs of anti-RBD, S protein-specific antibodies and neutralizing SARS-CoV-2 antibodies increased significantly at Month 6 post-vaccination in the Placebo group, not by Day 14 or Day 28. We theorized two possible explanations. First, some individuals might have been asymptomatic while infected with COVID-19, or COVID-19-positive patients did not report their symptoms to the study’s physicians. Second, individuals in the Placebo group might have taken a commercially available antibody test that was widely available in Russia and decided to receive another vaccination after observing their low antibody titres, thereby confounding their treatment status. Two participants were excluded from the PPS for this reason, but many more could have been unreported.

Adenovirus exposure is common. Pre-existing anti-Ad5 immunity may affect the immunogenicity of Ad5-based vaccines, and as a result, their efficacy against COVID-19. However, the proportion of individuals with high anti-Ad5 titres in a population varies across regions [17–19] and it is unclear to what extent previous exposure influences existing titres and the speed of their decline. A titre of 1:200 was selected as the cut-off point for low and high anti-Ad5 antibodies during the phase 1 and 2 studies in China. Those with low baseline anti-Ad5 titres (≤ 200) had RBD-specific antibody and 1: neutralising antibody levels roughly twice as high as those with high baseline anti-Ad5 titres (>1:200) [9, 10]. Our data analysis used a cut-off of 1:200 in line with the Chinese studies, but given the variable levels of anti-Ad5 titres around the world, a limit of 1:200 may not necessarily be appropriate in all regions. The selection of an appropriate cut-off point in the current study was hampered by a lack of published data on anti-Ad5 levels in the Russian population. Among the Russian participants, those with baseline levels of anti-Ad5 antibodies showed a very small group of 7 (1.4%) participants with titres of approximately 1:320 (Fig. 4). All of them were in the subgroup of participants aged 18 to 45 years.

In this study, we uncovered the response of pre-existing immunity to Ad5 and showed how it affects subsequent humoral immune responses as well as the longevity of immunity to COVID-19. In participants vaccinated with Ad5-nCoV, there was no difference in humoral immunity to COVID-19 between those with baseline Ad5 titres above or below the cut-off of 1:200. However using a cut-off of 1:5, we showed that those with baseline Ad5 titres of 1:5 displayed a greater amount of GMTs to anti-RBD and S protein antibodies as well as neutralising SARS-CoV-2 antibodies than those with a higher baseline ratio of Ad5 titres (Fig. 5). The relationship between the levels of baseline Ad5 GMTs to the respective antigen tends to weaken over time following vaccination (Fig. 6). Studies with longer follow-up periods would help to clarify the longevity of the immune response in those vaccinated with Ad5-nCoV. Moreover, studies with a larger dataset would help to determine if pre-existing immunity to Ad5 correlates to protection efficacy.

Our study provides data on the incidence of anti-Ad5 immunity in the Russian population, admittedly in a relatively small sample size. Almost all participants had pre-existing neutralising antibodies to Ad5, although levels were generally very low. No such data were available before this study, and only limited and highly variable data are available for the level of Ad5 immunity in Europe.

The results from this study demonstrate that the experimental Ad5-nCoV vaccine has a good safety profile comparable with the findings of preceding clinical trials in healthy adults. Most post-vaccination AEs were mild or moderate in severity. Although the proportions of participants who reported adverse reactions such as an increase in body temperature, headache, and pain at the injection site were higher in those that received the Ad5-nCoV vaccine, adverse reactions within 28 days were generally mild to moderate and the majority resolved within 7 days after vaccination. All Grade 3 AEs occurred among participants from the Ad5-nCoV group and were similar to commonly reported AEs after other types of immunisation.

There are currently multiple recombinant Ad-vectored COVID-19 candidate vaccines in development [4–7, 9, 10, 20, 21]. The most relevant comparator vaccine in terms of the practicalities of storage, transport and administration is the Ad26.COV2.S vaccine [22]. Like the Ad5-nCoV vaccine, it can be administered as a single dose, stored in a standard refrigerator without the need for ultra-low temperature freezing and is stable at room temperature prior to administration. An interim analysis of Ad26.COV2.S showed that vaccine-induced neutralising antibodies against wild-type virus were detected in 95% or more of participants on day 29 after dosing. Our qualitative analysis used a similar definition of seroconversion to that used to assess the Ad26.COV2.S vaccine [22]; as well as including participants who seroconverted from below the LLOQ of the assay at baseline to detectable antibodies at Day 28, it included those with a 4-fold or greater increase in antibody titre over the same period. The Ad5-nCoV vaccine provided a similar antibody response to Ad26.COV2.S: 95.9% of subjects developed antibodies to the S protein and 92.5% to the smaller anti-RBD region by Day 28.

This study has several limitations. First, the participants included in this study were all white, although conversely, this also provided the first data in a non-Chinese, European population. Second, this trial did not include children or pregnant women, and there were only a small number of older adults (35 were ≥60 years in the Ad5-nCoV group). An ideal candidate vaccine for COVID-19 should cover vulnerable populations of all ages. Anti-S protein-specific antibodies have been reported to decline rapidly in individuals who have recovered from COVID-19, especially those who were asymptomatic or had mild symptoms [23]. Third, the sample size was relatively small and some of the calculated 95% CIs were wide. Finally, virus mutation, an emerging problem, may reduce the effectiveness of current vaccines [24]. It is not known whether participants of this study were exposed to any COVID-19 variants. Further study is underway to determine neutralising antibodies to the widely circulating variants following vaccination with Ad5-nCoV, which include but are not limited to the Alpha (B.1.1.7), Beta (B.1.351) and Gamma (P.1) variants.

### Conclusions

Analysis of data from this phase 3 trial demonstrated the immunogenicity and safety of this Ad5-vector based COVID-19 vaccine. More data are required to determine whether this vaccine reduces infections and transmission. Overall, this stable, single-dose vaccine could contribute to the global fight against the evolving SARS-CoV-2 virus.

## DECLARATIONS

## Supporting information

S1 Checklist

S1 Protocol

## Data Availability

All data produced in the present work are contained in the manuscript and supplmentary materials

## Acknowledgements

Medical writing support was provided by Dr Justin Cook, Dr Antria Siakalli and Dr Caitlin Tolbert of Niche Science and Technology Ltd, Richmond-upon-Thames, Surrey, UK; this was paid for by NPO Petrovax Pharm LLC, 1, Moscow region, Russian Federation.

## Supporting Information

S1 Protocol. Study Protocol.

S1 Checklist. CONSORT Checklist.

S1 Table. Systemic (general) post-vaccination reactions (Safety population).

S2 Table. Systemic (general) post-vaccination reactions by severity (Safety population).

S3 Table. Summary of all severe adverse events that led to hospitalisation or prolonged hospitalisation.

S4 Table. Local post-vaccination reactions (Safety population).

S5 Table. Local post-vaccination reactions by severity (Safety population).

S6 Table. Other adverse events unrelated to vaccination reactions (Safety population).

## Author Contributions

### Conceptualization

Vitalina Dzutseva, Grigory A. Efimov, Mikhail Tsyferov, Anton Tikhonov, Andrei Afanasiev.

### Data Curation

Dmitry Lioznov, Irina Amosova.

### Formal Analysis

Dmitry Zubkov.

### Investigation

Savely A. Sheetikov, Ksenia V. Zornikova, Yana Serdyuk, Grigory A. Efimov, Dmitry Lioznov, Irina Amosova.

### Methodology

Grigory A. Efimov, Dmitry Lioznov, Tao Zhu, Luis Barreto, Vitalina Dzutseva.

### Project Administration

Vitalina Dzutseva, Mikhail Khmelevskii.

### Resources

Savely A. Sheetikov, Ksenia V. Zornikova, Yana Serdyuk, Grigory A. Efimov, Vitalina Dzutseva.

### Supervision

Grigory A. Efimov, Vitalina Dzutseva, Dmitry Lioznov.

### Validation

Irina Amosova, Grigory A. Efimov, Dmitry Lioznov.

### Visualisation

Dmitry Zubkov, Anton Tikhonov.

### Writing – Original Draft Preparation

Anton Tikhonov, Nadezhda Khomyakova, Andrei Afanasiev.

### Writing – Review and Editing

Anton Tikhonov, Nadezhda Khomyakova, Dmitry Lioznov, Grigory A. Efimov, Mikhail Tsyferov, Irina Amosova, Savely A. Sheetikov, Ksenia V. Zornikova, Yana Serdyuk, Dmitry Zubkov, Andrei Afanasiev, Tao Zhu, Luis Barreto.

## Funding and role of study sponsor

The study was designed, funded, and managed by NPO Petrovax Pharm LLC (Moscow, Russian Federation). NPO Petrovax Pharm LLC in partnership with CanSino Biologics, Inc. (Tianjin, China) are funding and managing the clinical development of the Ad5-nCoV vaccine in the Russian Federation.

## Conflicts of interest/competing interests

All authors have read the journal’s policy and the authors of this manuscript have the following competing interests:. MT, MK, DZ, AA, NK, AT and VD are employees of NPO Petrovax Pharm LLC. TZ and LB are employees of CanSino Biologics, Inc. IA, SS, KZ and YS have received funding from NPO Petrovax Pharm LLC for consultation services. DL and GE have received personal fees from NPO Petrovax Pharm LLC for consultation services.

## Availability of data and material

The authors confirm that the data supporting the findings of this study are available within the article.

## Transparency declaration

The corresponding author and guarantor (Vitalina Dzutseva) affirms that this manuscript is an honest, accurate, and transparent account of the study being reported; that no important aspects of the study have been omitted; and that any discrepancies from the study as planned have been explained.

## Notes

### Clinical Trial

NCT04540419

### Funding Statement

The study was funded by NPO Petrovax Pharm LLC
(Moscow, Russian Federation). NPO Petrovax Pharm LLC in partnership with CanSino
Biologics, Inc. (Tianjin, China) are funding and managing the clinical development of
the Ad5-nCoV vaccine in the Russian Federation.

### Author Declarations

Independent Ethics Committees of the involved sites and the Ethics Council of the Ministry of Health of the Russian Federation gave ethical approval for this work.

