## Supplementary material for "Immunogenicity and safety of a recombinant adenovirus type-5 COVID-19 vaccine in adults: data from a randomised, double-blind, placebo-controlled, single-dose, phase 3 trial in Russia": S1 Protocol

### CLINICAL TRIAL PROTOCOL

**Multicentre, randomised, double-blind, placebo-controlled, parallel-group clinical trial to evaluate the efficacy, reactogenicity and safety of recombinant novel coronavirus vaccine (Ad5-nCoV) in adults**

| | |
|---|---|
| Protocol Code: | Prometheus_Rus |
| Investigational Product: | Ad5-nCoV, Recombinant Novel Coronavirus SARS-Cov-2 Vaccine (Adenovirus Type 5 Vector), solution for intramuscular injection |
| Sponsor: | NPO Petrovax Pharm LLC<br>1, Sosnovaya St., Moscow region, Podolsk, Pokrov village 142143, Russia<br>Tel./fax: +7 (495) 926 21 07 |
| Study Phase: | III |
| Protocol Version: | Final version 3.0 |
| Protocol Date: | 01 October 2020 |

**Confidentiality Statement**

*The information herein is strictly confidential and is intended solely as a guide for clinical trials. Any partial or full reproduction or disclosure of the information herein to parties not related to the clinical trial, or its use for any other purpose without the prior written consent of NPO Petrovax Pharm LLC is prohibited.*

Translated from Russian into English of 01/02/2021

### SIGNATURE PAGE 1 (SPONSOR)

By signing this Protocol approval page, the Sponsor represented by its authorised representative agrees to the content of the Protocol, the list of the collected data and the application of relevant common medical standards and clinical trial principles.

#### Sponsor's Representative:

| | |
|---|---|
| Natalya Gordeeva | Director for Drug Development, Registration and Pharmacovigilance<br>NPO Petrovax Pharm LLC |
| Signature: | */signature/* |
| Date: | *26/08/2020* |

#### SIGNATURE PAGE 2

#### (CONTRACT RESEARCH ORGANISATION)

By signing this Protocol approval page, the company conducting the clinical trial on behalf of the Sponsor agrees that the trial will be conducted in accordance with the Protocol and Good Clinical Practice.

| | |
|---|---|
| Dmitry Sharov | CEO<br>OCT Rus Ltd. |
| Signature: | ______________________________ |
| Date: | ______________________________ |

**SIGNATURE PAGE 3**
**(PRINCIPAL INVESTIGATOR)**

By signing this Protocol approval page, the investigator agrees with the content of the Protocol and confirms his/her readiness to conduct the clinical trial at his/her trial site in accordance with all requirements of the Protocol, including confidentiality of data, and in compliance with the Good Clinical Practice and current regulatory requirements of the Russian Federation.

Trial site name and address: ________________________________________

________________________________________

________________________________________

________________________________________

Full name and title of the investigator: ________________________________________

________________________________________

Signature: ________________________________________

Date: ________________________________________

### LIST OF ABBREVIATIONS

| | |
|---|---|
| **AE** | Adverse event |
| **ALP** | Alkaline phosphatase |
| **ALT** | Alanine aminotransferase |
| **aPTT** | Activated partial thromboplastin time |
| **ARVI** | Acute respiratory viral infection |
| **AST** | Aspartate aminotransferase |
| **ATC** | Anatomical Therapeutic Chemical classification system |
| **BMI** | Body mass index |
| **CI** | Confidence interval |
| **CNS** | Central nervous system |
| **COVID-19** | Coronavirus disease 2019 |
| **CRF** | Case Report Form |
| **CRO** | Contract research organisation |
| **DAP** | Diastolic arterial pressure |
| **ECG** | Electrocardiography |
| **EIA** | Enzymoimmunoassay |
| **ELISA** | Enzyme-linked immunosorbent assay |
| **Elispot** | Enzyme-linked immunospot assay |
| **ESR** | Erythrocyte sedimentation rate |
| **GC** | Glucocorticoid |
| **GCP** | Good Clinical Practice |
| **GIT** | Gastrointestinal tract |
| **GMT** | Geometric mean titer |
| **HIV** | Human immunodeficiency virus |
| **HR** | Heart rate |
| **IBD** | Immunobiological drugs |
| **IC** | Informed consent |
| **ICH** | International Council for Harmonisation |
| **IFN** | Interferon |
| **IL** | Interleukin |
| **IWRS** | Interactive Web Response System |
| **LDH** | Lactate dehydrogenase |
| **LEC** | Local ethics committee |
| **MedDRA** | Medical Dictionary for Regulatory Activities |
| **PCR** | Polymerase chain reaction |
| **PT** | Prothrombin time |
| **QPPV** | Qualified person for pharmacovigilance |
| **RBD** | Receptor-binding domain |
| **RNA** | Ribonucleic acid |
| **RR** | Respiratory rate |
| **SAP** | Systolic arterial pressure |
| **SARS** | Severe acute respiratory syndrome |
| **SARS-CoV** | Severe acute respiratory syndrome coronavirus |
| **SD** | Standard deviation |
| **SUADR** | Serious unexpected adverse drug reaction |
| **sADR** | Serious adverse drug reaction |
| **SAE** | Serious adverse event |
| **SOP** | Standard operating procedure |
| **TNF** | Tumour necrosis factor |
| **VP** | Viral particles |
| **WHO** | World Health Organisation |

### 1 STUDY ADMINISTRATIVE STRUCTURE

| | |
|---|---|
| **Sponsor** | NPO Petrovax Pharm LLC<br>1, Sosnovaya St., Moscow region, Podolsk, Pokrov village 142143, Russia<br>Tel./fax: +7 (495) 926 21 07 |
| **Sponsor's responsible person** | Natalya Gordeeva<br>Director for Drug Development, Registration and Pharmacovigilance<br>NPO Petrovax Pharm LLC |
| **Qualified Person for Pharmacovigilance (QPPV)** | Jeffrey Poplavsky<br>NPO Petrovax Pharm LLC<br>12, Presnenskaya embankment, Federation Tower East, floor 38, Moscow 123112, Russia<br>Tel.: +7 (800) 234 44 80<br>Fax: +7 (495) 730 75 60<br> |
| **Monitoring** | Yuri Zaretsky<br>3M Veritas LLC<br>54, Anri Barbusa St., office 004–006, Perm 614107, Russia<br>Tel.: +7 (342) 255 49 02, ext. 750<br> |
| **Contract research organisation (CRO)** | OCT Rus Ltd.<br>5, Kovenskiy per., lit. B, 7$^{th}$ floor, Saint Petersburg 191014, Russia<br>Tel.: +7 (812) 449-86-34<br>Fax: +7 (812) 449-86-35 |
| **Central independent laboratories** | INVITRO Independent Laboratory LLC<br>16, 4$^{th}$ Tverskaya Yamskaya Street, bldg 3, 3$^{rd}$ floor, office I, room 3, Moscow 125047, Russia<br><br>NEXELIS LABORATORIES CANADA INC<br>525 Boulevard Cartier West, Laval, Quebec Canada, H7V 3S8<br><br>Federal State Budgetary Institution National Medical Research Centre for Haematology of the Ministry of Health of the Russian Federation, Laboratory of Transplantation Immunology<br>4, Novy Zykovskiy proyezd, Moscow 125167, Russia<br><br>Federal State Budgetary Institution Smorodintsev Research Institute of Influenza of the Ministry of Health of the Russian Federation, Biotechnology Department, Cell Culture Laboratory<br>15/17, Professora Popova St., Saint Petersburg 197376, Russia |
| **Trial sites** | Information will be provided in a separate document. |

### 2 SYNOPSIS

| | |
|---|---|
| **Study Title** | Multicentre, randomised, double-blind, placebo-controlled, parallel-group clinical trial to evaluate the efficacy, reactogenicity and safety of recombinant novel coronavirus vaccine (Ad5-nCoV) in adults |
| **Protocol Code** | Prometheus_Rus |
| **Protocol Version and Date** | Final version 3.0 of 01 October 2020 |
| **Study Phase** | III |
| **Investigational Product** | Ad5-nCoV, Recombinant Novel Coronavirus SARS-Cov-2 Vaccine (Adenovirus Type 5 Vector), solution for intramuscular injection, $5x10^{10}$ VP per dose (0.5 mL) |
| **Comparator Product** | Placebo |
| **Study Purpose** | Evaluation of efficacy, reactogenicity and safety of the Ad5-nCoV vaccine compared with placebo in adults aged 18 to 85 years. |
| **Study Objectives and Endpoints** | The primary objective of the study is to prove the superiority of the Ad5-nCoV vaccine compared with placebo in terms of seroconversion (proportion of subjects with at least four-times increase in antibody titers against the receptor-binding domain [RBD] of the SARS-CoV-2 S protein) on Day 28 after vaccination.<br>*Secondary Objectives*<br>1. To evaluate the immunogenicity of the Ad5-nCoV vaccine compared with placebo based on the following:<br>• Geometric mean titer of serum antibodies against the RBD and SARS-CoV-2 S protein on Day 14, 28 and 6 months after vaccination.<br>• Seroconversion rate (proportion of subjects with at least four-times increase in antibody titers against the RBD and SARS-CoV-2 S protein) on Day 14, 28 (S protein only) and 6 months after vaccination.<br>• Geometric mean fold-rise in titers of serum antibodies against the RBD and SARS-CoV-2 S protein on Day 14, 28 and 6 months after vaccination.<br>• Geometric mean titer of neutralising antibodies against SARS-CoV-2 on Day 14, 28 and 6 months after vaccination.<br>• Seroconversion rate (proportion of subjects with at least four-times increase in neutralising antibody titers against SARS-CoV-2) on Day 14, 28 and 6 months after vaccination.<br>• Geometric mean fold-rise in titers of neutralising antibodies against SARS-CoV-2 on Day 14, 28 and 6 months after vaccination.<br>• Geometric mean titer of neutralising antibodies against the Ad5 vector on Day 28 and 6 months after vaccination.<br>• Geometric mean fold-rise in titers of neutralising antibodies against the Ad5 vector on Day 28 and 6 months after vaccination.<br>• Cellular immune response (the number of IFNy-secreting T cells [ELISpot]; percentage of CD4+ and CD8+ T cells expressing IFNy, TNF and IL-2 [flow cytometry]) on Day 14, 28 and 6 months after vaccination.<br>2. To evaluate the efficacy of the Ad5-nCoV vaccine (exploratory analysis) based on the following:<br>• Frequency of confirmed COVID-19 cases during 6 months after vaccination (except for the cases occurred during the first 14 days after vaccination). *A confirmed COVID-19 case means the presence of clinical manifestations and a positive laboratory test result for SARS-CoV-2 RNA.*<br>• Frequency of confirmed COVID-19 cases requiring hospitalisation (except for the cases occurred during the first 14 days after vaccination).<br>• Frequency of severe COVID-19 cases (except for the cases that occurred during the first 14 days after vaccination).<br>• Frequency of lethal COVID-19 cases (except for the cases that occurred during the first 14 days after vaccination). |

| | |
|---|---|
| | 3. To evaluate the reactogenicity of the Ad5-nCoV vaccine compared with placebo based on the following:<br>• Frequency and nature of systemic and local immunisation reactions on the day of vaccination and within 7 days after vaccination.<br>4. To evaluate the safety of the Ad5-nCoV vaccine compared with placebo based on the following:<br>• Frequency and nature of adverse events (Day 0–Day 28) and serious adverse events (Day 0–Day 28; Day 0–Month 6).<br>• Vital signs.<br>• Physical examination results.<br>• Electrocardiography.<br>• Biochemistry.<br>• Haematology.<br>• Coagulation test.<br>• Urinalysis.<br>• Serum immunoglobulin E concentration. |
| **Study Design** | This is a multicentre, randomised, double-blind, placebo-controlled, parallel-group study. The study will be conducted in the Russian Federation and the Republic of Belarus at about 10 trial sites.<br>Five hundred (500) volunteers will be randomised in two treatment (vaccination) groups in a 3:1 ratio (Ad5-nCoV:Placebo) as follows:<br>• Group 1: a single dose of the Ad5-nCoV vaccine.<br>• Group 2: a single dose of placebo.<br>The study design includes 7 outpatient visits to trial sites and 5 phone calls with the study doctor:<br>• Screening visit (Day −10...−1)<br>• Visit 1 (Day 0) — randomisation, a single administration of the investigational product, volunteer's stay at the trial site for 2 hours after vaccination, a phone call with the study doctor in 5–8 hours after vaccination<br>• Visit 2 (Day 2) — reactogenicity and safety evaluation in 48–72 hours after vaccination<br>• Visit 3 (Day 7) — reactogenicity and safety evaluation in 7 days after vaccination<br>• Visit 4 (Day 14) — safety and immunogenicity evaluation in 14 days after vaccination<br>• Visit 5 (Day 28) — safety and immunogenicity evaluation in 28 days after vaccination<br>• Phone calls with the study doctor in 2, 3, 4 and 5 months after vaccination (safety evaluation)<br>• Visit 6 (Month 6) — safety and immunogenicity evaluation in 6 months after vaccination<br>During each next visit and phone call after vaccination, subjects will be interviewed regarding the signs of acute respiratory viral infection to detect and confirm a COVID-19 case (exploratory protective efficacy evaluation).<br>The investigation of the cellular immune response will be performed in a separate cohort of at least 60 subjects randomised at trial sites in Moscow. |
| **Inclusion Criteria** | Subjects must meet all of the following criteria to be eligible for participation in the study:<br>1. Signed and dated Informed Consent Form for participation in the study.<br>2. Men and women, aged 18–85 years.<br>3. Body mass index 18.5–30.0 kg/m$^2$.<br>4. Negative SARS-CoV-2 RNA PCR test at screening.<br>5. Negative SARS-CoV-2 IgM and IgG antibody test at screening. |

| | |
|---|---|
| | 6. No history of COVID-19.<br>7. No close contacts with persons suspected for SARS-CoV-2 infection or persons with laboratory-confirmed COVID-19 within the last 14 days.<br>8. No signs of respiratory infection within the last 14 days.<br>9. Negative tests for human immunodeficiency virus (HIV), syphilis, HBV and HCV. |
| | 10. No history and screening examination findings of diseases and/or conditions that, in the investigator's opinion, may have an impact on the safety of the subject in the study and the assessment of the study results.<br>11. Subject's consent to use reliable methods of contraception during the entire study period. |
| **Exclusion Criteria** | Subjects cannot be included in the study in the case of meeting any of the following criteria:<br>1. Positive allergological anamnesis, drug hypersensitivity, including hypersensitivity to any component of the study drug, as well a history of serious adverse events to vaccines (such as allergic reactions, respiratory disturbance, angioedema, abdominal pain).<br>2. Axillary temperature ≥37.1 °C at screening/randomisation.<br>3. Systolic blood pressure > 139 mm Hg or < 100 mm Hg and/or diastolic blood pressure > 90 mm Hg or < 60 mm Hg.<br>4. Clinically relevant abnormalities during laboratory and/or instrumental examinations at screening.<br>5. Acute infectious diseases less than 4 weeks before screening.<br>6. Acute diseases or exacerbations of chronic diseases of the liver, kidneys, gastrointestinal, cardiovascular, respiratory, nervous or endocrine system.<br>7. History of moderate and severe asthma or pulmonary fibrosis.<br>8. History of blood or hematopoietic disorders.<br>9. History of diabetes mellitus.<br>10. History of epilepsy, epileptic syndrome or convulsive seizures.<br>11. Congenital or acquired immunodeficiency, HIV infection, lymphoma, leukaemia, lupus erythematosus, juvenile rheumatoid arthritis or a history of other autoimmune disorders.<br>12. History of malignant neoplasms.<br>13. Administration of immunotropic drug products (immunomodulatory agents, immunostimulants, immunosuppressants), allergy medications and cytotoxic drugs for > 10 consecutive days within the last 3 months (except for inhaled and topical glucocorticosteroids [GCs]) or the administration of these drugs less than 4 weeks before screening.<br>14. Administration of immunoglobulins or transfusion less than 3 months before screening.<br>15. Administration of antipyretics (including nonsteroidal anti-inflammatory drugs and anilides) within 24 hours before randomisation.<br>16. Blood donation or blood loss (≥450 mL of blood or plasma) less than 3 months before screening.<br>17. Vaccination within 6 months before screening or unwillingness to skip any other vaccination during the study.<br>18. History of alcohol and drug use/drug abuse or mental disorders.<br>19. Major surgery scheduled for the next 6 months or performed within the last 6 months before screening.<br>20. Piercings, permanent makeup/tattoos made less than 1 month before screening or during the study.<br>21. Pregnancy or lactation.<br>22. Participation in another clinical trial within 3 months before screening. |

| | |
|---|---|
| | 23. Psychological, physical or other reasons not allowing the subject to comply with conditions and procedures of the Protocol.<br>24. Subjects who are employees of healthcare facilities and in contact with persons diagnosed with COVID-19.<br>25. Subjects who are employees at the trial site.<br>26. Other reasons not allowing the subject to participate in the study in the opinion of the study doctor. |
| **Dosage and Administration of Investigational Products** | The investigational product (Ad5-nCoV or placebo) will be administered intramuscularly in the deltoid muscle of the upper arm as a single dose of 0.5 mL (1 pre-filled syringe). |
| **Duration of Participation** | The total duration of the subject's participation in the study will be about 6.5 months (about 205 days). |
| **Sample Size and Justification** | To determine the number of subjects required to test the hypothesis relating to the primary variable (proportion of subjects with seroconversion on Day 28 after vaccination), the following conservative assumptions are used:<br>- study power is about 90 %<br>- two-sided significance level (alpha) is 5 %; Pocock's corrected alpha level for multiple comparisons of the primary variable due to an interim analysis is 0.02616 and 0.03039 for the interim and final analysis (performed after obtaining primary variable data for all randomised subjects), respectively<br>- proportion of subjects with seroconversion in the placebo group is 20 %<br>- proposed difference between the placebo and vaccine group in terms of the primary variable is 30–60 % (odds ratio is 4–16)<br>- randomisation ratio between the vaccine and placebo group is 3:1, respectively<br>- two countries<br>- dropout from the study/statistical analysis during the observation period of 28 days is about 10 %.<br>According to the results of the sample size calculation performed using the PASS 12 software (Professional License, NCSS LLC (www.ncss.com)), 180 subjects should be included in the statistical analysis to ensure the power of 90 % for the between-group comparison of the primary variable considering the assumption of 20 % seroconversion rate in the placebo group and the superiority of the vaccine group of at least 30 % (conservative assumption, odds ratio = 4), as well as the corrected two-sided significance level of 0.02616 (one-sided level of 0.01308) and randomisation ratio of 3:1. The number of subjects corresponding to the conservative estimate of differences between the vaccine and placebo groups in terms of the primary variable is chosen to ensure the provision of safety data. Considering the possible dropout from the study/statistical analysis during the observation period of 28 days (10 % of subjects), the number of randomised subjects should be increased up to 200 (in a 3:1 ratio).<br>To provide more detailed safety and efficacy data (including age subgroups), it is planned to randomise 500 subjects (in a 3:1 ratio). Additionally, the extended sample size will allow to provide descriptive data on exploratory endpoints related to the frequency of confirmed COVID-19 cases within 6 months after vaccination (except for the cases occurred during the first 14 days after vaccination).<br>Because the study includes an unblinded interim analysis after obtaining a fraction of information ($\tau = 0.4$) for the assessment of the primary variable (based on the data obtained approximately from the first 200 randomized volunteers through Visit 5, including early dropouts before Visit 5) and the Pocock alpha spending function is used, to adjust the significance level due |

| | |
|---|---|
| | to multiple comparisons of the primary variable (interim analysis), the two-sided alpha level for the interim and final analysis is set to be 0.02616 and 0.03039, respectively (overall two-sided significance level is 5 %). |
| **Interim Analysis** | One unblinded interim analysis is planned for the study, which will be performed after obtaining efficacy, reactogenicity and safety data through Visit 5 (Day 28) for the first 200 volunteers randomised in the study. The analysis will be performed by an independent statistician. If statistically significant findings of the between-group comparison of the primary variable are obtained during the interim analysis, the study will not be suspended. The results of the interim analysis will be provided to regulatory authorities to make a decision on the registration of the Ad5-nCoV vaccine. |

### 3 INTRODUCTION

#### 3.1 General Information

At the end of 2019, there were reports on the outbreak of pneumonia in Wuhan, China caused by a new type of coronavirus, SARS-CoV-2. On 30 January 2020, the World Health Organisation declared this outbreak a public health emergency of international concern and a pandemic on 11 March. On 11 February 2020, the disease was officially named as COVID-19 (coronavirus disease 2019). As of 20 July 2020, there were 14,348,858 confirmed cases of the infection and 603,694 confirmed deaths according to the WHO. In the Russian Federation as of 20 July 2020, there were 777,486 confirmed cases of the disease and 12,427 confirmed deaths. In the Republic of Belarus, there were 66,095 cases of COVID-19 and 499 confirmed deaths (https://www.who.int/emergencies/diseases/novel- coronavirus-2019).

The novel coronavirus belongs to the β genus of coronavirus. Covered by an envelope, the shape of particles could be round or oval, often polymorphic, with a diameter of 60 to 140 nm. Its genetic characteristics are significantly different from SARSr-CoV and MERSr-CoV. The gene sequences (bat-SL-CoVZC45 and bat-SL-CoVZXC21) of the two bats in Zhoushan region of China have 88 % homology by Chinese scientists. The novel coronavirus discovered in Wuhan currently is the seventh coronavirus that can infect humans, and it has not been found in humans before.

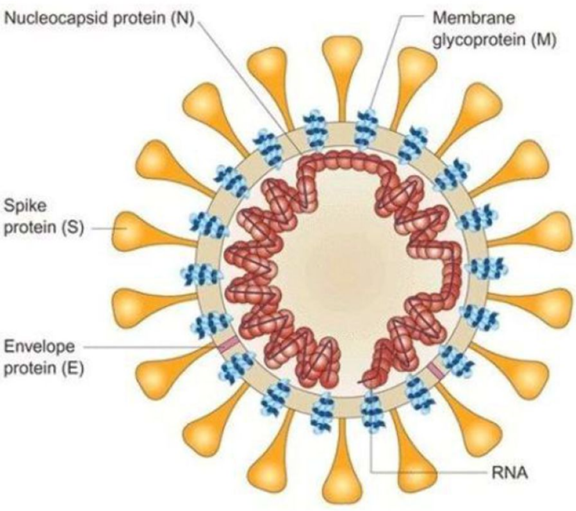

**Figure 1 Schematic of Coronavirus**

As shown in Figure 1, the CoV contains the spike protein (S), envelope protein (E), membrane protein (M), and nucleocapsid protein (N).

Among them, S protein is the most important surface protein of coronavirus, which plays a major role in the development of the infection. The S protein contains two subunits, S1 and S2. S1 protein contains the receptor-binding domain (RBD) and is responsible for identifying cellular receptors. And S2 protein contains the basic elements required for the membrane fusion process.

The virus uses the same receptor, angiotensin-converting enzyme 2 (ACE2) as that for SARS-CoV, and mainly spreads through the respiratory tract. The main symptoms of the disease are fever, dry cough and fatigue. A few patients have symptoms such as nasal congestion, runny nose, sore throat, myalgia, and diarrhoea. Patients with severe infections usually have dyspnoea and/or hypoxemia one week after the onset of symptoms, and most severe patients can quickly develop severe acute respiratory distress syndrome, septic shock, metabolic acidosis that is difficult to correct, as well as coagulation dysfunction and functional failure of multiple organs causing death. The COVID-19 pandemic started in early 2020 and has caused significant disruptions to all countries around the world. At present time, there is no effective treatment for COVID-19, and no approved vaccine to prevent this disease. There are no specifically approved drugs for COVID-19 (except approved for Emergency Authorisation Use during the pandemic). It is necessary and very urgent to develop preventive vaccines against COVID-19 to meet global needs.

In early 2020, CanSino Biologics Inc. and the Beijing Institute of Biotechnology jointly developed a recombinant novel coronavirus vaccine (Ad5-nCoV) to respond to the COVID-19 pandemic situation. Building on the adenovirus vector technology platform that was previously used for the licensed Ebola virus disease vaccine (Ad5-EBOV), this vaccine is a recombinant replication-defective human Ad5 vector-based vaccine expressing spike S protein from SARS-CoV-2. It can stimulate human immune systems to create antibodies and cellular immune responses against the virus and COVID-19 disease.

The pre-clinical study results demonstrated that the Ad5-nCoV vaccine is immunogenic and can effectively express coronavirus S-protein antigen in target cells and can induce strong humoral and cellular immune responses in animal models including mice, guinea pigs, SD rats, and cynomolgus monkeys. There is a strong correlation between the dosage levels and the immune responses in the animal models. In the animal challenge studies, the results demonstrate that the novel coronavirus vaccine Ad5-nCoV has good protection against live coronavirus challenges and can reduce viral infection significantly in hACE2 transgenic mice, ferrets and rhesus monkey models. Besides, from the acute toxicity study in rats and repeated dose toxicity study in cynomolgus monkeys, the results demonstrate that Ad5-nCoV vaccine has a very good safety profile.

In China, two clinical trials of Ad5-nCoV in adults aged 18 years and older are currently approved and conducting: phase I study involving 108 volunteers (started in March 2020) and phase II study involving 508 volunteers (started in April 2020). The interim results of phase I and II clinical trials including data of all enrolled volunteers in 28 days after vaccination indicate that Ad5-nCoV has a favourable safety profile and high immunogenicity. The study plans include a 6-month follow-up period that is currently ongoing.

NPO Petrovax Pharm LLC has entered into an Agreement for Joint Clinical Development and Manufacture and Quality Technical Agreement with CanSino Biologics Inc. (China, a cosponsor of the trial), based on which it has received the right to organise the conduct of the clinical trial of recombinant vaccine Ad5-nCoV as a Sponsor, manufacture it from an intermediate product provided by CanSino Biologics Inc., as well as to register and sell it in the territory including the territory of the Eurasian Economic Union (EAEU).

This trial is conducted to provide evidence on the efficacy, reactogenicity and safety of recombinant novel coronavirus vaccine Ad5-nCoV in healthy volunteers aged from 18 to 85 years in the Eurasian Economic Union. The results of the trial will be provided to the EAEU regulatory authorities for registration of the Ad5-nCoV vaccine.

#### 3.2 Description of the Investigational Product

The Ad5-nCoV vaccine consists of the replication-defective recombinant human type 5 adenovirus expressing spike protein (S protein) of novel coronavirus SARS-CoV-2.

The S protein has been chosen as the antigen based on the experience in developing vaccines against SARS (severe acute respiratory syndrome) and MERS (middle east respiratory syndrome). The study results of these vaccines show a strong immune response and high protective efficacy of S protein vaccines [1]. Despite that the receptor-binding domain (RBD) is the main target for coronavirus neutralisation, neutralisation epitopes have been also identified outside the RBD [2,3]. Adenovirus vector used in Ad5-nCoV is a highly effective and well-known platform for vaccine antigen delivery. Currently, several adenovirus vector vaccines against novel coronavirus are under development that use S protein as the antigen [4].

**Dosage form:** solution for intramuscular injection.

**Formulation:** *Active ingredient:* one dose (0.5 mL) contains $\geq 4 \times 10^{10}$ VP (target $5 \times 10^{10}$ VP).
*Excipients:* mannitol, sucrose, sodium chloride, magnesium chloride, HEPES, polysorbate 80 and glycerine.

**Manufacturer:**

Manufacturing and primary packaging: CanSino Biologics Inc., floor 3 and 4, 185 South Ave., TEDA West District, Tianjin, China. 16 Xinwei Road, TEDA West District, Tianjin, China.

Secondary packaging and release quality control: NPO Petrovax Pharm LLC, 1, Sosnovaya St., Moscow region, Podolsk, Pokrov village 142143, Russia.

#### 3.3 Summary of Pre-clinical and Clinical Studies

##### *3.3.1 Pre-clinical Studies of Ad5-nCoV*

###### *3.3.1.1 Immunogenicity*

It was shown that administration of Ad5-nCoV could stimulate S protein-specific antibody response as well as cellular immune response. Antibodies raised by Ad5-nCoV could neutralise wild type SARS-CoV-2 virus strains. No antibody-dependent enhancement was observed in the preclinical challenge studies.

###### *3.3.1.1.1 Immunogenicity in Mice*

BALB/c mice were intramuscularly immunised with low dose ($5x10^7$ VP), medium dose ($5x10^8$ VP), and high dose ($5x10^9$ VP) of the recombinant Ad5-nCoV vaccine, with 10 mice in each group. The vector adenovirus (Ad5-NULL) was used as control. Samplings were performed at Day 9, Day 14, Day 28, Day 42, Day 56, Day 84, Day 112, Day 140 and Day 168 to detect the specific antibodies (ELISA) against S protein and neutralising antibodies (Vero E6 culture, $EC_{50}$) against SARS-CoV-2. The cellular immune response of $5x10^8$ VP recombinant Ad5-nCoV vaccine at Day 14 after vaccination were evaluated by ELISpot and intracellular staining.

The results showed that on Day 9 after immunisation, the geometric mean titer of S protein antibodies in the high-dose, medium-dose and low-dose group were 137,205 ± 40,120, 57,900 ± 15,950 and 10,961 ± 7258, respectively. In 14 days after immunisation, the geometric mean titer of antibodies in the high-dose, medium-dose and low-dose group were 220,331 ± 59,612, 73,608 ± 14,783 and 27,025 ± 15,076, respectively. In 28 days after immunisation, the geometric mean titer of antibodies in the high-dose, medium-dose and low-dose group were 242,807 ± 58,331, 163,003 ± 35,280 and 50,557 ± 26,854, respectively. Good immunogenicity and dose-response of antibodies were achieved. The antibody levels were increased from Day 9 to Day 14 and reached the highest level at Day 28 in all three groups (Figure 2). These results are consistent with the Ad5-EBOV vaccine in mice. Furthermore, the antibody titers of Ad5-nCoV were significantly higher (10–50 folds) compared to Ad5-EBOV.

In 14 days after immunisation, neutralising antibodies in the medium and high-dose groups were analysed. The geometric mean titer (GMT) of neutralising antibodies in the medium and high-dose group was 13 ± 27 and 58 ± 43, respectively. The results show a correlation between the dose and the neutralising antibody levels.

On Day 14 after immunisation, the splenocytes were stimulated with S protein-peptide pool. The expressions of IFN-γ, TNF-α and IL-2 (both by CD4+ and CD8+ T cells) in the immunised mice were significantly higher than that of the control group (P < 0.001) (Figures 3–4). These results suggest that intramuscular injection of the Ad5-nCoV vaccine can induce strong cellular immune responses in mice.

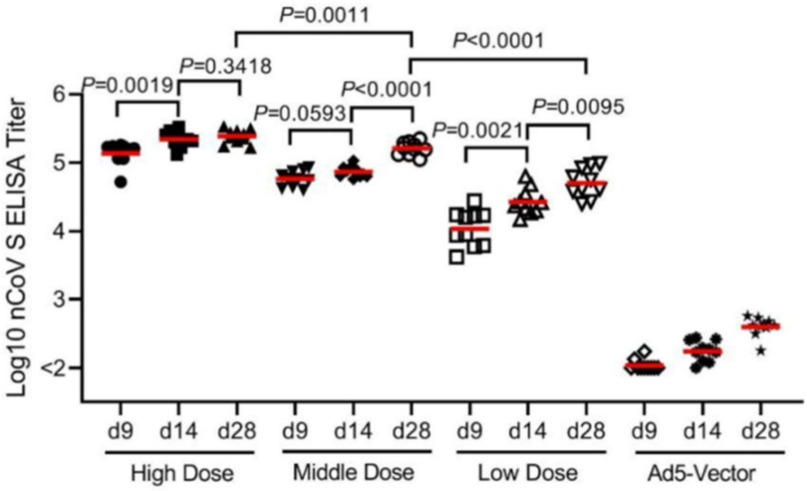

**Figure 2 IgG Antibodies against S Protein on Day 9, Day 14 and Day 28 after Single Immunisation**

1. The analyses were performed for 10 BALB/c mice from the low-dose, medium-dose and high-dose group, as well as the Ad5 vector control group.
2. S protein antibodies on Day 9, 14 and 28 post immunisation were detected using ELISA.

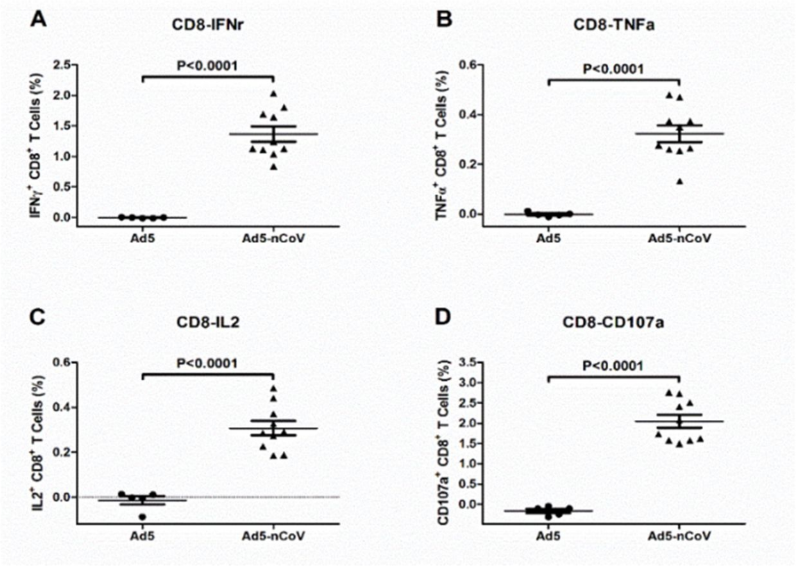

**Figure 3 CD8+ T Cellular Immune Response in Mice Induced by Ad5-nCoV**

1. The analyses were performed for 10 BALB/c mice in the Ad5-nCoV group and 5 BALB/c mice in the Ad5 vector control group.
2. Percentage of CD8+ T cells secreting IFN-γ, TNF-α, IL-2 and CD107a.
3. IFN = interferon. TNF = tumour necrosis factor. IL = interleukin. CD107a = also called lysosome-associated membrane protein 1 (LAMP-1).

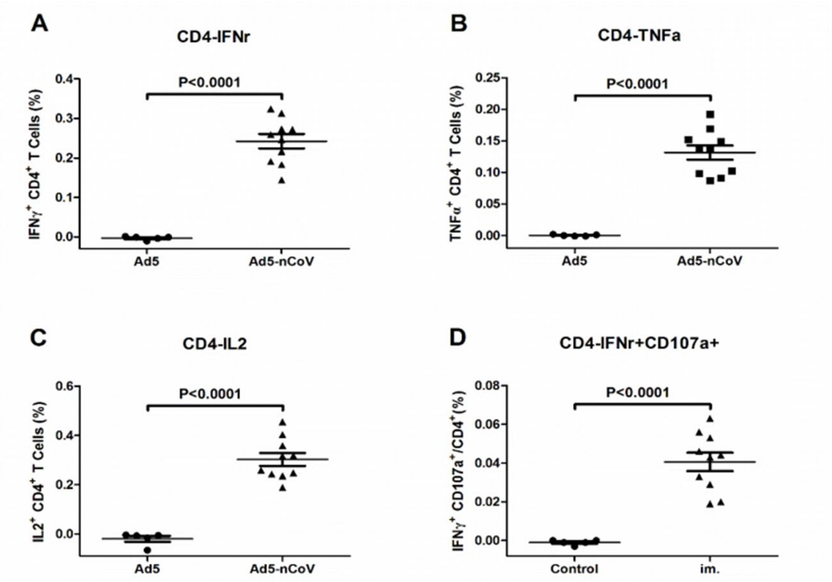

**Figure 4 CD4+ T Cellular Immune Response in Mice Induced by Ad5-nCoV**

1. The analyses were performed for 10 BALB/c mice in the Ad5-nCoV group and 5 BALB/c mice in the Ad5 vector control group.
2. Percentage of CD4+ T cells secreting IFN-γ, TNF-α and IL-2.
3. Percentage of CD4+ T cells secreting the combination of IFN-γ and CD107a.
4. IFN = interferon. TNF = tumour necrosis factor. IL = interleukin. CD107a = also called lysosome-associated membrane protein 1 (LAMP-1).

##### *3.3.1.1.2 Immunogenicity in Guinea Pigs*

Guinea pigs were intramuscularly immunised with low dose (5x$10^7$ VP), medium dose (5x$10^8$ VP), and high dose (5x$10^9$ VP) of the recombinant Ad5-nCoV vaccine, with 10 or 14 guinea pigs in each group. The vector adenovirus (Ad5-NULL) was used as control. Samplings were performed on Day 14, Day 28, Day 42, Day 56, Day 84, Day 112, Day 140 and Day 168 to detect specific antibodies (ELISA) against S protein and neutralising antibodies (Vero E6 culture, $EC_{50}$) against SARS-CoV-2.

The results showed that on Day 14 after single immunisation, the GMT of IgG antibodies against S protein in the high-dose, medium-dose and low-dose group were 43,386 ± 27,575, 36,801 ± 31,736 and 9997 ± 8784, respectively. On Day 28 after single immunisation, the GMT of IgG antibodies against S protein in the high-dose, medium-dose and low-dose group were 164,408 ± 84,483, 87,953 ± 37,944 and 34,551 ± 21,686, respectively. The results are shown in Figure 5 below. The results demonstrated very good immunogenicity and dose correlations of IgG antibodies in guinea pigs after a single dose of novel coronavirus vaccine Ad5-nCoV.

Neutralising antibodies were tested in the high-dose group. The GMT of neutralising antibodies on Day 14 after a single dose in guinea pigs was 28 ± 18. Neutralising antibodies were detected in all of the test animals.

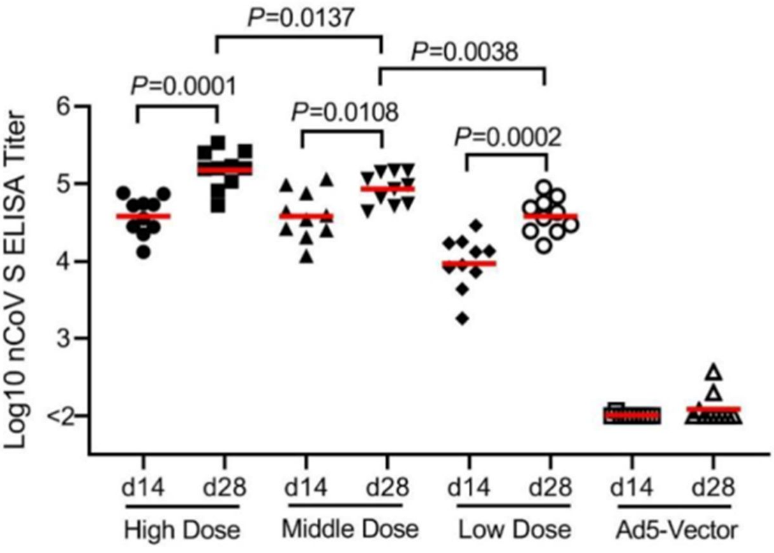

**Figure 5 IgG Antibodies against S protein in Guinea Pigs on Day 14 and Day 28 after Immunisation**

1. The analyses were performed for 14 guinea pigs in the low-dose, medium-dose, high-dose group, and 10 guinea pigs in the Ad5 vector control group.
2. S protein antibodies on Day 14 and 28 post immunisation were detected using ELISA.

##### *3.3.1.1.3 Immunogenicity in Rats*

Sprague Dawley rats were vaccinated with a single human dose (5x$10^{10}$ VP) of Ad5-nCoV by intramuscular injection (the study was conducted by JOINN Laboratories, Inc.). There were 10 rats in each group, half male and half female. One group received one human dose (5x$10^{10}$ VP) of Ad5-nCoV, and the other group was the control group. Blood samples were collected 15 days after immunisation, and the specific IgG antibodies against S protein were tested using an indirect ELISA method.

The results showed that the Ad5-nCoV vaccine had good immunogenicity. The GMT of IgG antibodies on Day 15 was 50 and 407,043 ± 8289 in the control and Ad5-nCoV group, respectively.

##### *3.3.1.1.4 Immunogenicity in Cynomolgus Monkeys*

In the study of repeated injections with a 2-week interval and a recovery period of 2 weeks conducted by JOINN Laboratories, cynomolgus monkeys were immunised with a single human dose (5x$10^{10}$ VP) or 3 human doses (15x$10^{10}$ VP) of the Ad5-nCoV vaccine, respectively. Blood samples of test monkeys were collected in 8, 11 and 15 days after immunisation, respectively. The specific IgG antibodies against S protein and adenovirus vector were tested using indirect ELISA and the neutralising antibodies were tested with live coronavirus.

The results showed that the Ad5-nCoV vaccine had good immunogenicity in cynomolgus monkeys. The control group (10/10) had no detectable IgG antibodies against S protein and no detectable IgG antibodies against adenovirus. In 10 days after immunisation (on Day 11), the positive rate of antibodies in the high dose group (3 human doses) was 100 % (10/10). The levels of IgG antibodies against S protein on Day 8, Day 11 and Day 15 are shown in Figure 6. Dose correlation of antibodies was detected, and the antibody level increased with the increase of the immunisation duration.

Also, cellular responses were elicited. Especially, CD8+ T and CD4+ T IFN-γ expression levels were significantly higher in test vaccine groups compared to the control group (Figure 7).

The results showed that the Ad5-nCoV vaccine-induced neutralising antibodies on Day 11 after a single immunisation. The average neutralising antibody titers ($EC_{50}$) in the high-dose group ($15x10^{10}$ VP) and the low-dose group ($5x10^{10}$ VP) on Day 11 after immunisation were 192 ± 303 and 61 ± 195, respectively. The results showed a correlation between the dose and the neutralising antibody levels.

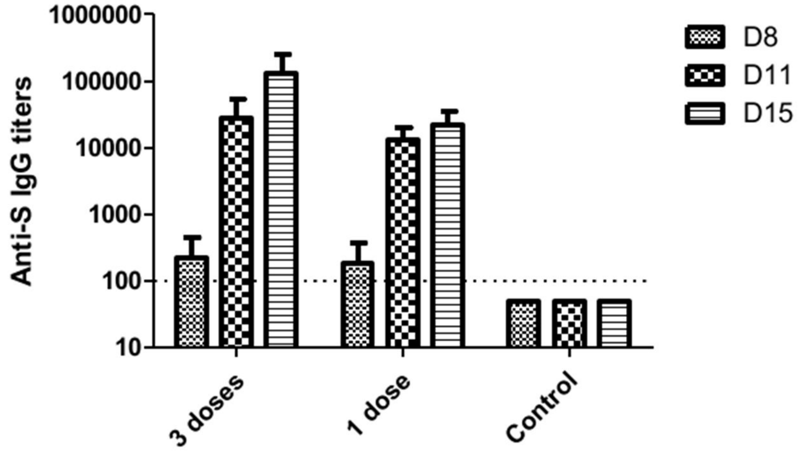

**Figure 6 Antibodies against S Protein on Day 8, 11 and 15 after Immunisation with Ad5-nCoV in Cynomolgus Monkeys**

The analyses were performed for 10 cynomolgus monkeys in the test vaccine group and 10 cynomolgus monkeys in the control group.

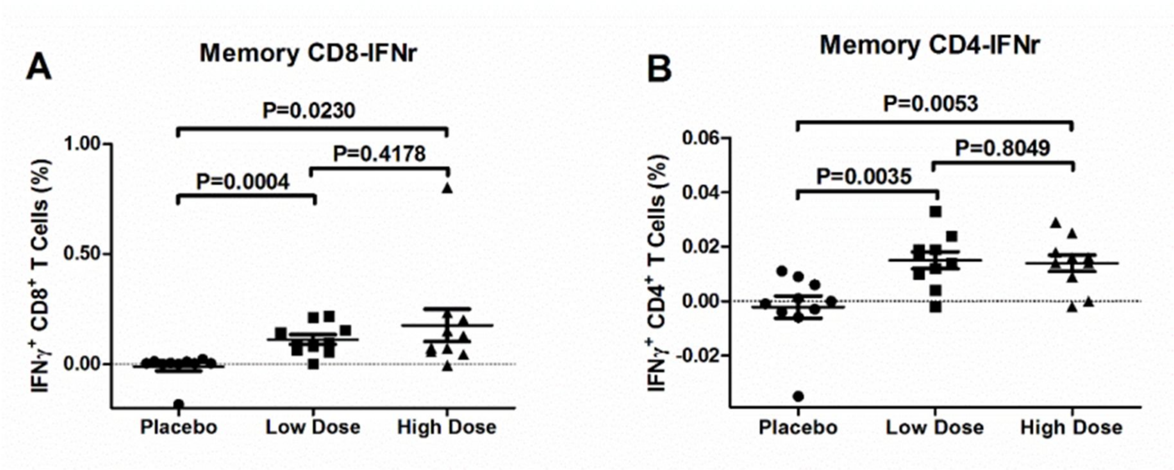

**Figure 7 Cellular Immune Responses after Immunisation in Cynomolgus Monkeys**

1. The analyses were performed for 10 cynomolgus monkeys in each group (low-dose and high-dose Ad5-nCoV group, as well as the placebo control group).
2. Percentage of memory CD4+ and CD8+ T cells secreting IFN-γ.
3. IFN = interferon.
4. Low dose = 1 dose ($5x10^{10}$ VP)

High dose = 3 doses ($15x10^{10}$ VP)

##### *3.3.1.2 Protection*

The protection studies in mice and rhesus monkeys were carried out by the Institute of Medical Laboratory Animals of the Chinese Academy of Medical Sciences. The protection studies in ferrets were performed by the Harbin Veterinary Research Institute of the Chinese Academy of Agricultural Sciences.

###### *3.3.1.2.1 Protection in hACE2 Transgenic Mice*

Eighteen (18) female hACE2 transgenic mice were randomly divided into 3 groups, including 6 mice in the high-dose group ($5x10^9$ VP/mouse), 6 mice in the low-dose group ($5x10^8$ VP/mouse) and 6 mice in the control group. All of the mice were intramuscularly immunised with a single dose of Ad5-nCoV on Day 0 (100 µL/mouse). Blood samples were collected on Day 0 before immunisation, on Day 14 after immunisation and the sacrifice day. Serum samples were obtained for antibody testing. Challenge with live coronavirus (SARS-CoV-2/WH-09/human/2020/CHN, $10^5TCID_{50}$/mouse) was performed on Day 14. After the challenge, body weight of the animals was observed and recorded. The viral load was tested using PCR (copies) in 3 days after the challenge. Immunopathological changes in lung tissues were evaluated by slide staining and microscopic observation.

Body weight of mice in the control group decreased by 3.36 %. In this group, the viral load in the lung tissue was $10^{6.18}$ copies/mL, and changes related to moderate interstitial pneumonia were observed.

Compared to the control group, body weight of mice in the high-dose group ($5x10^9$ VP/mouse) increased by 2.55 %, and no obvious symptoms were found. The viral load in the lung tissue was $10^{3.11}$ copies/mL, with a decrease by 3.07 log. Mild interstitial pneumonia with reduced lesions was observed in the lung tissues.

Compared to the control group, body weight of mice in the low-dose group ($5x10^8$ VP/mouse) decreased by 4.72 %, and no obvious symptoms were found. The viral load in the lung tissue was $10^{3.90}$ copies/mL, with a decrease of 2.28 log. Mild interstitial pneumonia with reduced lesions was observed in the lung tissues.

The above results suggest that a high dose of the Ad5-nCoV vaccine has obvious protection effect in immunised mice and a low dose of the Ad5-nCoV vaccine has significant antiviral effects. The Ad5-nCoV vaccine can effectively reduce the viral load of SARS-CoV-2 and a high dose of the Ad5-nCoV vaccine provides good protection in the transgenic mice model.

###### *3.3.1.2.2 Protection in Ferrets*

Eighteen (18) ferrets were randomly divided into three groups, including 6 ferrets in the high-dose group ($2x10^{10}$ VP), 6 ferrets in the low-dose group ($2x10^9$ VP) and 6 ferrets in the control group. Single intramuscular injection of the test vaccine or control article was performed on Day 0. In 14 days after immunisation, an intranasal challenge with the live novel coronavirus (SARS-CoV-2/CTan/human/2020/Wuhan, $10^5$ PFU/ferret) was performed in ferrets.
Blood samples were collected before immunisation, 14 days after immunisation and on the sacrifice day. Serum was obtained for antibody testing.

The results demonstrated that the ferrets produced IgG antibodies and neutralising antibodies in 14 days after immunisation with the Ad5-nCoV vaccine. When the ferrets were challenged with the SARS-CoV-2 virus in 14 days after vaccination, both low-dose and high-dose groups had much lower viral loads in the upper respiratory tract, and the virus was cleared out more quickly compared to the control group.

**Table 1 Protection Effect in Ferrets: Viral Load in Nasal Wash after Challenge (qPCR, Lg/mL)**

| | **2 days after challenge** | | | **4 days after challenge** | | | **6 days after challenge** | | | **8 days after challenge** | | |
|---|---|---|---|---|---|---|---|---|---|---|---|---|
| | **High dose** | **Low dose** | **Control** | **High dose** | **Low dose** | **Control** | **High dose** | **Low dose** | **Control** | **High dose** | **Low dose** | **Control** |
| Mean | 5.41 | 6.49 | 5.76 | 4.45 | 6.12 | 6.56 | **1.54** | **4.79** | **6.80** | **0.76** | **2.31** | **5.58** |
| SD | 0.52 | 0.95 | 0.89 | 2.23 | 1.17 | 0.69 | 2.38 | 2.48 | 1.03 | 1.85 | 2.53 | 1.13 |

SD = standard deviation

**Table 2 Protection Effect in Ferrets: Viral Load in Nasal Wash after Challenge (Plaque Counting, Log PFU/mL)**

| | 2 days after challenge | | | 4 days after challenge | | | 6 days after challenge | | | 8 days after challenge | | |
|---|---|---|---|---|---|---|---|---|---|---|---|---|
| | High dose | Low dose | Control | High dose | Low dose | Control | High dose | Low dose | Control | High dose | Low dose | Control |
| Mean | 2.69 | 3.20 | 3.07 | 1.91 | 1.98 | 3.18 | 0.67 | 0.34 | 2.88 | 0.36 | 0.00 | 0.36 |
| SD | 0.62 | 0.65 | 0.90 | 0.65 | 0.49 | 0.52 | 1.06 | 0.83 | 0.98 | 0.88 | 0.00 | 0.89 |

SD = standard deviation

Histopathological examination of the lungs and liver after the virus challenge demonstrated the benefit of the Ad5-nCoV vaccine in improving the lung condition after coronavirus challenge. There was no difference in terms of the liver condition after the challenge. No Antibody-Dependent Enhancement (ADE) was found. The study demonstrated that the Ad5-nCoV vaccine had very good immunogenicity, could induce neutralising antibodies in 14 days after vaccination and could provide protection from the SARS-CoV-2 virus in the ferret animal model.

##### *3.3.1.2.3 Protection in Rhesus Monkeys*

Twelve (12) rhesus monkeys were randomly divided into three groups, including 4 in the high-dose group ($20x10^{10}$ VP/monkey), 4 in the low-dose group ($5x10^{10}$ VP/monkey) and 4 in the control group. A single intramuscular injection was performed on Day 0. Blood samples were collected before immunisation, in 14 days after immunisation and on the sacrifice day for blood biochemistry and antibody tests. In 14 days after immunisation, a challenge with the live novel coronavirus (SARS-CoV- 2/WH-09/human/2020/CHN, $10^6$ $TCID_{50}$/monkey) was performed in monkeys. Body weight and body temperature were observed and recorded.

Three days and five days after virus challenge, the pharyngeal swab and anal swab were collected for viral load determination. Five days after the challenge, the lung tissues were collected for viral load determination. Immunopathological changes in lung tissues were evaluated by slide staining and microscopic observation. Serum IgG antibodies (ELISA) and neutralising antibodies were tested.

After the challenge, the decrease in body weight was registered in some monkeys. The body temperature of the monkeys in the control group was not significantly increased after challenge. Compared to the control group, there was a slight increase in the body temperature of three monkeys in the low-dose group. Animals in each group were tested for blood biochemistry before immunisation, 14 days after immunisation, and on the sacrifice day. In 14 days after immunisation, the mean value of uric acid in the low-dose group was 12.14 mmol/L, i. e. significantly higher ($p < 0.01$) than in the negative control group. Other index values were within the normal range. There were no abnormalities in the high-dose group.

In the low-dose and high-dose groups, the viral load in the pharyngeal swab, anal swab and lung tissues was lower than in the control group (Tables 3–5).

**Table 3 Protection Effect in Rhesus Monkeys: Viral Load in Pharyngeal Swab (log10 copies/mL)**

| Group/Time | 3 days after challenge | 5 days after challenge |
|---|---|---|
| Low Dose | 3.72* | 4.33** |
| High dose | 3.04 | 3.56 |
| Negative control | 4.31 | 5.38 |

(Compared with the negative control group, the difference of the test vaccine group is significant *p < 0.05 and very significant **p <0.01.)

**Table 4 Protection Effect in Rhesus Monkeys: Viral Load in Anal Swab (log10 copies/mL)**

| Group/Time | 3 days after challenge | 5 days after challenge |
|---|---|---|
| Low dose | 3.85 | 4.10 |
| High dose | 3.88* | 2.16* |
| Negative control | 4.49 | 5.77 |

(Compared with the negative control group, the difference of the test vaccine group is significant *p < 0.05 and very significant **p <0.01.)

**Table 5 Protection Effect in Rhesus Monkeys: Viral Load in Lung Tissues (log10 copies/mL)**

| Group/Time | Upper left lung | Middle left lung | Lower left lung | Upper right lung | Middle right lung | Lower right lung | Right accessory lung |
|---|---|---|---|---|---|---|---|
| Low dose | 0.00 | 0.00 | 2.36** | 0.00 | 0.00 | 1.18* | 0.00 |
| High dose | 0.00 | 0.00 | 1.22** | 0.00 | 0.00 | 2.03 | 0.00 |
| Negative control | 0.00 | 0.00 | 6.92 | 0.00 | 1.53 | 3.64 | 0.00 |

(Compared with the negative control group, the difference of the test vaccine group is significant *p < 0.05 and very significant **p <0.01.)

##### *3.3.1.3 Toxicology Studies*

###### *3.3.1.3.1 Single Dose Toxicity Study in SD Rats*

A total of 20 rats (10 male and 10 female) were randomly divided into 2 groups: the negative control group and the test vaccine group. Each group had 5 rats per sex. Rats in the test vaccine group received a single dose of Ad5-nCoV ($5x10^{10}$ VP/dose/rat) via intramuscular injection. The control group was administrated with 0.9 % sodium chloride (0.5 mL/rat). A single dose of vaccine (0.5 mL) was injected into the gastrocnemius muscle in two points, with a volume of 0.25 mL each. The mortality/morbidity, clinical manifestations, body weight and food consumption were monitored and recorded after a single immunisation for 14 days. Blood samples were collected 13 days after dosing (on Day 14). The IgG antibody levels against adenovirus vector and specific S protein were determined. All the animals were sacrificed on Day 15, and the gross necropsy was performed.

No deaths or morbidity were registered in the negative control group or one-dose test vaccine group ($5x10^{10}$ VP/dose). There were no abnormalities or toxicity effects on all parameters including clinical manifestations, body weight and food consumption. No obvious abnormal changes in each group were found in the gross necropsy, so no further microscopic examination was performed.

In 13 days after dosing (on Day 14), no IgG antibodies against adenovirus vector and no antibodies against S antigen were detected in the negative control group. In the test vaccine group, high level of S protein-specific antibody titers (1:384,733.095–1:409,600) and low level of antibodies against adenovirus vector (1:50–1:200) were observed.

###### *3.3.1.3.2 Repeated Dose Toxicity Study in Cynomolgus Monkeys*

A total of 30 cynomolgus monkeys were randomly divided into 3 groups (10 animals per group): the negative control group (0.9 % sodium chloride injection, 1.5 mL/monkey), the low dose group (1 dose, $5x10^{10}$ VP, 0.5 mL/monkey) and the high dose group (3 doses, $15x10^{10}$ VP, 1.5 mL/monkey). Each group had 5 rats per sex. Test vaccine and placebo were injected into the right hind limb quadriceps in single or multiple points, with a maximum volume of 0.5 mL each. The immunisation was performed twice with an interval of 2 weeks on Day 1 and Day 15.

After immunisation, clinical manifestations, body weight, body temperature, electrocardiogram, blood pressure, ophthalmoscopic examination, clinical pathology (haematology, coagulation, blood biochemistry and urine analysis), T lymphocyte subsets (CD3+, CD4+, CD8+, CD4+/CD8+), cytokines (IL-2, IL-4, IL-5, IL-6, TNF-α, IFN-γ) and C-reaction protein, complements (C3 and C4) and specific IgG antibodies (against adenovirus vector and S protein) were monitored and tested. Additionally, anti-nuclear antibody spectrum was tested. The monkeys were euthanized 3 days after the last dosing (on Day 18) and at the end of the two-week recovery period (on Day 29). Then, a gross necropsy was performed. The main organs were weighed and the organ-body ratio and the organ-brain ratio were calculated, and microscopic examination of the organs was performed.

No signs of toxicity were found in any group. The No Observed Adverse Effect Level (NOAEL) of Ad5-nCoV is 3 doses ($15x10^{10}$ VP). One week after the first dosing (on Day 8), a low level of adenovirus vector antibody titer and a high level of S protein-specific antibody titer were found in some animals from the test vaccine groups. Before the last dosing (on Day 15), S protein-specific antibodies were found in all animals from the test vaccine groups, with titer ranged from 7471.268 to 382,696.043. After the second dosing, the S protein-specific antibody titer was increased compared to the first dosing and was ranged from 6978.955 to 285,836.154. During the study, no immunotoxicity reactions were observed. The injection site examination showed that after the administration of 1 dose and 3 doses of Ad5-nCoV ($5x10^{10}$ VP/0.5 mL/dose), respectively, slight to mild irritant reactions could be seen in the injection site.

Detailed results of non-clinical studies are given in the Investigator's Brochure.

##### 3.3.2 Clinical Trials of Ad5-nCoV

Test vaccine Ad5-nCoV was approved for clinical trials in China. Currently, there are interim results of phase I and II clinical trials (on immunogenicity and safety in 28 days after vaccination). Detailed results of clinical trials are provided in the Investigator's Brochure.

###### *3.3.2.1 Phase I Clinical Trial in Adults [5]*

A phase I clinical trial (clinicaltrials.gov no. NCT04313127) is being conducted in Wuhan, China from March 2020. This is a single centre, open-label, dose-escalation phase I clinical trial involving volunteers aged from 18 to 60 years.

In May 2020, interim results of the study were published [5][1], including data of all enrolled volunteers in 28 days after vaccination. The study plan includes a 6-month follow-up period that is currently ongoing.

A total of 108 volunteers were enrolled in the study (36 volunteers per group). There were three dose groups: low dose group ($5x10^{10}$ VP), medium-dose group ($10x10^{10}$ VP) and high dose group ($15x10^{10}$ VP). The Ad5-nCoV vaccine was administered intramuscularly in the deltoid muscle of the upper arm as a single dose. The study plan includes visits in 3, 7, 10, 14, 28 days and 3 and 6 months after vaccination.

The primary endpoint is the occurrence of adverse reactions in all groups within 0–7 days after vaccination. The secondary endpoints include the occurrence of unsolicited adverse events in all groups within 0–28 days after vaccination, the occurrence of adverse events in all groups within 0–28 days after vaccination, the occurrence of serious adverse events in all groups within 6 months after vaccination, the change of laboratory indicators from Day 0 to Day 7 after vaccination (including white blood cell count, lymphocyte count, neutrophils, platelets, haemoglobin, alanine aminotransferase, aspartate aminotransferase, total bilirubin, random blood glucose and creatinine), GMT of serum S-specific antibodies (EIA), GMT of neutralising antibodies against SARS-CoV-2, GMT of neutralising antibodies against Ad5, specific cellular immune responses and some other exploratory endpoints.

Gender and age distribution of all subjects is summarised in Table 6.

**Table 6 Phase I Clinical Trial: Gender and Age of All Subjects**

| | | Low dose (N = 36) | Medium dose (N = 36) | High dose (N = 36) | Total (N = 108) |
|---|---|---|---|---|---|
| Sex | | | | | |
| Male | n (%) | 18 (50.00 %) | 19 (52.78 %) | 18 (50.00 %) | 55 (50.93 %) |
| Female | n (%) | 18 (50.00 %) | 17 (47.22 %) | 18 (50.00 %) | 53 (49.07 %) |
| Total | N (MISS) | 36 (0) | 36 (0) | 36 (0) | 108 (0) |
| Age (years) | | | | | |
| [18–30] | n (%) | 9 (25.00 %) | 12 (33.33 %) | 10 (27.78 %) | 31 (28.70 %) |
| [30–40] | n (%) | 13 (36.11 %) | 14 (38.89 %) | 15 (41.67 %) | 42 (38.89 %) |
| [40–50] | n (%) | 8 (22.22 %) | 3 (8.33 %) | 7 (19.44 %) | 18 (16.67 %) |
| [50–60] | n (%) | 6 (16.67 %) | 7 (19.44 %) | 4 (11.11 %) | 17 (15.74 %) |
| Total | N (MISS) | 36 (0) | 36 (0) | 36 (0) | 108 (0) |

*Safety Results*

At least one adverse reaction within the first 7 days after vaccination was reported in 87 (81 %) of 108 participants: in 30 (83 %) subjects in the low-dose group, 30 (83 %) subjects in the medium-dose group, and 27 (75 %) subjects in the high-dose group. Among them, severe adverse reactions (grade 3) were registered in 10 (9 %) of 108 participants: in 2 (6 %), 2 (6 %) and 6 (17 %) subjects in the low-dose, medium-dose and high-dose group, respectively.

Information on local adverse reactions by their severity and frequency is provided in Table 7 and Table 8 below. The most common local adverse reaction was pain, which was reported in 58 (54 %) vaccinated participants. Other local reactions included swelling (8 [7 %] subjects), itching (5 [5 %] subjects), redness (4 [4 %] subjects), induration at the injection site (4 [4 %] subjects) and muscle weakness (1 [1 %] sub-

[1] Full version of the article is available at https://www.thelancet.com/action/showPdf?pii=S0140-6736%2820 %2931208-3. Attachments are available at https://www.thelancet.com/cms/10.1016/S0140-6736(20)31208- 3/attachment/d9d5f801-7fa5-4802-99a5-0dfc9b4836f5/mmc1.pdf

ject). Generally, all local reactions were mild to moderate. There were no significant differences between the dose groups.

**Table 7 Overview of Local Adverse Reactions (Day 0–7 after Vaccination)**

| Dose | N | Grade 1 | | Grade 2 | | Grade 3 | |
|---|---|---|---|---|---|---|---|
| | | n | % | n | % | n | % |
| Low | 36 | 16 | 44.4 | 2 | 5.6 | 0 | 0 |
| Medium | 36 | 19 | 52.8 | 3 | 8.3 | 0 | 0 |
| High | 36 | 20 | 55.6 | 2 | 5.6 | 0 | 0 |

**Table 8 Frequency of Local Adverse Reactions (Day 0–7 after Vaccination)**

| Adverse reaction | Low dose | | Medium dose | | High dose | |
|---|---|---|---|---|---|---|
| | Grade 1 | Grade 2 | Grade 1 | Grade 2 | Grade 1 | Grade 2 |
| Pain | 17 (47.2 %) | 0 | 20 (55.6 %) | 0 | 19 (52.8 %) | 2 (5.6 %) |
| Swelling | 2 (5.6 %) | 2 (5.6 %) | 1 (2.8 %) | 3 (8.3 %) | 0 | 0 |
| Itching | 2 (5.6 %) | 0 | 2 (5.6 %) | 1 (2.8 %) | 0 | 0 |
| Redness | 1 (2.8 %) | 1 (2.8 %) | 0 | 1 (2.8 %) | 1 (2.8 %) | 0 |
| Induration | 1 (2.8 %) | 1 (2.8 %) | 0 | 1 (2.8 %) | 1 (2.8 %) | 0 |
| Muscle weakness | 0 | 0 | 0 | 0 | 1 (2.8 %) | 0 |

Information on systemic adverse reactions by their severity and frequency is provided in Table 9 and Table 10 below. The most commonly reported systemic adverse reactions were fever (50 [46 %] subjects), fatigue (47 [44 %] subjects), headache (42 [39 %] subjects), and myalgia (18 [17 %] subjects). Most adverse reactions were mild to moderate.
Severe adverse reactions were registered in 10 (9 %) participants. They included fever (9 [8 %] subjects), fatigue (2 [2 %] subjects), myalgia, arthralgia and dyspnoea (1 [1 %] subject each). There were no significant differences between the groups in terms of the frequency of individual adverse reactions or the frequency of adverse events in general.

Episodes of severe fever (grade 3) with an axillary temperature greater than 38.5 °C were observed in 2 [6 %] participants in the low-dose group, 2 [6 %] participants in the medium-dose group, and 5 [14 %] participants in the high-dose group. Among them, one volunteer (3 %) from the high-dose group reported severe fever accompanied by severe fatigue, dyspnoea and muscle pain. One participant from the high-dose group reported severe fatigue and joint pain. These reactions occurred within 24 hours after vaccination and persisted for not more than 48 hours.

**Table 9 Overview of Systemic Adverse Reactions**

| Dose | N | Grade 1 | | Grade 2 | | Grade 3 | |
|---|---|---|---|---|---|---|---|
| | | n | % | n | % | n | % |
| Low | 36 | 19 | 52.8 | 4 | 11.1 | 2 | 5.6 |
| Medium | 36 | 15 | 41.7 | 9 | 25.0 | 2 | 5.6 |
| High | 36 | 15 | 41.7 | 5 | 13.9 | 6 | 16.7 |

Note: No serious adverse events (SAEs) were reported.

**Table 10 Frequency of Systemic Adverse Reactions**

| AR | Low dose | | | Medium dose | | | High dose | | |
|---|---|---|---|---|---|---|---|---|---|
| | Grade 1 | Grade 2 | Grade 3 | Grade 1 | Grade 2 | Grade 3 | Grade 1 | Grade 2 | Grade 3 |
| Fever | 10 (27.8 %) | 3 (8.3 %) | 2 (5.6 %) | 6 (16.7 %) | 7 (19.4 %) | 2 (5.6 %) | 10 (27.8 %) | 5 (13.9 %) | 5 (13.9 %) |
| Fatigue | 15 (41.7) | 2 (5.6 %) | 0 | 13 (36.1 %) | 1 (2.8 %) | 0 | 11 (30.6 %) | 3 (8.3 %) | 2 (5.6 %) |
| Headache | 13 (36.1 %) | 1 (2.8 %) | 0 | 9 (25.0 %) | 2 (5.6 %) | 0 | 13 (36.1 %) | 4 (11.1 %) | 0 |
| Lost of appetite | 6 (16.7 %) | 0 | 0 | 4 (11.1 %) | 1 (2.8 %) | 0 | 5 (13.9 %) | 1 (2.8 %) | 0 |
| Muscle pain | 6 (16.7 %) | 1 (2.8 %) | 0 | 3 (8.3 %) | 0 | 0 | 5 (13.9 %) | 2 (5.6 %) | 1 (2.8 %) |
| Diarrhoea | 3 (8.3 %) | 0 | 0 | 4 (11.1 %) | 0 | 0 | 5 (13.9 %) | 0 | 0 |
| Joint pain | 2 (5.6 %) | 0 | 0 | 2 (5.6 %) | 0 | 0 | 4 (11.1 %) | 0 | 1 (2.8 %) |
| Cough | 1 (2.8 %) | 0 | 0 | 2 (5.6 %) | 0 | 0 | 3 (8.3 %) | 0 | 0 |
| Sore throat | 1 (2.8 %) | 0 | 0 | 3 (8.3 %) | 0 | 0 | 4 (11.1 %) | 0 | 0 |
| Nausea | 2 (5.6 %) | 0 | 0 | 1 (2.8 %) | 0 | 0 | 2 (5.6 %) | 1 (2.8 %) | 0 |
| Other | 4 (11.1 %) | 0 | 0 | 1 (2.8 %) | 0 | 0 | 6 (16.7 %) | 0 | 1 (2.8 %) |

Note: Fever Grade 1: 37.3–38.0 °C, Grade 2: 38.0–38.5 °C, Grade 3: 38.5–39.5°C.

Overall, within 0–28 days after vaccination, adverse events were registered in 88 (81 %) of 108 participants: 31 (86 %) subjects in the low-dose group, 30 (83 %) subjects in the medium-dose group, and 27 (75 %) subjects in the high-dose group. No serious adverse events were registered as of 30 April 2020.

On Day 7 after vaccination, 9 (8 %) participants had mild to moderate total bilirubin increase, 10 (9 %) participants had alanine aminotransferase increase, and 4 (4 %) participants had fasting hyperglycaemia, but no cases were considered clinically significant.

*Immunogenicity Results*

On Day 28, the GMT of binding antibodies was 1445.8 (95 % CI 935.5–2234.5) in the high-dose group, 806.0 (528.2–1229.9) in the medium-dose group and 615.8 (405.4–935.5) in the low dose group (high dose vs low dose 1611.5, 531.5–2691.5). At least a 4-fold increase of this parameter was noted in 35/36 (97 %) participants in the low-dose group, 34/36 (94 %) participants in the medium-dose group, and 36/36 (100 %) participants in the high-dose group. Neutralising antibodies against live SARS-CoV-2 were all negative on Day 0, increased moderately on Day 14 and peaked in 28 days after vaccination. In the high-dose group, the GMT of neutralising antibodies was 34.0 (95 % CI 22.6–50.1), which was significantly higher than 16.2 (10.4–25.2) in the medium-dose group and 14.5 (9.6–21.8) in the low-dose group. On Day 28, an estimated difference between the high-dose group and the medium-dose group and between the high-dose group and the low-dose group was 27.7 (1.0–54.4) and 33.2 (6.5–59.9), respectively. Meanwhile, at least a 4-fold increase of the neutralising antibody titer by Day 28 was noted in 18/36 (50 %) participants in the low-dose group, 18/36 (50 %) participants in the medium-dose group, and 27/36 (75 %) participants in the high-dose group. Detailed data are provided in the table below.

**Table 11 Specific Antibody Responses to the Receptor-Binding Domain (RBD) and Neutralising Antibodies to Live SARS-CoV-2**

| | Day 14 | | | | Day 28 | | | |
|---|---|---|---|---|---|---|---|---|
| | **Low-dose group (n = 36)** | **Medium-dose group (n = 36)** | **High-dose group (n = 36)** | **p-value** | **Low-dose group (n = 36)** | **Medium-dose group (n = 36)** | **High-dose group (n = 36)** | **p-value** |
| **Antibodies to the receptor-binding domain (ELISA)** | | | | | | | | |
| GMT (mean) | 76.5 | 91.2 | 132.6 | 0.29 | 615.8 | 806.0 | 1445.8 | 0.016 |
| 95 % CI | (44.3–132.0) | (55.9–148.7) | (80.7–218.0) | | (405.4–935.5) | (528.2–1229.9) | (935.5–2234.5) | |
| ≥ 4-fold increase | 16 (44 %) | 18 (50 %) | 22 (61 %) | 0.35 | 35 (97 %) | 34 (94 %) | 36 (100 %) | 0.77 |
| **Neutralising antibodies to live SARS-CoV-2** | | | | | | | | |
| GMT (mean) | 8.2 | 9.6 | 12.7 | 0.24 | 14.5 | 16.2 | 34.0 | 0.0082 |
| 95 % CI | (5.8–11.5) | (6.6–14.1) | (8.5–19.0) | | (9.6–21.8) | (10.4–25.2) | (22.6–50.1) | |
| ≥ 4-fold increase | 10 (28 %) | 11 (31 %) | 15 (42 %) | 0.42 | 18 (50 %) | 18 (50 %) | 27 (75 %) | 0.046 |

1. Data are mean (95 % CI) or n (%). The p values are the result of the comparison between the three dose groups. If the difference between the groups was significant, 95 % CIs were estimated. SARS-CoV-2 = severe acute respiratory syndrome coronavirus.
2. GMT = geometric mean titer.

ELISpot responses at baseline were undetectable with spot-forming cells below the level of detection in all participants but peaked on Day 14 post-vaccination. The proportions of positive responders ranged from 83–97 % across the dose groups, with a mean number of spot-forming cells per 100,000 cells of 20.8 (95 % CI 12.7–34.0) in the low-dose group, 40.8 (27.6–60.3) in the medium-dose group, and 58.0 (39.1–85.9) in the high-dose group. T-cell responses in the high-dose group were significantly higher than that in the low-dose group ($p < 0.0010$), but not significant compared with that in the medium-dose group.

Thus, the interim results of the phase I clinical trial indicate that Ad5-nCoV has a favourable safety profile and can induce a strong immune response against novel coronavirus.

##### *3.3.2.2 Phase II Clinical Trial in Adults [6]*

A phase II clinical trial started on 12 April 2020 in Wuhan, China. This is a single centre, randomised, double-blind, placebo-controlled trial involving volunteers aged 18 years and older. The purpose of the trial is to further evaluate the immunogenicity and safety of the vaccine in healthy volunteers aged 18 years and above. The recruitment into this trial was completed in April 2020. A total of 508 volunteers have been enrolled in the trial, and the follow-up period is still ongoing.

Preliminary results of the phase II clinical trial (as of Day 28 after vaccination) were published by Feng-Cai Zhu et al. in July 2020 [6][2]. The baseline characteristics of the enrolled subjects are shown in the table below.

[2] Full version of the article is available at https://www.thelancet.com/action/showPdf?pii=S0140-6736%2820%2931605-6. Attachments are available at https://www.thelancet.com/cms/10.1016/S0140-6736(20)31605-6/attachment/ef971a01-a648-4890-9fbf-09be76a9643d/mmc1.pdf

**Table 12 Baseline Characteristics of Phase II Clinical Trial Participants**

| | Vaccine at 10x10$^{10}$ VP | Vaccine at 5x10$^{10}$ VP | Placebo |
|---|---|---|---|
| | (n = 253) | (n = 129) | (n = 126) |
| Age, years | | | |
| 18–44 | 152 (60 %) | 80 (62 %) | 77 (61 %) |
| 45–54 | 67 (26 %) | 32 (25 %) | 35 (28 %) |
| ≥ 55 | 34 (13 %) | 17 (13 %) | 14 (11 %) |
| Mean (SD) | 40.0 (12.8) | 39.7 (12.1) | 39.2 (12.5) |
| Sex | | | |
| Male | 126 (50 %) | 64 (50 %) | 64 (51 %) |
| Female | 127 (50 %) | 65 (50 %) | 62 (49 %) |
| Body mass index, kg/m$^2$ | | | |
| Mean (SD) | 24.2 (2.8) | 24.2 (2.7) | 23.3 (2.6) |
| Underlying diseases | | | |
| Yes | 8 (3 %) | 8 (6 %) | 6 (5 %) |
| No | 245 (97 %) | 121 (94 %) | 120 (95 %) |
| Pre-existing Ad5 neutralising antibody titres | | | |
| ≤ 1:200 | 127 (50 %) | 54 (42 %) | 61 (48 %) |
| > 1:200 | 126 (50 %) | 75 (58 %) | 62 (52 %) |

Blood samples were taken on Day 0 immediately after vaccination, on Day 14 and on Day 28 after vaccination to measure the specific antibody titer against the receptor-binding domain (RBD) using ELISA. The limit of detection of specific antibodies against the RBD using ELISA was 1:40. Neutralising antibody titer against live SARS-CoV-2 (SARS-CoV-2/human/CHN/Wuhan_IME-BJ01/2020, GenBank No. MT291831.1) or pseudovirus (spike glycoprotein-expressing pseudovirus system based on the vesicular stomatitis virus) and cellular immune response before vaccination and in 28 days after vaccination were also measured. Limits of detection of neutralising antibodies against live SARS-CoV-2 and pseudovirus were 1:8 and 1:10, respectively. If antibodies were undetectable, a half LoD value was used for calculations. Cellular immune response (IFN-γ expression) stimulated by the pool of spike glycoprotein was evaluated using ELISpot. Positive IFN-γ ELISpot response was defined as ≥ 5 spot-forming cells per 1x10$^5$ peripheral blood mononuclear cells and at least two-fold increase of this parameter compared to the baseline. Titers of neutralising antibodies against the Ad5 vector of the test vaccine were measured by a serum neutralisation test.

*Humoral Immunity Assessment Results*

The results are shown in Figure 8 below. On Day 28 after vaccination, the seroconversion rate (at least four-fold increase in antibody titer compared to the baseline) was 96 % (95 % CI 93–98) in the 10x10$^{10}$ VP group [seroconversion was detected in 244 of 253 volunteers], 97 % (95 % CI 92–99) in the 5x10$^{10}$ VP group [seroconversion was detected in 125 of 126 volunteers], and no antibody titer increase was observed in the placebo group. RBD-specific antibody titer on Day 28 was 656.5 (575.2–749.2) in the 10x10$^{10}$ VP group and 571.0 (467.6–697.3) in the 5x10$^{10}$ VP group.

The seroconversion rate of neutralising antibodies against live SARS-CoV-2 was 59 % (95 % CI 52–65) in the 10x10$^{10}$ VP group and 47 % (95 % CI 39–56) in the 5x10$^{10}$ VP group. No significant differences between the two dose groups in the seroconversion rate of neutralising antibodies against live virus and pseudovirus were observed.

Subgroup analysis of the participants with different pre-existing anti-Ad5 titers showed that the subjects with low pre-existing anti-Ad5 immunity had at about twice higher RBD-specific ELISA antibody and neutralising antibody levels than the participants with high pre-existing anti-Ad5 immunity. Another independent factor affecting the immune response was advanced age (according to the RBD-specific antibody level [$p = 0.0018$] and the level of neutralising antibodies against the live virus [$p < 0.0001$] and pseudovirus [$p = 0.046$]). The results of the analysis stratified by age showed that the participants aged 55 years and above had a relatively low humoral response in both vaccine groups, especially in terms of neutralising antibody titer against live virus. Nevertheless, the RBD-specific antibody titers and neutralising antibody titers on Day 28 in vaccinated participants were significantly higher than in the placebo group. Post-vaccination RBD-specific antibody titers and neutralising antibody titers were similar in both male and female participants from the vaccine groups.

RBD-specific and pseudovirus-neutralising antibody titers significantly correlated to the neutralising antibody titer against live virus. The correlation coefficient was 0.75 and 0.72 (both $p < 0.0001$), respectively.

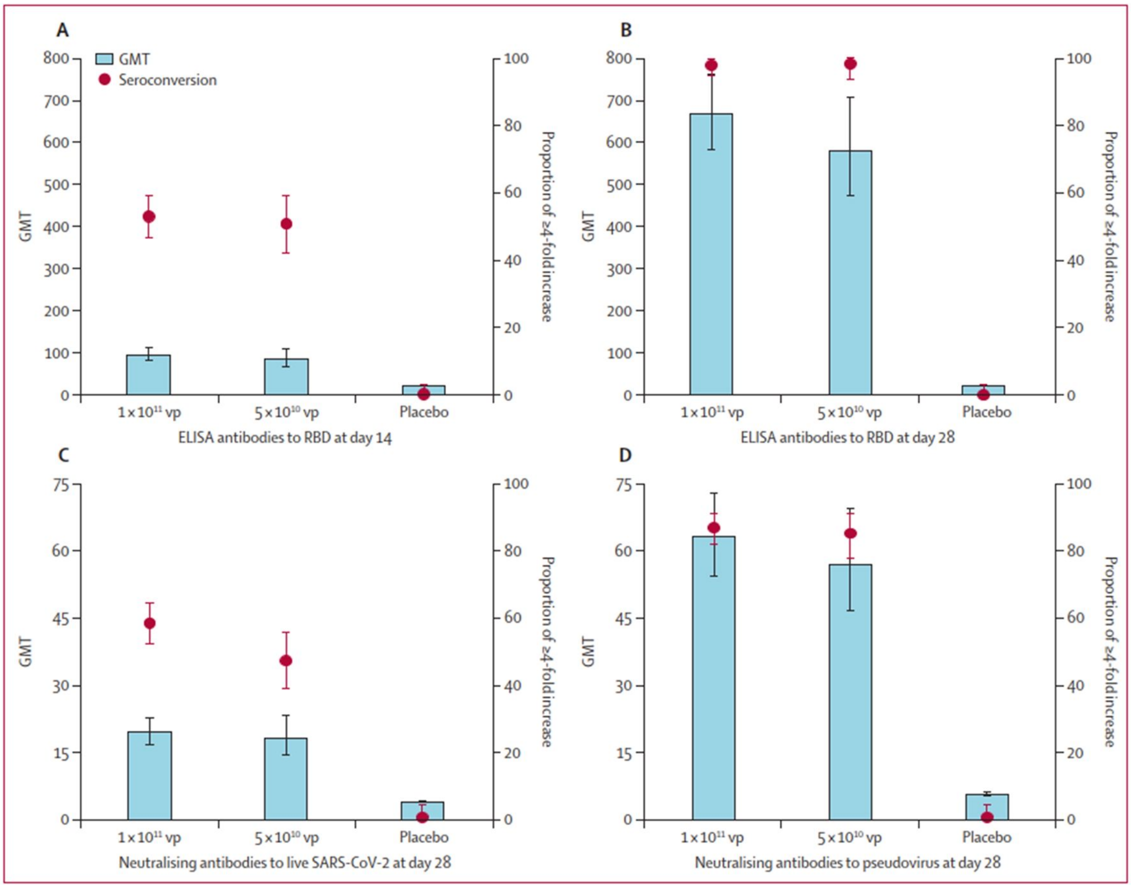

**Figure 8 Post-vaccination Titers of Specific Antibodies against the RBD, Neutralising Antibodies to Live SARS-CoV-2 and Pseudovirus**

GMT = geometric mean titer. RBD = receptor-binding domain. VP = viral particles. Seroconversion was defined as an increase in post-vaccination titer at least four-times compared to the baseline. All the differences between the three groups were highly significant ($p < 0.0001$). Multiple comparisons showed no significant differences between the $1x10^{11}$ VP dose group and the $5x10^{10}$ VP dose group.
$1x10^{11}$ VP = $10x10^{10}$ VP

##### *Cellular Immunity Assessment Results*

Baseline ELISpot T-cell response was negative in > 99 % (506/508) participants. Administration of the Ad5-nCoV vaccine resulted in a significant specific IFN-γ response to the spike glycoprotein of SARS-CoV-2 (ELISpot) on Day 28 after immunisation. The response was detected in 227 participants (90 %, 95 % CI 85–93) in the $10x10^{10}$ VP group and 113 participants (88 %, 95 % CI 81–92) in the $5x10^{10}$ VP group.
On Day 28, a median of spot-forming cells per $1x10^5$ peripheral blood mononuclear cells was 11.0 (interquartile range [IQR] 5.0–25.0) and 10.0 (IQR 6.0–21.0) in the $10x10^{10}$ VP dose group and the $5x10^{10}$ VP dose group, respectively. In both groups, this parameter increased by > 10-fold compared to the baseline.

There was no significant difference in the ELISpot IFN-γ response between the groups on Day 28. No positive T-cell response was observed in the placebo group after vaccination.

Significant T-cell response (spot-forming cells) increase by Day 28 after vaccination was detected in the participants with low and high pre-existing Ad5 neutralising antibody titres. In both groups, positive T-cell response (ELISpot IFN-γ level) after vaccination was observed in 88 % of the participants with a significant pre-existing response to Ad5 (in 111 of 126 subjects in the $10x10^{10}$ VP dose group and 66 of 75 subjects in the $5x10^{10}$ VP dose group, respectively). The T-cell response to vaccination was age and sex independent. Additionally, 241 subjects (95 %, 95 % CI 92–97) in the $10x10^{10}$ VP dose group and 118 subjects (91 %, 95 % CI 85–95) in the $5x10^{10}$ VP dose group showed either positive T-cell response or seroconversion of neutralising antibodies against live SARS-CoV-2 on Day 28 after vaccination.

*Safety Results*

Within 14 days after vaccination, 183 (72 %) participants in the $10x10^{10}$ VP dose group and 96 (74 %) participants in the $5x10^{10}$ VP dose group reported at least one solicited adverse reaction, both of which were significantly higher compared with 46 (37 %) participants in the placebo group ($p < 0.0001$) (see Table below). Severe adverse reactions (grade 3) were registered in 24 (9 %) subjects from the $10x10^{10}$ VP dose group and in one (1 %) subject from the $5x10^{10}$ VP dose group.

The most common local reaction was pain in the injection site. It was reported by 145 (57 %) and 72 (56 %) subjects in the $10x10^{10}$ VP and $5x10^{10}$ VP dose group, respectively.

The most common systemic reactions were fatigue, fever, and headache. They were reported by 106 (42 %), 82 (32 %) and 73 (29 %) subjects in the $10x10^{10}$ VP dose group and by 44 (34 %), 21 (16 %) and 36 (28 %) subjects in the $5x10^{10}$ VP dose group, respectively.

Although most adverse reactions were reported as either mild or moderate, 24 (9 %) participants receiving the $10x10^{10}$ VP dose had severe (grade 3) adverse reactions, which was significantly higher than in the $5x10^{10}$ VP dose group (1 participant [1 %]) or placebo group (0 cases) ($p = 0.0011$ and $p = 0.0004$, respectively). The most common grade 3 adverse reactions included fever reported in 20 (8 %) subjects in the $10x10^{10}$ VP dose group and 1 (1 %) subject in the $5x10^{10}$ VP dose group. Lesser frequency of post-vaccination fever was associated with a high level of pre-existing Ad5 antibody titres, advanced age and male sex. Grade 3 reactions were self-limited and resolved within 72–96 hours without any medication.

The unsolicited adverse reactions occurred within 14 days post-vaccination were reported by 19 (8 %), 7 (5 %) and 7 (6 %) participants in the $10x10^{10}$ VP dose, $5x10^{10}$ VP dose and placebo group, respectively, showing no significant differences between the groups. Overall, 196 (77 %), 98 (76 %) and 61 (48 %) participants receiving the $10x10^{10}$ VP dose, $5x10^{10}$ VP dose and placebo experienced at least one or more adverse events within 28 days after vaccination, respectively. No serious adverse events were documented within this period.

The results indicate that the $5x10^{10}$ VP dose of the vaccine has a more favourable safety profile.

All the participants had no IgG and IgM antibodies against the nucleocapsid protein of SARS-CoV-2 on Day 28, so it could be assumed that no subjects were infected by SARS-CoV-2 during this period.

**Table 13 Adverse Reactions within 14 Days after Vaccination and Overall Adverse Events within 28 Days after Vaccination**

| | Vaccine at $10x10^{10}$ VP (n = 253) | Vaccine at $5x10^{10}$ VP (n = 129) | Placebo (n = 126) | p-value |
|---|---|---|---|---|
| **Solicited adverse reactions within 14 days** | | | | |
| Any | 183 (72 %) | 96 (74 %) | 46 (37 %) | < 0.0001 |
| Grade 3 | 24 (9 %) | 1 (1 %) | 0 | < 0.0001 |
| Injection-site adverse reactions | | | | |
| Pain | 145 (57 %) | 72 (56 %) | 11 (9 %) | < 0.0001 |
| Induration | 12 (5 %) | 2 (2 %) | 0 | 0.014 |
| Grade 3 induration | 2 (1 %) | 0 | 0 | 0.75 |
| Redness | 5 (2 %) | 1 (1 %) | 2 (2 %) | 0.81 |
| Swelling | 10 (4 %) | 5 (4 %) | 0 | 0.049 |
| Grade 3 swelling | 1 (< 1 %) | 0 | 0 | 1.0 |
| Itching | 14 (6 %) | 3 (2 %) | 0 | 0.0075 |
| Systemic adverse reactions | | | | |
| Fever (any grade) | 82 (32 %) | 21 (16 %) | 12 (10 %) | < 0.0001* |
| Grade 3 fever | 20 (8 %) | 1 (1 %) | 0 | 0.0001‡ |
| Headache | 73 (29 %) | 36 (28 %) | 17 (13 %) | 0.0031 |
| Grade 3 headache | 2 (1 %) | 0 | 0 | 0.75 |
| Fatigue | 106 (42 %) | 44 (34 %) | 21 (17 %) | < 0.0001 |
| Grade 3 fatigue | 1 (< 1 %) | 0 | 0 | 1.0 |
| Vomiting | 4 (2 %) | 1 (1 %) | 1 (1 %) | 0.88 |
| Diarrhoea | 19 (8 %) | 10 (8 %) | 4 (3 %) | 0.22 |
| Muscle pain | 39 (15 %) | 23 (18 %) | 3 (2 %) | 0.0002 |
| Grade 3 muscle pain | 1 (< 1 %) | 0 | 0 | 1.0 |
| Joint pain | 34 (13 %) | 13 (10 %) | 4 (3 %) | 0.0074 |

| | | | | |
|---|---|---|---|---|
| Grade 3 joint pain | 1 (< 1 %) | 0 | 0 | 1.0 |
| Oropharyngeal pain | 22 (9 %) | 7 (5 %) | 6 (5 %) | 0.27 |
| Cough | 12 (5 %) | 2 (2 %) | 3 (2 %) | 0.24 |
| Nausea | 20 (8 %) | 6 (5 %) | 4 (3 %) | 0.14 |
| Hypersensitivity | 0 | 0 | 2 (2 %) | 0.061 |
| Dyspnoea | 1 (< 1 %) | 0 | 0 | 1.0 |
| Grade 3 dyspnoea | 1 (< 1 %) | 0 | 0 | 1.0 |
| Impaired appetite | 27 (11 %) | 7 (5 %) | 3 (2 %) | 0.0089 |
| Syncope | 1 (< 1 %) | 1 (1 %) | 0 | 1.0 |
| Mucosal abnormality | 2 (1 %) | 2 (2 %) | 2 (2 %) | 0.65 |
| Itching | 6 (2 %) | 4 (3 %) | 6 (5 %) | 0.40 |
| **Unsolicited adverse reactions within 14 days** | | | | |
| Any | 19 (8 %) | 7 (5 %) | 7 (6 %) | 0.65 |
| Grade 3 | 1 (< 1 %) | 0 | 0 | 1.0 |
| **Overall adverse events within 28 days** | | | | |
| Any | 196 (77 %) | 98 (76 %) | 61 (48 %) | < 0.0001 |
| Grade 3 | 24 (9 %) | 1 (1 %) | 2 (2 %) | 0.0002 |

Data show the number of participants (%).
Any refers to the participants with adverse events or reactions of any grade.
VP = viral particles.
Adverse events or reactions were graded according to the scale of the National Medical Product Administration of China.
Grade 3 = severe (i.e., prevents activities of daily living).
P-value was generated by comparisons across the three study groups.
*Multiple comparisons between the $10x10^{10}$ VP dose group and $5x10^{10}$ VP dose group, p = 0.0008.
‡Multiple comparisons between the $10x10^{10}$ VP dose group and $5x10^{10}$ VP dose group, p = 0.0038.

#### 3.4 Background Information

##### *3.4.1 Overall Study Design and Plan*

This is a phase III multicentre, randomised, double-blind, placebo-controlled, parallel-group study. The purpose of the study is the evaluation of the efficacy, reactogenicity and safety of the Ad5-nCoV vaccine compared with placebo in adults aged 18 to 85 years. The study will be performed in the Russian Federation and the Republic of Belarus.

The study design and methodology have been developed in line with the FDA [7], EMA [8] and EAEU [9] guidelines, as well as the regulatory documents of the Russian Federation [10, 11] and the Republic of Belarus [12, 13, 14].

As far as there are no licenced vaccines against novel coronavirus infection, the use of placebo as the study drug is highly justified.

The primary objective of the study is to prove the immunological superiority of the Ad5-nCoV vaccine compared to placebo. This study design is determined by the extreme urgency of the availability of vaccines to prevent novel coronavirus infection.

Efficacy and immunogenicity endpoints used in this study (seroconversion rate, geometric mean titer and geometric mean fold-rise in titers of serum RBD-specific antibodies, antibodies against the S protein of SARS-CoV-2 and neutralising antibodies against SARS-CoV-2, geometric mean titer and geometric mean fold-rise in titers of Ad5 neutralising antibodies and cellular immune response) are common for vaccine immunogenicity studies. Immunogenicity assessment time points (Day 0, Day 14, Day 28, Month 6) are consistent with the objectives of the study and chosen based on the previous clinical studies of Ad5-nCoV.

Protective efficacy assessment (investigation of COVID-19 morbidity) is an exploratory objective of this study due to the limited sample size.

##### *3.4.2 Study Population*

The study will include male and female volunteers aged from 18 to 85 years with no history of COVID-19, with negative tests for SARS-CoV-2 (serum IgM and IgG antibodies, virus RNA [nose and throat swab tests]), with no signs of acute diseases and clinically significant abnormalities at screening and without diseases and/or conditions possibly affecting the safety and the assessment of the study results. Considering data obtained in previous clinical trials of Ad5-nCoV (section 3.3.2), including safety data of subjects older than 55 years, the study population is highly justified and appropriate for the investigation of safety, reactogenicity and efficacy of the vaccine against novel coronavirus infection in adults in the EAEU.

##### *3.4.3 Justification for the Route of Administration, Dosage and Dosing Regimen of the Investigational Vaccine*

The investigational vaccine (Ad5-nCoV) will be administered intramuscularly in the deltoid muscle of the upper arm as a single dose of $5x10^{10}$ VP/0.5 mL (1 pre-filled syringe).

The route of administration, dosage and dosing regimen are chosen based on the results of pre-clinical and clinical studies of Ad5-nCoV (section 3.3).

The results of the dose-escalation phase I and II clinical trials demonstrate that the $5x10^{10}$ VP dose can induce a strong immune response and has a favourable safety profile.

#### 3.5 Summary of Known and Potential Risks and Benefits for Study Subjects

In this study, potential health benefit could be obtained by volunteers randomised in the Ad5-nCoV group (i.e. they will be vaccinated against novel coronavirus infection and undergo medical examinations free of charge). Subjects in the placebo group will get additional information about their health based on the results of medical examinations. Medical examinations will be performed during the study free of charge.

For ethical reasons, to ensure efficient vaccination of the maximum number of volunteers, the subjects will be randomised in the Ad5-nCoV and placebo group in a 3:1 ratio (Ad5-nCoV:placebo).

Risks related to the participation in this study include risks associated with the administration of the investigational vaccine, study procedures and experimental nature of the study.

*Risks Associated with the Administration of the Investigational Vaccine*

Risks associated with the administration of the investigational vaccine relate to possible adverse reactions, including systemic and local post-vaccination reactions and complications.

In a phase I trial, post-vaccination reactions to the vaccine within the first 7 days were registered in 30 of 36 subjects received a single dose of $5x10^{10}$ VP/0.5 mL. Moreover, severe adverse reactions (grade 3) were registered only in two participants (fever in both cases). Local reactions to the vaccine administration included pain (47 %), swelling (11 %), induration (6 %), redness (6 %) and itching (6 %) at the injection site. Systemic reactions included fatigue (47 %), body temperature increase (42 %), headache (39 %), myalgia (19 %), appetite impairment (17 %), diarrhoea (8 %), arthralgia (6 %), nausea (6 %), sore throat (3 %), cough (3 %), functional GI disorders (3 %), vomiting (3 %), dizziness (3 %) and pruritus (3 %). Overall, during the first 28 days after vaccination, adverse reactions were registered in 31 (86 %) of 36 participants in the $5x10^{10}$ VP/0.5 mL dose group [5].

In a phase II trial, post-vaccination reactions to the vaccine within the first 14 days were registered in 96 (74 %) of 129 subjects received a single dose of $5x10^{10}$ VP/0.5 mL. Moreover, a severe adverse reaction was registered only in one participant (fever).

Local reactions to the vaccine administration included pain (56 %), swelling (4 %), induration (2 %), itching (2 %) and redness (1 %). Systemic reactions included fatigue (34 %), headache (28 %), myalgia (18 %), body temperature increase (16 %), arthralgia (10 %), diarrhoea (8 %), nausea (5 %), appetite impairment (5 %), oropharyngeal pain (5 %), pruritus (3 %), cough (2 %), mucosal abnormality (2 %), vomiting (1 %) and syncope (1 %).

Adenovirus vector used in Ad5-nCoV is a highly effective and well-known platform for vaccine antigen delivery. However, there are data on the possible increase of sensitivity to the HIV infection after using adenovirus vector-based vaccines, which can relate to the activation of CD4+ T cells by Ad5 [3]. The potential correlation between the increased sensitivity to the HIV and the administration of adenovirus vector-based vaccines is a subject of debate, and its mechanism is not clear, but these potential risks should be considered during the investigation of adenovirus vector-based vaccines. Due to this, a repeated HIV test is performed at the end of the follow-up period during clinical trials of Ad5-nCoV, including this study.

Safety measures taken during the first hours and days after vaccination in this trial will include the monitoring of the volunteer's condition by a study doctor for 2 hours and a phone call in 5–8 hours after vaccination, as well as visits to the trial site for examination in 48–72 hours and 7 days after immunisation. Further safety evaluation will be performed by questioning and examining the volunteer during visits in 14, 28 days and 6 months after vaccination. Collection of information about adverse events will be performed during phone calls in 2, 3, 4 and 5 months after immunisation.

Thus, the total period of safety evaluation will include at least 6 months after the administration of the vaccine. The duration of the follow-up after vaccination, the list of procedures performed to evaluate the reactogenicity and safety of the vaccine, as well as time points and frequency of such procedures comply with the guidelines for vaccine safety assessment.

In the case of any significant reaction, study participants may come to an unscheduled visit to the trial site to receive necessary medical care and follow-up supervision.

*Risks Related to Study Procedures*

Risks related to study procedures are associated with blood sampling from the vein for immunologic tests and safety laboratory analyses. The total volume of blood withdrawn during the trial per subject will be around 145 mL. During blood sampling, participants may experience dizziness, weakness, lightheadedness, pain at the puncture site, haemorrhages, the formation of thrombi and forearm vein inflammation. The procedure will be performed by qualified personnel of the trial site using accepted aseptic techniques. In the case of adverse reactions, volunteers will be immediately provided with medical help.

During an ECG recording, subjects may feel some discomfort at the site of electrode attachment. In some cases, bruises may occur after placement of sensors on the chest (if suction cups are used).
Allergic skin reactions (e.g. rash and/or burning sensation) to conductive gel or adhesive (if sticky pads are used) can also be seen in rare cases.

Other risks accepted by the study subjects are determined by an experimental nature of the trial as its exact result is unknown.

#### 3.6 Legal Basis

This clinical trial will be conducted in accordance with the requirements of the Good Clinical Practice of the International Conference on Harmonisation of Technical Requirements for Registration of Pharmaceuticals for Human Use (ICH GCP), the ethical principles of the most recent version of the World Medical Association Declaration of Helsinki, and the national regulatory requirements.

### 4 STUDY PURPOSE AND OBJECTIVES

#### 4.1 Study Purpose

The purpose of the study is the evaluation of the efficacy, reactogenicity and safety of the Ad5-nCoV vaccine compared with placebo in adults aged 18 to 85 years.

#### 4.2 Study Objectives

##### *4.2.1 Primary Objective*

The primary objective of the study is to prove the superiority of the Ad5-nCoV vaccine compared with placebo in terms of seroconversion (proportion of subjects with at least four-times increase in antibody titers against the receptor-binding domain [RBD] of the SARS-CoV-2 S protein) on Day 28 after vaccination.

##### *4.2.2 Secondary Objectives*

1. To evaluate the immunogenicity of the Ad5-nCoV vaccine compared with placebo based on the following:

- Geometric mean titer of serum antibodies against the RBD and SARS-CoV-2 S protein on Day 14, 28 and 6 months after vaccination.
- Seroconversion rate (proportion of subjects with at least four-times increase in antibody titers against the RBD and SARS-CoV-2 S protein) on Day 14, 28 (S protein only) and 6 months after vaccination.
- Geometric mean fold-rise in titers of serum antibodies against the RBD and SARS-CoV-2 S protein on Day 14, 28 and 6 months after vaccination.
- Geometric mean titer of neutralising antibodies against SARS-CoV-2 on Day 14, 28 and 6 months after vaccination.
- Seroconversion rate (proportion of subjects with at least four-times increase in neutralising antibody titers against SARS-CoV-2) on Day 14, 28 and 6 months after vaccination.
- Geometric mean fold-rise in titers of neutralising antibodies against SARS-CoV-2 on Day 14, 28 and 6 months after vaccination.
- Geometric mean titer of neutralising antibodies against the Ad5 vector on Day 28 and 6 months after vaccination.
- Geometric mean fold-rise in titers of neutralising antibodies against the Ad5 vector on Day 28 and 6 months after vaccination.
- Cellular immune response (the number of IFNy-secreting T cells [ELISpot]; percentage of CD4+ and CD8+ T cells expressing IFNy, TNF and IL-2 [flow cytometry]) on Day 14, 28 and 6 months after vaccination.

2. To evaluate the efficacy of the Ad5-nCoV vaccine (exploratory analysis) based on the following:

- Frequency of confirmed COVID-19 cases during 6 months after vaccination (except for the cases occurred during the first 14 days after vaccination).

  *A confirmed COVID-19 case means the presence of clinical manifestations and a positive laboratory test result for SARS-CoV-2 RNA.*
- Frequency of confirmed COVID-19 cases requiring hospitalisation (except for the cases occurred during the first 14 days after vaccination).
- Frequency of severe COVID-19 cases (except for the cases that occurred during the first 14 days after vaccination).

- Frequency of lethal COVID-19 cases (except for the cases that occurred during the first 14 days after vaccination).

3. To evaluate the reactogenicity of the Ad5-nCoV vaccine compared with placebo.
4. To evaluate the safety of the Ad5-nCoV vaccine compared with placebo.

### 5 STUDY DESIGN

#### 5.1 Study Design Description

This is a phase III multicentre, randomised, double-blind, placebo-controlled, parallel-group study.

The study will be conducted in the Russian Federation and the Republic of Belarus at about 10 trial sites. Accrual will be competitive.

The purpose of the study is the evaluation of the efficacy, reactogenicity and safety of the Ad5-nCoV vaccine compared with placebo in adults aged 18 to 85 years.

Five hundred (500) volunteers will be randomised in two treatment (vaccination) groups in a 3:1 ratio (Ad5-nCoV:Placebo) as follows:

- Group 1: a single dose of the Ad5-nCoV vaccine.
- Group 2: a single dose of placebo.

The study design includes 7 outpatient visits to trial sites and 5 phone calls with the study doctor:

- Screening visit (Day −10...Day −1).
- Visit 1 (Day 0) — randomisation, a single administration of the investigational product, volunteer's stay at the trial site for 2 hours after vaccination, a phone call with the study doctor in 5–8 hours after vaccination.
- Visit 2 (Day 2) — reactogenicity and safety evaluation in 48–72 hours after vaccination.
- Visit 3 (Day 7) — reactogenicity and safety evaluation in 7 days after vaccination.
- Visit 4 (Day 14) — safety and immunogenicity evaluation in 14 days after vaccination.
- Visit 5 (Day 28) — safety and immunogenicity evaluation in 28 days after vaccination.
- Phone calls with the study doctor in 2, 3, 4 and 5 months after vaccination (safety evaluation).
- Visit 6 (Month 6) — safety and immunogenicity evaluation in 6 months after vaccination.

During each next visit and phone call after vaccination, subjects will be interviewed regarding the signs of acute respiratory viral infection to detect and confirm a COVID-19 case (exploratory protective efficacy evaluation).

The investigation of the cellular immune response will be performed in a separate cohort of at least 60 subjects randomised at trial sites in Moscow.

The study flowchart is shown in Figure 9.

Schedule of procedures is presented in Table 14. A detailed description of the study plan is presented in section 6.

One interim and one final analysis are planned for the study. The unblinded interim analysis will be performed after obtaining efficacy, reactogenicity and safety data through Visit 5 (Day 28) for the first 200 volunteers randomised in the study. Based on the results of this analysis, an interim clinical trial report will be prepared, which will be submitted to regulatory authorities to make a decision on the registration of the Ad5-nCoV vaccine.

The final clinical trial report will be prepared after obtaining all data for all volunteers randomised in the study, i.e. after the completion of the final visit by all the volunteers in 6 months after vaccination. The final clinical trial report will be also submitted to regulatory authorities.

**Screening** **Vaccination** **Follow-up period**

| **Ad5-nCoV** |
|---|
| **Placebo** |

**Randomisation**
**Single vaccina-tion**

| Day −10–−1 | Day 0 | Day 2 | Day 7 | Day 14 | Day 28 | Month 2, 3, 4, 5 | Month 6 |
|---|---|---|---|---|---|---|---|
| **Visit 0** | **Visit 1** | **Visit 2** | **Visit 3** | **Visit 4** | **Visit 5** | **Phone call** | **Visit 6** |

**Reactogenicity evaluation**

**Efficacy and safety evaluation**

**Figure 9 Study Flowchart**

*Visit 1 includes volunteer's stay at the trial site for 2 hours after vaccination and phone call with the study doctor in 5–8 hours after vaccination

#### 5.2 Measures Taken to Minimise/Avoid Bias

To minimise bias, randomisation and blinding will be used for the study.

##### *5.2.1 Randomisation*

At Visit 1 (Day 0), screened and eligible volunteers will be randomly allocated into the Ad5-nCoV group or the placebo group in a 3:1 ratio (Ad5-nCoV:placebo).

The randomisation plan with randomisation numbers for the subjects will be provided by an independent statistician using a validated system including a pseudorandom number generator with a seed value. Thus, the resulted randomisation sequence with numbers and prescribed products will be both reproducible and non-predictable. Block randomisation and stratification by the trial site will be used in this study.

Centralised randomisation of subjects in the study will be performed using the Interactive Web Response System (IWRS). To get familiar with the IWRS, the investigators will be trained to work with the system and receive the IWRS User Guide. A username and password are used to access the system. They are sent to the authorised employee of the trial site at the beginning of the study.
As a result of the randomisation procedure, the volunteer will be assigned a randomisation number. Randomisation codes will be kept by authorised personnel of the CRO responsible for the organisation and implementation of randomisation in this study.

##### *5.2.2 Blinding*

This study will be conducted according to a double-blind design, so neither the study doctor nor study participants will be aware of the vaccine (Ad5- nCoV or placebo) administered to the subjects.

The appearance of Ad5-nCoV and placebo vials and their packaging will be identical. Labelling of the products will be performed to maintain the blinding (masking).

##### *5.2.3 Emergency Unblinding*

Unblinding during the study is only possible in the case of emergency, e.g. in the case of a serious and/or unexpected adverse event related to the use of the investigational vaccines as judged by the investigator, if the treatment of such adverse event is not possible without the information about the administered product.

Information about the product assigned to the volunteer will be available to the investigator via the IWRS, and the unblinding procedure will be described in the IWRS User Guide.

All cases of emergency unblinding during the study must be fully documented.

#### 5.3 Efficacy and Safety Endpoints

##### *5.3.1 Efficacy Endpoints*

###### *5.3.1.1 Primary Efficacy Endpoint*

Seroconversion rate (proportion of subjects with at least four-times increase in antibody titers against the receptor-binding domain [RBD] of the SARS-CoV-2 S protein) on Day 28 after vaccination.

###### *5.3.1.2 Secondary Efficacy Endpoints*

- Geometric mean titer of serum antibodies against the RBD and SARS-CoV-2 S protein on Day 14, 28 and 6 months after vaccination.
- Seroconversion rate (proportion of subjects with at least four-times increase in antibody titers against the RBD and SARS-CoV-2 S protein) on Day 14, 28 (S protein only) and 6 months after vaccination.
- Geometric mean fold-rise in titers of serum antibodies against the RBD and SARS-CoV-2 S protein on Day 14, 28 and 6 months after vaccination.
- Geometric mean titer of neutralising antibodies against SARS-CoV-2 on Day 14, 28 and 6 months after vaccination.
- Seroconversion rate (proportion of subjects with at least four-times increase in neutralising antibody titers against SARS-CoV-2) on Day 14, 28 and 6 months after vaccination.
- Geometric mean fold-rise in titers of neutralising antibodies against SARS-CoV-2 on Day 14, 28 and 6 months after vaccination.
- Geometric mean titer of neutralising antibodies against the Ad5 vector on Day 28 and 6 months after vaccination.
- Geometric mean fold-rise in titers of neutralising antibodies against the Ad5 vector on Day 28 and 6 months after vaccination.
- Cellular immune response (the number of IFNy-secreting T cells [ELISpot]; percentage of CD4+ and CD8+ T cells expressing IFNy, TNF and IL-2 [flow cytometry]) on Day 14, 28 and 6 months after vaccination.

###### *5.3.1.3 Exploratory Efficacy Endpoints*

- Frequency of confirmed COVID-19 cases during 6 months after vaccination (except for the cases occurred during the first 14 days after vaccination).

  *A confirmed COVID-19 case means the presence of clinical manifestations and a positive laboratory test result for SARS-CoV-2 RNA.*

- Frequency of confirmed COVID-19 cases requiring hospitalisation (except for the cases occurred during the first 14 days after vaccination).
- Frequency of severe COVID-19 cases (except for the cases that occurred during the first 14 days after vaccination).
- Frequency of lethal COVID-19 cases (except for the cases that occurred during the first 14 days after vaccination).

##### *5.3.2 Safety Endpoints*

- Reactogenicity (frequency and nature of systemic and local immunisation reactions on the day of vaccination and within 7 days after vaccination).
- Frequency and nature of adverse events (Day 0–Day 28) and serious adverse events (Day 0–Day 28; Day 0–Month 6).
- Vital signs.
- Physical examination results.
- Electrocardiography.

- Biochemistry.
- Haematology.
- Coagulation test.
- Urinalysis.
- Serum immunoglobulin E concentration.

#### 5.4 Selection and Withdrawal of Subjects

##### *5.4.1 Selection of Subjects*

A total of 500 subjects aged 18–85 years old will be randomised into the study.

Volunteers will be invited to participate in the trial specifically through advertising. All advertising materials for potential subjects will be initially provided to the ethics committee for approval.

##### *5.4.2 Inclusion Criteria*

Subjects must meet all of the following criteria to be eligible for participation in the study:

1. Signed and dated Informed Consent Form for participation in the study.
2. Men and women aged 18–85 years.
3. Body mass index 18.5–30.0 kg/m$^2$.
4. Negative SARS-CoV-2 RNA PCR test at screening.
5. Negative SARS-CoV-2 IgM and IgG antibody test at screening.
6. No history of COVID-19.
7. No close contacts with persons suspected for SARS-CoV-2 infection or persons with laboratory-confirmed COVID-19 within the last 14 days.
8. No signs of respiratory infection within the last 14 days.
9. Negative tests for human immunodeficiency virus (HIV), syphilis, HBV and HCV.
10. No history and screening examination findings of diseases and/or conditions that, in the investigator's opinion, may have an impact on the safety of the subject in the study and the assessment of the study results.
11. Subject's consent to use reliable methods of contraception during the entire study period.

##### *5.4.3 Exclusion Criteria*

Subjects cannot be included in the study in the case of meeting any of the following criteria:

1. Positive allergological anamnesis, drug hypersensitivity, including hypersensitivity to any component of the study drug, as well a history of serious adverse events to vaccines (such as allergic reactions, respiratory disturbance, angioedema, abdominal pain).
2. Axillary temperature ≥ 37.1 °C at screening/randomisation.
3. Systolic blood pressure > 139 mm Hg or < 100 mm Hg and/or diastolic blood pressure > 90 mm Hg or < 60 mm Hg.
4. Clinically relevant abnormalities during laboratory and/or instrumental examinations at screening.
5. Acute infectious diseases less than 4 weeks before screening.
6. Acute diseases or exacerbations of chronic diseases of the liver, kidneys, gastrointestinal, cardiovascular, respiratory, nervous or endocrine system.
7. History of moderate and severe asthma or pulmonary fibrosis.
8. History of blood or hematopoietic disorders.
9. History of diabetes mellitus.
10. History of epilepsy, epileptic syndrome or convulsive seizures.
11. Congenital or acquired immunodeficiency, HIV infection, lymphoma, leukaemia, lupus erythematosus, juvenile rheumatoid arthritis or a history of other autoimmune disorders.

12. History of malignant neoplasms.
13. Administration of immunotropic drug products (immunomodulatory agents, immunostimulants, immunosuppressants), allergy medications and cytotoxic drugs for > 10 consecutive days within the last 3 months (except for inhaled and topical glucocorticosteroids [GCs]) or the administration of these drugs less than 4 weeks before screening.
14. Administration of immunoglobulins or transfusion less than 3 months before screening.
15. Administration of antipyretics (including nonsteroidal anti-inflammatory drugs and anilides) within 24 hours before randomisation.
16. Blood donation or blood loss (≥450 mL of blood or plasma) less than 3 months before screening.
17. Vaccination within 6 months before screening or unwillingness to skip any other vaccination during the study.
18. History/data of alcohol and drug use/drug abuse or mental disorders.
19. Major surgery scheduled for the next 6 months or performed within the last 6 months before screening.
20. Piercings, permanent makeup/tattoos made less than 1 month before screening or during the study.
21. Pregnancy or lactation.
22. Participation in another clinical trial within 3 months before screening.
23. Psychological, physical or other reasons not allowing the subject to comply with conditions and procedures of the Protocol.
24. Subjects who are employees of healthcare facilities and in contact with persons diagnosed with COVID-19.
25. Subjects who are employees at the trial site.
26. Other reasons not allowing the subject to participate in the study in the opinion of the study doctor.

##### *5.4.4 Inclusion of Volunteers Incapable of Giving Informed Consent*

It is not intended to include volunteers who are incapable of giving informed consent.

##### *5.4.5 Premature Withdrawal from the Study*

According to the World Medical Association Declaration of Helsinki and other applicable regulatory documents, a volunteer may withdraw from the study at any time and due to any reason.

Study subjects may be prematurely withdrawn from the study in the case of early termination of the study upon the Sponsor's decision (section 5.4.6).

Additionally, study subjects may be prematurely withdrawn from the study due to the following reasons:

1. Subject's request to discontinue for any reason.
2. Development of conditions not meeting the subject's eligibility criteria for the study (the decision to exclude the subject should be made by the investigator and agreed with the Sponsor on an individual basis).
3. Change in health status preventing the subject from further participation in the study as judged by the investigator (e.g. an adverse event or serious adverse event).
4. Lost to contact.
5. Violation of the trial protocol.
6. Pregnancy of the subject.

Reasons for premature withdrawal of a volunteer from the study must be registered in the source documents and the CRF.

Study subjects will not be replaced. The exception is made for volunteers completed the screening procedures and eligible for the study, but discontinued from the study before randomisation and administration of the study drug due to any reason.

A subject discontinued from the trial during the screening period before randomisation (e. g. due to ineligibility or informed consent withdrawal) should be classified as a screening failure.

Subjects discontinued after randomisation should be classified as dropouts. Data of all early withdrawn subjects will be included in the study database and final analysis. For volunteers withdrawn before the follow-up period, procedures expected at Visit 5 (Day 28) will be performed at the End of Study Visit. For volunteers withdrawn during the follow-up period, procedures expected at Visit 6 (Month 6) will be performed (section 6.4.2).

###### *5.4.6 Early Termination of the Study*

The Sponsor or local regulatory authorities may suspend or terminate the study at any time due to medicinal and/or administrative reasons.

In the case of early termination of the study regardless of the reason, the study doctor must immediately inform all volunteers about it and ensure their follow-up. If the study is suspended or terminated upon the Sponsor's decision, the sponsor must inform the investigator, regulatory authorities and ethic committee about the study termination in writing specifying the reason for such decision.

In the case of early termination of the study, the investigator should (whenever possible) perform the study procedures and examinations described in section 5.4.5 for all volunteers on the study at that time.

##### 5.5 Study Restrictions

*Restrictions in Concomitant Treatment*

Information about restrictions regarding concomitant therapy is provided in section 7.7.

*Contraception*

When signing the Informed Consent Form, all women of childbearing potential and men whose partners are women of childbearing potential should agree to use proper contraception methods during the study from the moment of signing of the Informed Consent Form and until the end of the follow-up period.

Requirements for registration and reporting of pregnancy cases are provided in section 8.4.2.8.9.

The following subjects are considered women of non-childbearing potential:

- postmenopausal women, i.e. women who had no menstruations for 12 consecutive months without any alternative medical reason
- women who underwent sterilisation (for example, bilateral tubal ligation more than one menstrual period before the enrolment, hysterectomy or bilateral oophorectomy).

Proper (reliable) contraception methods include:

1. Complete abstinence [periodic abstinence (for example, rhythm or symptothermal methods) is not a proper contraception method].
2. Male sterilisation (with the corresponding post-vasectomy confirmation of the absence of sperm in the ejaculate).
3. Intrauterine device or intrauterine system.
4. Hormonal methods of contraception: oral, injectable or implantable hormonal contraceptives. Women who use hormonal methods of contraception should use the specific method for at least 6 months before the administration of the study drug and should continue to use such method during the entire study until the end of the follow-up period.
5. Condom in combination with a spermicide (foam/gel/cream/film/suppositories).
6. Occlusive cap (diaphragm, cervical or vault caps) with a spermicide (foam/gel/cream/film/suppositories).
7. Double-barrier method: condom and occlusive cap (diaphragm, cervical or vault caps) with a spermicide (foam/gel/cream/film/suppositories).

*Other Restrictions*

On days of blood sampling for laboratory assessments, volunteers should come to the visit fasted.

During the first 14 days after vaccination, the volunteers should observe the following precautions against novel coronavirus infection as strictly as possible:

- Avoid travelling to other regions, using public transport and visiting crowded places (except for visits to the trial site).
- Avoid close contacts with persons with acute respiratory infection symptoms.
- Keep a 1.5-meter distance from others in public places, use disposable medical masks and change them every 2 hours.
- Practice personal hygiene (wash hands with soap, touch the face with clean napkins or washed hands only).
- Use personal protective equipment (for health-care workers).

After that, the subjects should observe preventive measures recommended in their region.

### 6 STUDY METHODOLOGY

#### 6.1 Schedule of Visits and Procedures

The study design includes 7 outpatient visits to trial sites and 5 phone calls with the study doctor:

- Visit 0 (screening) — Day −10...Day −1
- Visit 1 (randomisation, vaccination), phone contact in 5–8 hours — Day 0
- Visit 2 — Day 2
- Visit 3 — Day 7
- Visit 4 — Day 14
- Visit 5 — Day 28
- Follow-up period/Phone calls (Month 2, 3, 4, 5)
- Follow-up period/Visit 6 — Month 6

The total duration of the subject's participation in the study will be about 6.5 months (about 205 days). The study flowchart is shown in Figure 9 (section 5.1).

The schedule of visits and procedures is presented in Table 14 below.

The investigation of the cellular immune response will be performed in a separate cohort of at least 60 subjects randomised at trial sites in Moscow.

**Table 14 Schedule of Visits and Procedures**

| **Study visit** | **Visit 0 (V0)** | **Visit 1 (V1)** | **Visit 2 (V2)** | **Visit 3 (V3)** | **Visit 4 (V4)** | **Visit 5 (V5)** | **Phone contact** | **Visit 6 (V6)** |
|---|---|---|---|---|---|---|---|---|
| **Study day/month** | **Day −10–−1 (screening)** | **Day 0[1] (vaccination)** | **Day 2 [V1 + 2 days]** | **Day 7 [V1 + 7 days]** | **Day 14 [V1 + 14 days]** | **Day 28 [V1 + 28 days]** | **Month 2 (V5 + 1 month)<br>Month 3 (V5 + 2 months)<br>Month 4 (V5 + 3 months)<br>Month 5 (V5 + 4 months)** | **Month 6 (V5 + 5 months)** |
| **Visit window** | **-** | **-** | **+ 1 day[2]** | **± 1 day** | **± 1 day** | **± 1 day** | **± 5 days** | **± 15 days** |
| Informed consent | • | | | | | | | |
| Demographics/medical history[3] | • | | | | | | | |
| Prior treatments | • | • | | | | | | |
| Analysis for IgM and IgG antibodies to SARS-COV-2[4] | • | | | | | • | | |
| Analysis of SARS-COV-2 RNA (PCR) | • | *If COVID-19 is suspected, a nose and throat swab will be taken at home twice with the interval of 3 days* | | | | | | |
| Height, body weight, BMI | • | | | | | | | |
| Physical examination, including neurologic examination[5] | • | • | • | • | • | • | | • |

[1]Within 2 hours after vaccination, the volunteers must be observed at the trial site for systemic and local reactions to the vaccine administration, including immediate hypersensitivity. In 20 minutes (± 5 minutes) and 2 hours (± 10 minutes) after the vaccine administration, the examination of the injection site, body temperature and vital signs measurement, and AEs and concomitant treatment registration will be performed. In 2 hours (± 10 minutes), an additional limited physical examination will be performed. In 5–8 hours after vaccination, the study doctor will contact the volunteer by phone to collect data on AEs and concomitant treatment.
[2]Visit 2 is performed in 48–72 hours after vaccination.
[3]During the collection of medical history at screening, it is necessary to ask the volunteer about the signs and symptoms of COVID-19 (section 8.3) and obtain epidemiological anamnesis (regarding close contacts with persons suspected for SARS-CoV-2 infection or persons with laboratory-confirmed COVID-19 within the last 14 days).
[4]Tests for IgM and IgG antibodies to the N protein and IgG antibodies to the S protein of SARS-CoV-2 at screening and tests for IgM and IgG antibodies to the N protein of SARS-CoV-2 at Visit 5 (Day 28).
[5]Comprehensive physical examination is performed only at screening. At other visits, a limited examination is performed (general appearance, ears, nose, throat, skin and injection site, lymph nodes, cardiovascular system, respiratory system, and neurological system).

| **Study visit** | **Visit 0 (V0)** | **Visit 1 (V1)** | **Visit 2 (V2)** | **Visit 3 (V3)** | **Visit 4 (V4)** | **Visit 5 (V5)** | **Phone contact** | **Visit 6 (V6)** |
|---|---|---|---|---|---|---|---|---|
| **Study day/month** | **Day −10–−1 (screening)** | **Day 0[1] (vaccination)** | **Day 2 [V1 + 2 days]** | **Day 7 [V1 + 7 days]** | **Day 14 [V1 + 14 days]** | **Day 28 [V1 + 28 days]** | **Month 2 (V5 + 1 month) Month 3 (V5 + 2 months) Month 4 (V5 + 3 months) Month 5 (V5 + 4 months)** | **Month 6 (V5 + 5 months)** |
| Vital signs [6] | • | • | • | • | • | • | | • |
| Body temperature[7] | • | • | • | • | • | • | | • |
| Electrocardiography | •[8] | | • | | | | | |
| Infections[9] | • | | | | | | | • |
| Haematology[10], biochemistry[11], coagulation test[12], urinalysis[13] | • | | • | | | • | | |
| Blood IgE test | • | | | | | • | | |
| Urine pregnancy test[14] | • | | | | | | | |
| Evaluation of inclusion/exclusion criteria | • | • | | | | | | |
| Randomisation | | • | | | | | | |
| Vaccination | | • | | | | | | |

[6]Arterial pressure, heart rate, breathing rate.
[7]Additionally, the volunteers should measure the body temperature at home by themselves within 7 days after vaccination (in 5–8 hours after the vaccine administration on the day of immunisation and then twice daily (morning and evening) until Visit 3 (Day 7)). During trial site visits, the volunteers should tell the study doctor about all increases in body temperature.
[8]ECG results obtained not more than 30 days before screening are also acceptable.
[9]Human immunodeficiency virus (HIV), syphilis, HBV and HCV. At Visit 6, only HIV test is performed.
[10]Haematology: haemoglobin, haematocrit, RBC, platelets, WBC, WBC differential and ESR.
[11]Biochemistry: total protein, ALT, AST, ALP, LDH, total bilirubin, creatinine, urea, fasting glucose, C-reactive protein.
[12]Coagulation test: aPTT, PT, fibrinogen.
[13]Urinalysis: specific gravity, pH, protein, glucose, erythrocytes, leucocytes, casts.
[14]For women of childbearing potential.

| **Study visit** | **Visit 0 (V0)** | **Visit 1 (V1)** | **Visit 2 (V2)** | **Visit 3 (V3)** | **Visit 4 (V4)** | **Visit 5 (V5)** | **Phone contact** | **Visit 6 (V6)** |
|---|---|---|---|---|---|---|---|---|
| **Study day/month** | **Day −10–−1 (screening)** | **Day 0[1] (vaccination)** | **Day 2 [V1 + 2 days]** | **Day 7 [V1 + 7 days]** | **Day 14 [V1 + 14 days]** | **Day 28 [V1 + 28 days]** | **Month 2 (V5 + 1 month)<br>Month 3 (V5 + 2 months)<br>Month 4 (V5 + 3 months)<br>Month 5 (V5 + 4 months)** | **Month 6 (V5 + 5 months)** |
| Reactogenicity evaluation[15] | | • | • | • | | | | |
| Blood tests for the S protein and RBD of SARS-CoV-2, neutralising antibodies against SARS-CoV-2, cellular immune response assessments[16] | | • | | | • | • | | • |
| Blood test for neutralising antibodies against Ad5 | | • | | | | • | | • |
| Blood sampling for immune response exploratory analyses | | • | | | • | • | | • |
| Assessment of the signs of acute respiratory infection[17] | | | • | • | • | • | • | • |
| Registration of concomitant treatment[18] | | • | • | • | • | • | • | • |
| Registration of adverse events[19] | | • | • | • | • | • | • | • |

[15]Assessment of local and systemic reactions to the vaccine administration by the study doctor within 7 days after vaccination, including in 20 minutes (± 5 minutes), 2 hours (± 10 minutes) and 5–8 hours after immunisation on Day 0. Reactogenicity assessment is performed during the registration of adverse events.
[16]The investigation of the cellular immune response will be performed in a separate cohort of at least 60 subjects randomised at trial sites in Moscow.
[17]During each visit/phone contact after the day of vaccination, the signs of acute respiratory infection are assessed. In the case of any signs and symptoms related to COVID-19 after immunisation, it is necessary to follow the procedures described in section 8.3.
[18]Visit 1–5: all concomitant treatments. Visit 5–6: only treatments related to SAEs.
[19]Visit 0–5: all AEs. Visit 5–6: only SAEs.

#### 6.2 Details on Study Periods and Visits

##### *6.2.1 Screening/Visit 0 (Day −10...−1)*

Screening will be performed in an outpatient setting during not more than 10 days. If necessary, the screening procedures may be performed within several days or on Day −1 before randomisation. Nevertheless, the eligibility of the subject should be confirmed after the completion of all tests and procedures and after the assessment of all the examination results.

At screening before any protocol required procedures, the volunteers will be invited to participate in this clinical trial and provided with the Subject Information Sheet and Informed Consent Form for review and making a decision on the participation.

The details about the informed consent procedure are provided in section 11.2.

After signing the Informed Consent Form, an individual identification number will be assigned to the volunteer by the investigator. In the case of early withdrawal from the study, the individual identification number will be reserved for the subject and will not be assigned to another volunteer. After receiving a written informed consent and assigning the number, the study doctor will complete and provide the volunteer with an original insurance policy for the period of the study (section 11.4).

The volunteer will also receive an emergency contact card. This card specifies that the volunteer is participating in this clinical trial and contains information about an emergency contact person.

During the screening period, the following procedures will be performed to assess the subject's eligibility[1]:

- obtaining written informed consent for participation in the study
- collection of demographic data (date of birth, sex, race, substance abuse)
- collection of medical history (prior and concomitant diseases) including prior or current signs and symptoms of COVID-19 (section 8.3) and epidemiological anamnesis (regarding close contacts with persons suspected for SARS-CoV-2 infection or persons with laboratory-confirmed COVID-19 within the last 14 days)
- assessment of prior treatments (including previous vaccinations)
- measurement of height, weight and body mass index (BMI)
- comprehensive physical examination
- measurement of body temperature (axillary temperature)
- recording of vital signs
- analysis for SARS-COV-2 RNA (PCR)
- analysis for IgM and IgG antibodies to SARS-COV-2
- analysis for infections (HIV, syphilis, HBV, HCV)
- haematology
- biochemistry
- coagulation test
- urinalysis
- urine pregnancy test (for women of childbearing potential)
- blood IgE test
- ECG (results obtained not more than 30 days before screening are also acceptable)
- evaluation of inclusion/exclusion criteria.

At the end of the screening period, the study doctor should evaluate the examination results to confirm the subject's eligibility.

##### *6.2.2 Vaccination/Visit 1 (Day 0)*

Successfully screened and eligible subjects will come to the trial site for vaccination.

The following procedures will be performed during the visit:

- assessment of prior treatments
- limited physical examination, including the examination of the injection site
- measurement of body temperature

[1] Hereinafter, the list of procedures is presented in the recommended order.

- recording of vital signs
- evaluation of inclusion/exclusion criteria
- randomisation
- blood tests for immunogenicity (antibodies against the S protein and RBD of SARS-CoV-2, neutralising antibodies against SARS-CoV-2, neutralising antibodies against Ad5, cellular immune response assessment [separate cohort[1]]), blood sampling for immune response exploratory analyses
- vaccination.

Within 2 hours after vaccination, the volunteers must be observed at the trial site for systemic and local reactions to the vaccine administration, including immediate hypersensitivity.

In 20 minutes (± 5 minutes) after the vaccine administration, the following procedures will be performed:

- examination of the injection site
- measurement of body temperature
- recording of vital signs
- assessment of systemic and local reactions to the vaccine administration (section 8.4.2.1)
- assessment and registration of adverse events
- registration of concomitant treatments.

In 2 hours (± 10 minutes) after the vaccine administration, the following procedures will be performed:

- limited physical examination, including the examination of the injection site
- measurement of body temperature
- recording of vital signs
- assessment of systemic and local reactions to the vaccine administration (section 8.4.2.1)
- assessment and registration of adverse events
- registration of concomitant treatments.

In 5–8 hours after the vaccine administration, the study doctor will contact the volunteer by phone to collect and register the following:

- data on systemic and local reactions to the vaccine administration (section 8.4.2.1)
- data on adverse events
- data on concomitant treatments.

##### *6.2.3 Visit 2 (Day 2)*

At the visit in 48–72 hours after vaccination, the following procedures will be performed:

- limited physical examination, including the examination of the injection site
- measurement of body temperature
- recording of vital signs
- assessment of systemic and local reactions to the vaccine administration (section 8.4.2.1)
- ECG
- haematology
- biochemistry
- coagulation test
- urinalysis
- assessment of the signs and symptoms of acute respiratory infection (in the case of any signs and symptoms related to COVID-19 after immunisation, it is necessary to follow the procedures described in section 8.3)
- assessment of adverse events
- registration of concomitant treatments.

##### *6.2.4 Visit 3 (Day 7)*

The following procedures will be performed during this visit:

- limited physical examination, including the examination of the injection site

[1]The investigation of the cellular immune response will be performed in a separate cohort of at least 60 subjects randomised at trial sites in Moscow.

- measurement of body temperature
- assessment of systemic and local reactions to the vaccine administration (section 8.4.2.1)
- recording of vital signs
- assessment of the signs and symptoms of acute respiratory infection (in the case of any signs and symptoms related to COVID-19 after immunisation, it is necessary to follow the procedures described in section 8.3)
- assessment of adverse events
- registration of concomitant treatments.

##### *6.2.5 Visit 4 (Day 14)*

The following procedures will be performed during this visit:

- limited physical examination, including the examination of the injection site
- measurement of body temperature
- recording of vital signs
- blood tests for immunogenicity (antibodies against the S protein and RBD of SARS-CoV-2, neutralising antibodies against SARS-CoV-2, cellular immune response assessment [separate cohort[1]])
- blood sampling for immune response exploratory analyses
- assessment of the signs and symptoms of acute respiratory infection (in the case of any signs and symptoms related to COVID-19 after immunisation, it is necessary to follow the procedures described in section 8.3)
- assessment of adverse events
- registration of concomitant treatments.

##### *6.2.6 Visit 5 (Day 28)*

The following procedures will be performed during this visit:

- limited physical examination, including the examination of the injection site
- measurement of body temperature
- recording of vital signs
- blood tests for immunogenicity (antibodies against the S protein and RBD of SARS-CoV-2, neutralising antibodies against SARS-CoV-2, neutralising antibodies against Ad5, cellular immune response assessment [separate cohort[1]])
- analysis for IgM and IgG antibodies to SARS-COV-2
- blood sampling for immune response exploratory analyses
- blood IgE test
- haematology
- biochemistry
- coagulation test
- urinalysis
- assessment of the signs and symptoms of acute respiratory infection (in the case of any signs and symptoms related to COVID-19 after immunisation, it is necessary to follow the procedures described in section 8.3)
- assessment of adverse events
- registration of concomitant treatments.

##### *6.2.7 Follow-up period/Phone calls (Month 2, 3, 4, 5)*

The study doctor will contact the volunteers during Month 2, 3, 4, and 5 to collect the following:

- information on adverse events
- information on concomitant treatments
- information on the signs and symptoms of acute respiratory infection (in the case of any signs and symptoms related to COVID-19 after immunisation, it is necessary to follow the procedures described in section 8.3).

After that, the information about SAEs and concomitant treatments related to the SAEs will be registered

[1]The investigation of the cellular immune response will be performed in a separate cohort of at least 60 subjects randomised at trial sites in Moscow.

in the subject's CRF.

###### *6.2.8 Follow-up Period/Month 6*

The following procedures will be performed during this visit to the trial site:

- limited physical examination, including the examination of the injection site
- measurement of body temperature
- recording of vital signs
- blood tests for immunogenicity (antibodies against the S protein and RBD of SARS-CoV-2, neutralising antibodies against SARS-CoV-2, neutralising antibodies against Ad5, cellular immune response assessment [separate cohort[1]])
- blood sampling for immune response exploratory analyses
- blood HIV test
- assessment of the signs and symptoms of acute respiratory infection (in the case of any signs and symptoms related to COVID-19 after immunisation, it is necessary to follow the procedures described in section 8.3)
- registration of SAEs and concomitant treatments regarding the SAEs.

##### 6.3 Permissible Deviations from the Schedule of Visits and Procedures

Permissible deviations from the schedule of visits (visit windows) are shown in Table 14.

##### 6.4 Unscheduled Visits and Early Withdrawal Visit

###### *6.4.1 Unscheduled Visit*

Volunteers may come to an unscheduled visit to receive medical care.

To register an unscheduled visit, the investigator should indicate the date and reason for such visit in the source documents and CRF together with the results of all procedures and examinations performed during such visit. If the reason for the visit is an adverse event/reaction or if an adverse event/reaction is identified during the unscheduled visit, such adverse event will be registered in an established manner.

###### *6.4.2 End of Study Visit*

Study completion according to the Protocol means the completion of all scheduled visits. Early withdrawal from the study is possible due to reasons specified in sections 5.4.5 and 5.4.6.

In the case of early withdrawal of a randomised and vaccinated volunteer, the investigator should collect as much information as possible about the subject's condition at the time of the withdrawn. For volunteers withdrawn before the follow-up period, procedures expected at Visit 5 (Day 28) will be performed at the End of Study Visit. For volunteers withdrawn during the follow-up period, procedures expected at Visit 6 (Month 6) will be performed.

The feasibility of blood sampling for immunology tests at the Early Withdrawal Visit should be considered by the investigator and the Sponsor on an individual basis.

The results of examinations performed at the End of Study Visit will be included in the source documents and the CRF together with the reason for the volunteer's withdrawal.

##### 6.5 Protocol Deviations

A protocol deviation is any change, divergence, or departure from the study design or procedures described in the clinical trial protocol. The investigator is responsible for reporting protocol deviations upon discovery. The decision to withdraw the volunteer with protocol deviations from the study should be made on an individual basis and agreed with the Sponsor.

Each deviation will be documented on a case-by-case basis and reviewed by the contract research organisation and the Sponsor before the database lock. All protocol deviations will be listed and summarised in the clinical trial report.

[1] The investigation of the cellular immune response will be performed in a separate cohort of at least 60 subjects randomised at trial sites in Moscow.

### 7 STUDY TREATMENTS

#### 7.1 Treatment (Vaccination) of Volunteers

On Day 1 after randomisation and allocation into groups, the volunteers will be vaccinated with test vaccine Ad5-nCoV or placebo.

Before immunisation, each volunteer will be examined, and in the case of contraindications (e.g. axillary body temperature ≥ 37.1 °C), the vaccination will not be performed.

The investigational product (Ad5-nCoV or placebo) will be administered intramuscularly in the deltoid muscle of the upper arm as a single dose of 0.5 mL (1 pre-filled syringe).

The vaccination should be performed in a specially equipped immunisation room only by an authorised employee of the trial site having proper qualification and skill. Emergency antishock medications should be provided in immunisation rooms.

Before the vaccine administration, it is necessary to check the syringe. It is not allowed to administer the product in the case of compromised integrity or labelling, presence of foreign matters, change of physical properties (colour, transparency), expiration and violation of storage conditions.

The vaccination procedure should be performed in strict adherence to the aseptic and antiseptic techniques. The injection site should be swabbed with alcohol or other antiseptics. The skin at the injection site should be completely dry before injection.

Avoid injecting the vaccine into blood vessels, intradermally or subcutaneously.

After the vaccination, the volunteers must stay at the trial site for 2 hours to register any hypersensitivity an immunisation reaction (section 6.2.2).

#### 7.2 Description of Investigational Products, Dosage, Administration and Dosing Regimen

Investigational products (Ad5-nCoV and placebo) will be provided by the Sponsor (NPO Petrovax Pharm LLC, Russia). Description of investigational products is given below.

**Ad5-nCoV Vaccine**

| | |
|---|---|
| Name | Recombinant Novel Coronavirus SARS- Cov-2 Vaccine (Adenovirus Type 5 Vector)<br>Abbreviated name: Ad5-nCoV |
| Formulation | *Active ingredient:* one dose (0.5 mL) contains ≥ $4x10^{10}$ VP (target $5x10^{10}$ VP). |
| Dosage form | *Excipients:* mannitol, sucrose, sodium chloride, magnesium chloride, HEPES (4-(2-hydroxyethyl)-1-piperazineethanesulfonic acid), polysorbate 80 and glycerine.<br>Solution for intramuscular injection |
| How supplied | 0.5 mL (1 dose) in a pre-filled syringe |
| Storage conditions | 2–8 °C |
| Manufacturer | *Manufacturing and primary packaging:*<br>CanSino Biologics Inc., floor 3 and 4, 185 South Ave., TEDA West District, Tianjin, China<br>16 Xinwei Road, TEDA West District, Tianjin, China<br>*Secondary packaging and release quality control:* NPO Petrovax Pharm LLC, 1, Sosnovaya St., Moscow region, Podolsk, Pokrov village 142143, Russia |

**Placebo**

| | |
|---|---|
| Formulation | Mannitol, sucrose, sodium chloride, magnesium chloride, HEPES, polysorbate 80 and glycerine. |
| Dosage form | Solution for intramuscular injection |
| How supplied | 0.5 mL (1 dose) in a pre-filled syringe |
| Storage conditions | 2–8 °C |
| Manufacturer | *Manufacturing and primary packaging:*<br>CanSino Biologics Inc., floor 3 and 4, 185 South Ave., TEDA West District, Tianjin, China<br>16 Xinwei Road, TEDA West District, Tianjin, China<br>*Secondary packaging and release quality control:* NPO Petrovax Pharm LLC, 1, Sosnovaya St., Moscow region, Podolsk, Pokrov village 142143, Russia |

#### 7.3 Dose Selection for Each Subject

All study subjects will be administered with a similar volume (0.5 mL/1 pre-filled syringe) of the investigational products (Ad5-nCoV or placebo).

Justification for dosage and dosing regimen of investigational vaccine Ad5-nCoV ($5x10^{10}$ VP) is provided in section 3.4.3.

#### 7.4 Packaging, Labelling, and Storage Conditions

Packaging and labelling of the investigational products will be carried out in accordance with applicable local regulatory requirements.

The investigational products (Ad5-nCoV and placebo) will be supplied in pre-filled syringes containing 0.5 mL of solution.

The secondary packaging will consist of a carton box containing 1 pre-filled syringe. The secondary packaging will contain a study group code. The label will include the statement “For clinical trial use”.

The investigational products should be stored in the refrigerator with restricted access at the temperature of + 2 to + 8 °C protected from light.

An authorised investigator or pharmacist of the trial site should be responsible for the storage of the products during the study. An authorised employee of the trial site should monitor the temperature in the refrigerator where the products are kept and record the temperature in a temperature log. In the case of deviations from the specified storage conditions, the authorised employee should notify the monitor of the trial site about it. Storage conditions of the product will be reviewed by the monitor during the visits to the trial site.

Shelf life: 2 years. Do not use the products after the expiry of the shelf-life period.

#### 7.5 Product Supply and Accountability

The Sponsor and the authorised CRO will deliver the required amount of the investigational products to the trial site before the beginning of the study. Deliveries to the trial sites will be conducted according to the SOP of the Sponsor or CRO.

An employee responsible for taking the deliveries should check the number of products received and the integrity of the packaging and confirm the receipt in writing. The investigator agrees to distribute the product only among the participants of the clinical trial strictly according to randomisation numbers and use the product in line with the Protocol.

Packages of used products and all unused products should be accounted and kept until the completion of the study. Distribution of the product during the study will be reviewed by the monitor at regular visits to the trial sites.

Unused products and packages of used products will be disposed at the end of the study according to the order of the Sponsor.

#### 7.6 Treatment Compliance

Not applicable. The products will be administered by authorised personnel of the trial sites.

#### 7.7 Concomitant Treatment

**Prohibited Treatment**

- Administration of nonsteroidal anti-inflammatory drugs and anilides, any drugs or food supplements that might increase body temperature (e.g. caffeine), and physical therapy within 24 hours before randomisation.
- Vaccination within 6 months before screening or during the study (except for the investigational product).
- Administration of systemic immunotropic drug products (immunomodulatory agents, immunostimulants, immunosuppressants) and cytotoxic drugs for > 10 consecutive days within 3 months before screening or during the study.
- Administration of allergy medications for > 10 consecutive days within 3 months before screening or during the first 28 days after vaccination (allergy medications are allowed for treatment of adverse events).
- Administration of immunoglobulins or transfusion less than 3 months before screening or during the study.
- Administration of chemoprophylaxis agents (chloroquine, hydroxychloroquine, mefloquine) during the study.

**Allowed Treatment**

The volunteers may receive any drugs prescribed for the treatment of adverse events or concurrent diseases, except for the prohibited products specified above.

### 8 STUDY PROCEDURES AND ENDPOINTS

#### 8.1 Collection of Demographic and Other Baseline Characteristics

##### *8.1.1 Demographics*

The following demographic data will be registered at screening:

- date of birth
- sex
- race
- substance abuse: smoking, drinking, drug use, drug abuse.

##### *8.1.2 Medical History*

During screening, detailed medical history data will be collected, including the information about all current and significant (according to the investigator) prior diseases.

Information about all prior and current diseases at screening or diseases that occur during the interval from the signing of the informed consent form until the administration of the investigational product will be noted in the Medical History section of the subject's Case Report Form (CRF). CRFs should at least include the following information: diagnosis/symptom, onset date and end date (if resolved).

Any untoward medical occurrence in a volunteer administered the investigational product should be registered as an adverse event.

During the collection of medical history at screening, it is necessary to ask the volunteer about the signs and symptoms of COVID-19 (section 8.3) and obtain epidemiological anamnesis (regarding close contacts with persons suspected for SARS-CoV-2 infection or persons with laboratory-confirmed COVID-19 within the last 14 days).

##### *8.1.3 Prior/Concomitant Treatment*

CRFs should include information about the treatment of prior and current diseases, including medications and non-medication treatment. In this study, prior therapy will be registered within 3 months before randomisation, and the information about vaccinations will be collected within 6 months before screening.

The subject's CRF should also include data on any concomitant treatment performed after the enrolment and until Visit 5 (Day 28). After that, concomitant treatments regarding SAEs should be registered.

##### *8.1.4 Height, Body Weight, BMI*

Height, body weight and body mass index (BMI) will be measured at screening.

##### *8.1.5 Urine Pregnancy Test*

A urine pregnancy test will be performed for women of childbearing potential.

The test will be done at the central laboratory using urine test strips. The investigators will be provided with a separate laboratory manual containing detailed instructions on handling biological samples (collection, storage and transportation).

##### *8.1.6 Blood Analysis for Infections*

Blood analysis for significant infections at screening includes tests for syphilis (Rapid Plasma Reagin), hepatitis B (HBsAg), hepatitis C (antibodies to hepatitis C antigens) and HIV (antibodies to HIV-1 and HIV-2 and HIV-1 and HIV-2 antigens). Repeated HIV test will be performed at Visit 6 (6 months after vaccination).

The tests will be performed at the central laboratory. The investigators will be provided with a separate laboratory manual containing detailed instructions on handling biological samples (collection, storage and transportation).

##### *8.1.7 Analysis for IgM and IgG Antibodies to SARS-COV-2*

Analysis for serum IgM and IgG antibodies to the N protein and IgG antibodies to the S protein of SARS-CoV-2 will be performed for all volunteers at screening. In the case of the positive result, the volunteer will not be included in the study.

At Visit 5 (Day 28), an additional analysis for IgM and IgG antibodies to the N protein of SARS-CoV-2 will be performed. This analysis is required to detect possible SARS-CoV-2 infection after the volunteer's enrolment and vaccination.

The analysis will be performed at the central laboratory using the EIA. The investigators will be provided with a separate laboratory manual containing detailed instructions on handling biological samples (collection, storage and transportation).

##### *8.1.8 Analysis of SARS-COV-2 RNA (PCR)*

The analysis will be performed for all volunteers at screening. In the case of the positive result, the volunteer will not be included in the study. To perform the analysis, a nose and throat swab will be collected by appropriately trained trial site personnel.

If COVID-19 is suspected in the subject during the study, two SARS-CoV-2 RNA tests will be performed with an interval of 3 days (section 8.3).
The analysis for SARS-COV-2 RNA will be performed at the central laboratory using polymerase chain reaction (PCR). The investigators will be provided with a separate laboratory manual containing detailed instructions on handling biological samples (collection, storage and transportation).

Date and time of the smear, analysis results and date should be recorded in the CRF.

#### 8.2 Assessment of Vaccine Efficacy

The list of primary and secondary efficacy endpoints is provided in section 5.3.1.

The analysis for serum antibodies against the receptor-binding domain (RBD) and S protein of SARS-CoV-2 will be performed on Day 0, Day 14, Day 28 and Month 6 after vaccination. The pre-existing antibody titer will be the titer detected before the investigational product administration (on Day 0). The analysis will be performed at the independent central laboratory using the EIA.

Neutralising antibodies against SARS-CoV-2 and cellular immune response endpoints (the number of IFNy-secreting T cells [ELISpot]; percentage of CD4+ and CD8+ T cells expressing IFNy, TNF and IL-2 [flow cytometry]) will be analysed on Day 0, Day 14, Day 28 and Month 6 after vaccination. The analysis will be performed at the independent central laboratory. The investigation of the cellular immune response will be performed in a separate cohort of at least 60 subjects randomised at trial sites in Moscow.

Neutralising antibodies against Ad5 will be analysed on Day 0, Day 28 and Month 6 after vaccination. The analysis will be performed at the independent central laboratory.

Additionally, blood samples will be collected from all study participants on Day 0 (3 samples), Day 14 (2 samples), Day 28 (3 samples) and Month 6 (3 samples) after vaccination for the exploratory analysis of secondary immune response endpoints (additional assessments of neutralising antibodies, etc.). The tests will be performed at the central laboratory, and the results will be analysed outside the framework of the clinical trial report.

The investigators will be provided with a separate laboratory manual containing detailed instructions on handling biological samples (collection, storage and transportation).

#### 8.3 Registration of COVID-19 Cases

Registration of COVID-19 cases will be performed for 6 months after vaccination via observation and phone contacts with study participants.
In the case of signs and symptoms related to COVID-19, the volunteers should immediately contact the study doctor by phone.

A suspected COVID-19 case means the presence of the following clinical manifestations of acute respiratory infection and the lack of any other reasons explaining the subject's condition regardless of epidemiological anamnesis:

- body temperature > 37.5 °C

and at least one the following:

- cough (dry or with scanty sputum)
- dyspnoea
- chest congestion
- sore throat
- nasal congestion or moderate rhinorrhoea
- impairment or loss of smell (hyposmia or anosmia)
- loss of taste (dysgeusia)
- conjunctivitis
- fatigue
- muscle pain
- headache
- vomiting
- diarrhoea
- skin rash.

If COVID-19 is suspected, the volunteer will have a SARS-CoV-2 RNA test at home (throat swab) using the PCR to detect the SARS-CoV-2 infection. In 3 days, the test will be repeated. Any positive result will be considered as a confirmed COVID-19 case.

If COVID-19 is confirmed, the volunteer will be required to report this to a local outpatient clinic or other medical institution, where he/she receives medical care. Further follow-up of the volunteer will be performed at the outpatient medical institution. After the recovery and required self-isolation period, the subject may resume visits to the trial site to undergo procedures scheduled according to the Protocol. To the first visit after the recovery, the volunteer should bring a discharge summary issued by the medical institution, where he/she has been receiving medical care due to the confirmed COVID-19 case.

The study doctor will record the information about the COVID-19 case on a separate form of the subject's CRF including the following information:

- onset and end dates
- severity:
    - mild
    - moderate
    - severe
    - extremely severe
- need for hospitalisation (yes/no/unknown)
- fatality (yes/no/unknown).

To collect the information, additional phone calls may be made.

Analysis of confirmed COVID-19 cases is the exploratory efficacy endpoint and they should not be registered as adverse events.

#### 8.4 Assessment of Safety

##### *8.4.1 Specification of Safety Parameters*

The list of safety parameters is provided in section 5.3.2.

##### *8.4.2 Methods and Timing for Assessing, Recording, and Analysing Safety Parameters*

###### *8.4.2.1 Reactogenicity Evaluation*

The evaluation of reactogenicity is performed by the investigator during visits on the day of vaccination and within the following 7 days (Visit 1/Day 0 (in 20 minutes (± 5 minutes), 2 hours (± 10 minutes), and 5–8 hours after vaccination), Visit 2/Day 2, Visit 3/Day 7) as a part of the registration of adverse events.

Reactogenicity of the investigational product will be evaluated by the study doctor according to local (injection site) and general (systemic) reactions to the vaccine administration. These reactions are expected reactions to the investigational vaccine administration, and they should be assessed for all study subjects.

Local reactions include redness, swelling, tightness/infiltration, pain, itching, arm weakness.
In the case of a local reaction, the corresponding AE should be registered, and its intensity should be assessed according to section 8.4.2.8.2.

General reactions include body temperature increase, fatigue, headache, loss of appetite, nausea, vomiting, diarrhoea, sore throat, cough, difficulty breathing (dyspnoea), myalgia, arthralgia. In the case of a general reaction, the corresponding AE should be registered, and its intensity should be assessed according to section 8.4.2.8.2.

###### *8.4.2.2 Physical Examination*

A comprehensive physical examination will be performed at screening and limited physical examinations will be carried out during the subsequent visits. At the vaccination visit, a limited physical examination will be performed before the vaccine administration and in 2 hours (± 10 minutes) after the administration.

The physical examination will include the examination of organs and systems described in the following table.

**Table 15 Systems for Physical Examination**

| Organ system | |
|---|---|
| *Comprehensive examination* | *Limited examination* |
| General appearance | General appearance |
| Ears, nose, throat | Ears, nose, throat |
| Skin and injection site | Skin and injection site |
| Lymph nodes | Lymph nodes |
| Cardiovascular system | Cardiovascular system |
| Respiratory system | Respiratory system |
| Nervous system | Nervous system |
| Abdominal organs | |
| Renal system | |
| Musculoskeletal system | |

During the physical examination, the investigator will assess each system as normal/abnormal. Clinically significant abnormal findings will be reported as adverse events.

###### *8.4.2.3 Vital Signs*

Vital signs will be measured at screening and during each subsequent visit. At the vaccination visit, vital signs will be measured before the vaccine administration and in 2 hours (± 10 minutes) after the administration.

Vital signs will be measured at rest (in the standing or sitting position) before blood sampling for laboratory assessments to register the following parameters:

- systolic arterial pressure (SAP) (mm Hg)
- diastolic arterial pressure (DAP) (mm Hg)
- heart rate (HR) (bpm)
- respiratory rate (RR) (breaths per minute).

Clinically significant vital sign abnormalities should be registered as adverse events.

###### *8.4.2.4 Body Temperature*

Axillary body temperature (°C) will be measured at screening and during each subsequent visit. At the vaccination visit, body temperature will be measured before the vaccine administration, in 20 minutes (± 5 minutes) and 2 hours (± 10 minutes) after the administration.

Additionally, the volunteers should measure the body temperature at home by themselves within 7 days after vaccination (in 5–8 hours after the vaccine administration on the day of immunisation and then twice daily (morning and evening) until Visit 3 (Day 7)). During the phone contact with the study doctor on Day 0 and during trial site visits, the volunteers should tell the study doctor about all increases in body temperature.

Body temperature of 37.1 °C or higher should be registered as an adverse event.

###### *8.4.2.5 Electrocardiography*

ECG will be performed at screening and on Day 2 according to the standard practice of the trial site.

At screening, the results obtained not more than 30 days before screening may be also used.

The following parameters will be recorded in the CRF:

- heart rate (HR)
- RR, PQ, QT intervals
- QRS complex
- corrected QT (QTc).

The rhythm record in the relevant leads must contain assessable data of at least three cardiac cycles. Clinically significant deviations in ECG parameters will be reported as adverse events.

*8.4.2.6 Laboratory Tests*

Laboratory tests for the safety assessment will be carried out at the central laboratory. The investigators will be provided with a separate laboratory manual containing detailed instructions on handling biological samples (collection, storage and transportation).

Laboratory parameters to be evaluated during the study are listed in the table below.

Adverse effects related to laboratory abnormalities should be registered according to section 8.4.2.8.2.

Clinically significant laboratory abnormalities will be reported as adverse events.

**Table 16 Laboratory Parameters**

| **Laboratory test** | **Visits** | **Parameters** |
|---|---|---|
| Haematology | Screening, Day 2, Day 28 | Haemoglobin, haematocrit, RBC, erythrocyte sedimentation rate (ESR), platelets, WBC and WBC differential (neutrophils, lymphocytes, monocytes, eosinophils and basophils (% and absolute counts)) |
| Biochemistry | Screening, Day 2, Day 28 | Total protein, alanine aminotransferase (ALT), aspartate aminotransferase (AST), alkaline phosphatase (ALP), lactate dehydrogenase (LDH), total bilirubin, creatinine, urea, fasting glucose, C-reactive protein |
| Coagulation test | Screening, Day 2, Day 28 | Activated partial thromboplastin time (aPTT), prothrombin time (PT), fibrinogen |
| Urinalysis | Screening, Day 2, Day 28 | Specific gravity, pH, protein, glucose, erythrocytes, leucocytes, casts |

*8.4.2.7 Blood IgE test*

Evaluation of the IgE level will be carried out to assess the allergenic effect of the investigational products. Blood sampling will be performed during the screening period and on Day 28.

The analysis will be carried out at the central laboratory using the EIA. The investigators will be provided with a separate laboratory manual containing detailed instructions on handling biological samples (collection, storage and transportation).

*8.4.2.8 Adverse Events*

An adverse event (AE) is defined as any untoward medical occurrence in a clinical study subject administered a pharmaceutical product and which does not necessarily have a causal relationship with this treatment.

An adverse event can therefore be any unfavourable and unintended sign, symptom (including an abnormal laboratory finding), or disease temporally associated with vaccination (not registered in a volunteer before the study or occurring during the study, or, if pre-existing, worsening during the study after vaccination with investigational products), whether or not causally related to the investigation product.

An adverse drug reaction means any adverse event, the causal relationship of which with vaccination with the investigational products is assessed at least as possible (possible, probable, definite), i.e. the relationship cannot be ruled out.

**An unexpected adverse drug reaction** is an adverse event, the nature or severity of which is not consistent with the applicable vaccine information provided in the Investigator's Brochure.

**A serious adverse event (SAE)** is any adverse event that:

- results in death
- life-threatening
- requires inpatient hospitalisation or prolongation of existing hospitalisation except for planned

hospitalisation (before this study), in-hospital treatment of the AE not considered as a SAE and not resulted in hospitalisation, or social admission

- results in persistent or significant disability/incapacity
- is a congenital anomaly/birth defect in a neonate/infant born to a subject exposed to the investigational product.

**Life-threatening** means that the volunteer has been at risk of death at the time of the adverse event in the opinion of the investigator. This does not include events that could theoretically result in the volunteer's death if they would be more severe.

**Persistent or significant disability/incapacity** means an event resulted in persistent or significant interference with normal daily activities.

Moreover, serious adverse events may include important medical manifestations, if based on the medical experience the investigator believes that without medical intervention the manifestation may lead to one of the above outcomes. Examples of such events are intensive treatment in an emergency room or at home for allergic bronchospasm or convulsions that do not result in hospitalisation.

**A serious unexpected adverse drug reaction (SUADR)** is a serious adverse drug reaction, the nature and severity of which is not consistent with the applicable product information provided in the Investigator's Brochure.

*8.4.2.8.1 Methods for Identification and Registration of Adverse Events*

The investigator is responsible for AE registration during the clinical study.

It is necessary to register AEs identified in volunteers from the time of vaccination until the end of the study. From Visit 0 to Visit 5, all AEs should be registered. From Visit 5 to Visit 6, only SAEs should be registered. Adverse events should be registered sequentially as they occur.
Adverse events are recorded based on complaints reported by the volunteer him/herself, results of an interview of the investigator with the volunteer, results of the general or instrumental examinations, and laboratory tests. The investigator will formulate his/her questions in such a way that they do not provoke volunteers to report false information. For example, a question such as "Have any aspects of your health changed since the last visit/interview?" may be asked.

All adverse events should be documented in the source documents and CRF on the Adverse Events form.

The following information will be specified upon registration of an adverse event:

- subject's identification number
- nature of the adverse event (it is preferable to state the diagnosis, not the list of symptoms)
- date of onset and resolution of the event (and time if applicable)
- AE severity according to the investigator
- causality with vaccination according to the investigator
- actions taken due to the AE
- correspondence of the adverse event to criteria of a serious adverse event
- outcome of the event.

*8.4.2.8.2 Severity of Adverse Events*

**Temperature Response**
The severity of temperature response (axillary body temperature increase) should be assessed using the table below.

**Table 17 Temperature Response Severity Assessment**

| | Mild (Grade 1) | Moderate (Grade 2) | Severe (Grade 3) | Life-threatening (Grade 4) |
|---|---|---|---|---|
| Fever (axillary body temperature increase) | 37.1–38.4 | 38.5–38.9 | 39.0–40 | > 40 |

**Local and Systemic Reactions to the Vaccination (Reactogenicity Assessment), Other Clinical and Laboratory Abnormalities**

The assessment of adverse event severity should be performed according to the FDA Guidance on Toxicity Grading Scale for Healthy Adult and Adolescent Volunteers Enrolled in Preventive Vaccine Clinical Trials (September 2007), https://www.fda.gov/media/73679/download

Tables to grade the severity of reactogenicity and other clinical and laboratory abnormalities from this guidance in English is provided in Appendix 1.

**Other Adverse Events**

The severity of adverse events not related to body temperature and not listed in Appendix 1 (as well laboratory abnormalities if the grading according to Appendix 1 is not possible) should be determined by the investigator based on the following criteria:

- **Mild (Grade 1)** — transient AE that does not require treatment or therapeutic intervention and does not interfere with the daily activity of the subject.
- **Moderate (Grade 2)** — AE that causes some interference with activity and does not require treatment.
- **Severe (Grade 3)** — AE that prevents daily activity and requires medical intervention.
- **Life-threatening (Grade 4)** — an immediate threat to life or AE leading to permanent mental or physical conditions that require ER visit or hospitalisation.

*8.4.2.8.3 Causal Relationship of Adverse Events with Vaccination*

Investigator's opinion of the causal relationship of an AE to the investigational vaccine should be based on clinical information available at the time of the completion of the CRF.

The causal relationship should be graded according to the following scale:

- **Definite** — clinical manifestations develop within a specific time interval after the vaccine administration and cannot be explained by any concurrent disease or administration of other drug products.
- **Probable** — available data confirm causal relationship with the vaccine administration; the influence of any concurrent disease or administration of other drug products is unlikely.
- **Possible** — available data confirm the causal relationship with the vaccine administration; it may also relate to any concurrent disease or administration of any drug product.
- **Doubtful** — manifestations of the AE are not temporary related to the vaccine administration; there are other factors (drugs, diseases, chemical compounds) that may be the reason for their occurrence.
- **Not related** — AE does not occur within the acceptable time interval after the vaccine administration; it may be explained by other diseases or administration of other drug products.
- **Unknown** — it is not possible to assess the causal relationship with the vaccine administration due to limited information and reasons for the clinical manifestation.

An AE is considered related to vaccination if the causal relationship is assessed at least as possible (possible, probable, definite), i.e. the relationship cannot be ruled out.

*8.4.2.8.4 Assessment of Adverse Event Outcome*

An outcome of an adverse event should be recorded in the source documents and subject's CRF using the following categories:

- resolved
- resolving

- not resolved
- resolved with sequelae
- death
- unknown.

*8.4.2.8.5 Follow-up of Adverse Events*

If an adverse event or its complications are not resolved, the investigator should follow up the volunteer. The adverse event or its complications should be followed up until their resolution or stabilisation (judged acceptable by the investigator), or until the determination of the reason of such adverse event not related to the use of the investigational product.

*8.4.2.8.6 Investigator's Responsibilities on Reporting of Serious Adverse Events*

The investigator should immediately report on all SAEs developed in the volunteers from the time of vaccination until the end of the study. In the case of an SAE, the investigator should:

- Provide the volunteer with necessary medical care.
- Complete the SAE reporting form and send it via fax or email to the Qualified Person for Pharmacovigilance (QPPV) within not more than 24 hours from the moment of its discovery.
- Inform the LEC of the medical institution within the period specified in the committee regulations.

If additional information about the SAE becomes available that has not been included in the initial SAE report, it is necessary to complete the second SAE report with the indication of such information and send it to the QPPV within 24 hours.

Before sending the SAE report, it is necessary to inform the QPPV or study monitor about the fax or email by phone.

If investigators have any additional questions, they should contact the study monitor of the trial site or the QPPV.

In the case of registration of an SAE related to the use of the investigational product, the report must be submitted even if the study has been completed.

Requests to provide further information received after the initial SAE report must be forwarded to the trial site by the QPPV (directly or through the monitor of the trial site) not later than 5 (for SAEs related to the study drug) or 30 (for SAEs not related to the study drug) days and continue with the same frequency until the complete resolution of the SAE or stabilisation of the subject's condition.

| |
|---|
| **In the case of a serious adverse event, the appropriate SAE form should be immediately completed and sent by fax and/or email to the Qualified Person for Pharmacovigilance (QPPV):**<br>**Fax:** +7 (495) 730 75 60<br>**E-mail:** |

###### *8.4.2.8.7 Other Investigator's Reports*

The investigator should report to the LEC of the medical institution on serious unexpected adverse drug reactions (SUADR) presumably related to the investigational product and other aspects of the treatment safety, which are subject to immediate reporting, including if they may influence the risk-benefit assessment of the vaccine or require significant changes in the study methodology.

###### *8.4.2.8.8 Sponsor's Responsibilities to Provide Safety Information in the Study*

**Reporting of Serious Unexpected Adverse Drug Reactions (SUADR) Presumably Related to the Investigational Product**

The QPPV will be submitting expedited reports on SUADRs to regulatory authorities.

The QPPV will be also reporting to investigators on all SUADRs registered during the study.

SUADRs that lead to death or may be life-threatening for a volunteer should be submitted to regulatory authorities within not more than 7 calendar days once the information on such event is obtained by the QPPV, and second reports with the necessary additional information should be submitted within the following 15 calendar days.

**Submission of Other Treatment Safety Reports**

- The QPPV will be submitting to regulatory authorities expedited reports on other aspects of vaccination safety if they affect the risk-benefit assessment of the vaccination or may require significant changes in the dosing regimen or study methodology.
- In addition to submission of expedited reports, the Sponsor will be preparing annual reports on vaccine safety throughout the whole study, including all new information obtained during the reported period. Such reports will be submitted to regulatory authorities at least once a year or with other intervals at the request of such authorities.

###### *8.4.2.8.9 Pregnancy*

The pregnancy itself is not an adverse event, except when there are grounds to believe that the administration of the investigational product has led to reduced efficacy of contraceptives. Any congenital anomalies and malformations in children of the volunteers are serious adverse events. Scheduled abortions carried out by medical indications, as well as any serious complications during the pregnancy (including spontaneous abortions), will be recorded as serious adverse events. Scheduled abortions without complications are not adverse events.

All pregnancies recorded during the study (including pregnancies of sex partners of the volunteers taking part in the study) will be properly registered.

In the case of confirmed pregnancy, the investigator should notify the contract research organisation/Sponsor about it within not more than 24 hours after obtaining the information by sending a completed pregnancy report form. Then, the information on the pregnancy outcome should be submitted. Pregnancy will be registered from the time of the vaccine administration until the last procedure conducted during the study.

The outcome of each pregnancy case (spontaneous abortion, elective abortion, birth of a normal child or a child with congenital anomalies or malformations) must be also registered.

During the collection of medical history at screening, pregnancies occurred within 30 days before vaccine administration should be registered and evaluated.

##### *8.4.3 Acceptability of Endpoints*

Methods of efficacy evaluation selected for this trial are widely used during clinical and epidemiology studies of vaccines. As far as there are no special criteria to evaluate vaccine efficacy against novel coronavirus infection COVID-19, the criteria used in the previous studies of Ad5- nCoV have been chosen. The safety assessment methods used in this trial are generally accepted for clinical trials of vaccines.

### 9 DATA MANAGEMENT AND STATISTICAL ANALYSIS

#### 9.1 Sample Size Calculation and Justification

To determine the number of subjects required to test the hypothesis relating to the primary variable (proportion of subjects with seroconversion on Day 28 after vaccination), the following conservative assumptions are used:

- study power is about 90 %
- two-sided significance level (alpha) is 5 %; Pocock's corrected alpha level for multiple comparisons of the primary variable due to an interim analysis is 0.02616 and 0.03039 for the interim and final analysis (performed after obtaining primary variable data for all randomised subjects), respectively
- proportion of subjects with seroconversion in the placebo group is 20 %
- proposed difference between the placebo and vaccine group in terms of the primary variable is 30–60 % (odds ratio is 4–16)
- randomisation ratio between the vaccine and placebo group is 3:1, respectively
- two countries (accrual will be competitive, so a uniform assignment is considered for the calculation)
- dropout from the study/statistical analysis during the observation period of 28 days is about 10 %.

According to the results of the sample size calculation performed using the PASS 12 software (Professional License, NCSS LLC (www.ncss.com)), 180 subjects should be included in the statistical analysis to ensure the power of 90 % for the between-group comparison of the primary variable considering the assumption of 20 % seroconversion rate in the placebo group and the superiority of the vaccine group of at least 30 % (conservative assumption, odds ratio = 4), as well as the corrected two-sided significance level of 0.02616 (one-sided level of 0.01308) and randomisation ratio of 3:1. The number of subjects corresponding to the conservative estimate of differences between the vaccine and placebo groups in terms of the primary variable is chosen to ensure the provision of safety data. Considering the possible dropout from the study/statistical analysis during the observation period of 28 days (10 % of subjects), the number of randomised subjects should be increased up to 200 (in a 3:1 ratio).

To provide more detailed safety and efficacy data (including age subgroups), it is planned to randomise 500 subjects (in a 3:1 ratio [Ad5-nCoV:placebo]). Additionally, the extended sample size will allow to provide descriptive data on exploratory endpoints related to the frequency of confirmed COVID-19 cases within 6 months after vaccination (except for the cases occurred during the first 14 days after vaccination).

Because the study includes an unblinded interim analysis after obtaining a fraction of information ($\tau$ = 0.4) for the assessment of the primary variable (based on the data obtained approximately from the first 200 randomised volunteers through Visit 5, including early dropouts before Visit 5) and the Pocock alpha spending function is used, to adjust the significance level due to multiple comparisons of the primary variable (interim analysis), the two-sided alpha level for the interim and final analysis is set to be 0.02616 and 0.03039, respectively (overall two-sided significance level is 5 %). If statistically significant findings of the between-group comparison of the primary variable are obtained during the interim analysis, the study will not be suspended and the interim analysis results will be provided to regulatory authorities for review.

**Sample Calculation Results Using PASS 12 — Professional License, NCSS LLC (www.ncss.com)**

**Tests for Two Proportions in a Stratified Design (Cochran/Mantel-Haenszel Test)**

**Numeric Results of Cochran-Mantel-Haenszel Test of an Odds Ratio**
**H0: OR1 = OR0. H1: OR1 > OR0.**

| Power | Total Sample Size (N) | Sample Size Multiplier (M) | Sample Size of Group 1 (N1) | Sample Size of Group 2 (N2) | H0 Odds Ratio (OR0) | Actual Odds Ratio (OR1) | Signif. Level Alpha | Beta |
|---|---|---|---|---|---|---|---|---|
| 0.9000 | 178 | 178.045 | 134 | 45 | 1.000 | 4.000 | 0.0131 | 0.1000 |
| 0.9000 | 134 | 133.703 | 100 | 33 | 1.000 | 4.900 | 0.0131 | 0.1000 |
| 0.9000 | 104 | 104.278 | 78 | 26 | 1.000 | 6.000 | 0.0131 | 0.1000 |
| 0.9000 | 67 | 67.243 | 50 | 17 | 1.000 | 9.300 | 0.0131 | 0.1000 |
| 0.9000 | 45 | 45.123 | 34 | 11 | 1.000 | 16.000 | 0.0131 | 0.1000 |

**Strata-Detail Report**

| Number of Strata | Proportion of Total Sample in each Strata | Proportion of this Strata in Group 1 | Proportion of this Strata in Group 2 | Group 1 Multiplier (R1) | Group 2 Multiplier (R2) | Strata Probability of Success |
|---|---|---|---|---|---|---|
| 2 | 0.5000 | 0.7500 | 0.2500 | 0.375 | 0.125 | 0.2000 |

**Report Definitions**
Power: the probability of rejecting a false null hypothesis.
N: the total sample size summed across all groups and strata.
M: the factor by which the values of R1 and R2 are multiplied.
N1 and N2: the sample sizes from groups 1 and 2 summed across all strata.
OR0: the odds ratio [P1/(1-P1)] / [P2/(1-P2)] assuming the null hypothesis (H0). OR1: the value of the odds ratio at which the power is computed.
Alpha: the probability of rejecting a true null hypothesis.
Beta: the probability of accepting a false null hypothesis.
In a treatment vs. control design, the treatment group is 1 and the control group is 2.

Note: P1 and P2 are the proportion of subjects with an event in the investigational product group and control group, respectively.

#### 9.2 Statistical Analysis Plan

##### *9.2.1 Statistical Analysis Sets*

The following analysis sets will be used:

*Randomised Set*

This population will include subjects completed the randomisation and allocated into one of the study groups. This population will be used to provide the disposition of study subjects.

*Safety Analysis Set*

This population will include all randomised volunteers received a dose of the vaccine. It will be used for safety analysis.

*Full Analysis Set (FAS, for immunogenicity analysis)*

This analysis set will include all eligible volunteers from the safety population with at least one immunogenicity assessment result (for interim analysis, it should be obtained until Visit 5 [inclusive]). This set will be the main population for immunogenicity analysis.

Cellular immune response parameters will be analysed in the FAS subgroup for the analysis of vaccine efficacy including subjects with at least one cellular immunity assessment result.

*Per-protocol Set (PPS, for immunogenicity analysis)*

The population will include all eligible volunteers from the FAS received a dose of the vaccine according to the randomisation and study scheme and provided data for immunogenicity assessment before and after vaccination in line with the study scheme (for interim analysis, they should be obtained until Visit 5 [inclusive]). Such volunteers should not administer prohibited medications, should not have other major protocol deviations potentially affecting immunogenicity assessment and should not have confirmed

COVID-19 cases within 14 days after vaccination. This set will be a supportive population for immunogenicity analysis.

*Full Analysis Set (FAS, for efficacy analysis)*

The population will include all eligible volunteers from the Safety Analysis Set contacted at least once after Day 14 excluding subjects with confirmed COVID-19 cases within 14 days after vaccination. This set will be the main population for efficacy analysis.

*Per-protocol Set (PPS, for efficacy analysis)*

The population will include all eligible volunteers from the FAS received a dose of the vaccine according to the randomisation and study scheme. Such volunteers should not administer prohibited medications, should not have other major protocol deviations potentially affecting efficacy assessment and should not have confirmed COVID-19 cases within 14 days after vaccination. This set will be a supportive population for efficacy analysis.

##### 9.2.2 General Provisions

A detailed description of statistical methods will be provided in the Statistical Analysis Plan before the database lock. General information on the proposed statistical analysis is provided below.

Descriptive statistics will be provided for all demographic and other baseline characteristics, immunogenicity, cellular immune response, efficacy and safety parameters, as well as their changes (if applicable) during the study by evaluation time points and treatment groups. Demographic characteristics and efficacy endpoints will also be presented by the pre-existing anti-Ad5 neutralising antibody titers (low and negative (≤ 1:200) vs high (> 1:200)). Descriptive statistics for quantitative parameters will include the mean, the standard deviation (SD), the median, the first and third quartiles, the minimum and maximum values and the number of valid observations. The geometric mean values will be also presented for antibody titres and fold-rises in titers. Qualitative data will be presented as rates and percentage. Where applicable, 95 % confidence intervals will also be provided.

For undetermined values of the geometric mean titer, geometric mean fold-rise and seroconversion rate, the following algorithm will be used: 1) if the antibody titer is below the lower limit of quantitation, a half LoD value will be used 2) if the antibody titer is above the maximum limit of quantitation, the maximum dilution will be used.

*Missing Data*

Only available values will be used for statistical analysis. Missing values will not be imputed.

*Procedures for Reporting Any Deviations from the Original Statistical Plan*

In the case of any deviations from the planned statistical analysis described in the protocol, the clinical trial report will include the description and justification of implemented changes.

##### 9.2.3 Efficacy Analysis

The primary variable will be analysed during the interim and final analysis using the corrected significance level (overall significance level is two-sided 5 %). Secondary variables will be analysed using the two-sided 5 % significance level.

Efficacy variables representing the seroconversion rate (proportion of subjects with at least four-times increase in antibody titers), including the primary efficacy endpoint, will be presented by evaluation time points and treatment groups.

To compare the seroconversion rate (including the primary efficacy endpoint) by evaluation time points, a statistical hypothesis of the statistical superiority of the vaccine against placebo will be tested:

$H_0$: OR = 1 $H_A$: OR > 1

OR = odds ratio: OR = $[P_1/(1-P_1)] / [P_2/(1-P_2)]$

$P_1$ and $P_2$ — proportion of subjects with seroconversion in the vaccine and placebo group, respectively.

The results will be descriptively provided by trial sites and countries. The primary variable will be analysed using the Mantel-Haenszel test with the country as a factor if at the time of the analysis the analysis set in each country will have a proper number of subjects with the assessed primary endpoint (at least 20 % of the planned number of subjects for the interim analysis in the vaccine and placebo groups). Otherwise, data will be analysed using the trial site as a factor. Detailed information about combining data for small trial sites will be presented in the statistical analysis plan. Hypothesis testing will be carried out using the $X$ test (or Fisher's exact test). Corrected and uncorrected two-sided confidence intervals for the percentage differences will be also presented.

Additionally, between-group comparisons will be also performed using logistic regression with the treatment group as a fixed factor, the country as a factor and pre-existing antibody titer as a covariate. Pre-existing anti-Ad5 neutralising antibody titers (low and negative (≤ 200) vs high (> 200)), sex and age will be also considered as potential model factors. Odds ratios will be presented with appropriate 95 % confidence intervals.

Similarly, the proportion of subjects with cellular immune response will be analysed.

Efficacy variable (antibody geometric mean titer (GMT) and neutralising antibody GMT) will be presented and compared by evaluation time points based on the calculated 95 % confidence interval using ANOVA/ANCOVA following the logarithmic transformation of base 10. The ANCOVA model will include the treatment group as a fixed factor, country/trial site as a factor and pre-existing (log-transformed) antibody titer as a covariate. Pre-existing anti-Ad5 neutralising antibody titers (low and negative (≤ 200) vs high (> 200)[1]), sex and age will be also considered as potential model factors. Mean log-transformed differences between the study groups (vaccine/placebo) will be evaluated with the corresponding 95 % confidence interval. The point estimates of the mean differences and the corresponding confidence intervals will be back-transformed.

Efficacy variable (geometric mean fold-rise in antibody titers and neutralising antibody titers) for each treatment group and each antibody GMT evaluation time point will be assessed with the corresponding two-sided 95 % CIs following log-transformation (log10), point estimation of the difference and confidence intervals, and back-transformation of obtained values. Between-group comparisons will be performed using ANOVA/ANCOVA following the logarithmic transformation of base 10. The ANCOVA model will include the treatment group as a fixed factor, country/trial site as a factor and pre-existing (log-transformed) antibody titer as a covariate. Pre-existing anti-Ad5 neutralising antibody titers (low and negative (≤ 200) vs high (> 200)[1]), sex and age will be also considered as potential model factors.

Exploratory efficacy endpoints (frequency of confirmed COVID-19 cases during 6 months after vaccination, frequency of confirmed COVID-19 cases requiring hospitalisation, frequency of severe COVID-19 cases, and frequency of lethal COVID-19 cases (except for the cases occurred during the first 14 days after vaccination)) will be presented descriptively by treatment groups. Kaplan-Meier survival estimates will be also presented if applicable.

##### *9.2.4 Safety Analysis*

The safety analysis will be performed in the safety analysis set using descriptive methods.

Adverse events will be coded using the MedDRA dictionary. The number (proportion) of volunteers with AEs/SAEs and the number of AEs/SAEs will be presented in the form of tables by system organ classes and preferred terms, as well as the relationship to vaccination and severity, and by treatment groups. Upon that, each subject will be counted only once for a system organ class and preferred term with the indication of the relationship to vaccination and maximum severity. The number (percentage) of volunteers with local and systemic immunisation reactions will also be provided.

Laboratory parameters and their changes after vaccination compared to the baseline values, as well as shift tables showing changes with respect to normal values will be tabulated by treatment groups and evaluation time points.

The results of ECG, body temperature and vital signs measurements will be tabulated by treatment groups and each evaluation time point along with changes from the last available measurement before vaccination.

The results of the physical examination will be provided descriptively by treatment groups and evaluation time points.

[1] For anti-Ad5 neutralising antibody titers, the pre-existing value will be included in the model only as a continuous variable.

Serum immunoglobulin E concentration and its changes in 28 days after vaccination compared to the baseline will be tabulated by treatment groups.

#### 9.3 Interim Analysis

One unblinded interim analysis is planned for the study, which will be performed after obtaining efficacy, reactogenicity and safety data through Visit 5 (Day 28) for the first 200 volunteers randomised in the study. The analysis will be performed by an independent statistician. If statistically significant findings of the between-group comparison of the primary variable are obtained during the interim analysis, the study will not be suspended. The results of the interim analysis will be provided to regulatory authorities to make a decision on the registration of the Ad5-nCoV vaccine.

The final clinical trial report will be prepared after obtaining all data for all volunteers randomised in the study, i.e. after the completion of the final visit by all the volunteers in 6 months after vaccination. The final clinical trial report will be also submitted to regulatory authorities.

### 10 QUALITY ASSURANCE AND QUALITY CONTROL

Quality assurance and quality control will be performed via regular monitoring of trial sites during the study, as well as through audits of the sites that may be performed upon the Sponsor's or CRO's request.

#### 10.1 Study Monitoring

At a site initiation visit, the designated monitor of the CRO will provide training for trial site personnel regarding protocol procedures, CRF completion and other aspects of the study. During the study, the monitor will maintain contact with the investigators by phone or email and will be performing regular visits to the trial sites. During the monitor's regular visits, the following aspects will be reviewed:

- completeness of source documents and accuracy of CRF records (complete verification with source documents), as well as consistency of different documents
- compliance with the requirements of the Protocol, Good Clinical Practice and local requirements of the Russian legislation
- recruitment of volunteers into the study
- observance of volunteers' rights and protection of their safety
- compliance with the requirements regarding storage, distribution and registration of the investigational product.

During the visits, trial site personnel should provide the monitor with the necessary assistance and ensure his/her access to the source documents and other required records.

The monitors will be given proper guidelines containing additional instructions on monitoring procedures.

#### 10.2 Audit and Inspection

Study audits may be performed by the Sponsor, CRO or other authorised representative of the Sponsor at any time during the study or within a reasonable period after its completion. Inspections may be also performed upon the decision of regulatory authorities. Trial site personnel should provide the auditor or inspector with access to all necessary study documents on demand and assist him/her during the audit and/or inspection.

### 11 ETHICAL ASPECTS OF THE STUDY

#### 11.1 General Requirements and Ethical Aspects of the Study

The study will be conducted in accordance with the principles of the most recent version of the World Medical Association Declaration of Helsinki, International Council for Harmonisation Good Clinical Practice (ICH GCP), as well as national regulatory requirements and standard operating procedures of the Sponsor and CRO.

The clinical study will be carried out in accordance with the Protocol. The Protocol and all protocol amendments, Subject Information Sheet and Informed Consent Form, as well as other essential study documents, should be reviewed and approved by the Ethics Council and Local Ethics Committees (LECs) of the trial sites before the beginning of the study.

Amendments to the Protocol may be made only after the review and approval of the corresponding amendment and updated Subject Information Sheet and Informed Consent Form by regulatory authorities and the LEC of the medical institution.

Protocol amendments made to eliminate immediate hazards to the volunteers may be implemented before the approval from regulatory authorities and the Sponsor. However, in this case, it is necessary to notify regulatory authorities and the LEC of the institution as soon as possible to approve the implemented amendments. Protocol amendments relating only to administrative aspects of the study may be also implemented before sending the corresponding notification to the LEC and regulatory authorities.

Member of the study team should have adequate qualifications and experience for activities related to the study and should undergo necessary training regarding the Protocol and study procedures. Qualifications of the trial site personnel involved in the study should be documented.

#### 11.2 Subject Information and Consent

At screening before any protocol required procedures, the volunteers will be invited to participate in this clinical trial and provided with the Subject Information Sheet and Informed Consent Form for review and making a decision on the participation.

After reading the information about the study, asking questions and receiving the answers, the volunteer should personally sign and date the corresponding informed consent form in duplicate. One signed and dated copy of the Subject Information Sheet and Informed Consent Form will be provided to the volunteer, and the other copy will be stored at the trial site.

The investigator should provide the volunteers with adequate information regarding the objectives and methods of the study, as well as expected benefits and possible risks associated with the participation in it. The volunteers should be given enough time to read the information, ask questions and receive answers to them. The volunteers should be informed about their right to withdraw from the study at any time without giving a reason and without any penalties.

In the case of protocol amendments and changes to documents containing information for the volunteers, such updated documents should be reviewed and approved by regulatory authorities and the LEC of the institution, as well as signed by the volunteers still participating in the study.

#### 11.3 Confidentiality

The investigator should ensure the confidentiality of personal data of the volunteers involved in the study. CRFs and other documents provided to the Sponsor and CRO will contain only the numbers of the volunteers for identification purposes. Documents that are not provided to the Sponsor (e.g. signed informed consent forms) should be kept by the investigator in compliance with strict confidentiality requirements.

On enrolling in the study, the volunteers will be notified that their medical records may be reviewed by the designated monitor, auditor of the Sponsor, LEC members and the regulatory inspector and that their personal data will be processed without disclosing any information to the extent possible according to the legislation. The volunteers should be also informed about a specific approach to the registration and transfer of their personal data obtained during the SARS-CoV-2 RNA test (PCR) in line with the national legislation.

If the results of the study are published, the subject's identity will remain confidential.

##### 11.4 Financing and Insurance

Clinical study subjects will be insured for risks associated with the threat to their life and health. The investigator should inform the participant about this insurance. Detailed information about the terms and conditions of such insurance is available in the Investigator's Site File.

This study will be financially supported by the Sponsor according to the agreements between the Sponsor and trial sites.

### 12 ADMINISTRATIVE ASPECTS OF THE STUDY

#### 12.1 Trial Documentation and Storage

##### *12.1.1 Source Documents*

Source documents include clinical and office charts, hospital records, visit logs, vaccine prescription and dispensing records, memoranda, laboratory notes containing test results, and other documents of the medical institution specified in section 1.52 of the ICH GCP. If paper copies of source documents are available, such copies should be signed and dated by an authorised employee of the trial site.

The investigator and the medical institution, in which the study is conducted, must provide access to the source documents for study monitors, auditors, representatives of the Ethics Council or LEC of the trial site and regulatory inspectors.

##### *12.1.2 Case Report Forms*

Before the initiation of the study, electronic Case Report Forms will be provided by the Sponsor. Trial site personnel authorised to complete the CRFs will undergo training on their completion.

The investigator is responsible for the completeness and accuracy of the data recorded in the CRF. Data reported in the CRF should be consistent with the source documents. The monitor will be performing a complete cross-verification of the data during his/her visits to the trial site. The investigator is responsible for timely completion of the CRF and responding to queries issued during the study or at the database lock. Study data will be processed by the authorised CRO.

##### *12.1.3 Archiving*

The investigator should archive all documents of the study after its completion according to the rules of the trial site and the procedures of the Sponsor. The documents should be archived to ensure access to them by the representatives of the Sponsor and regulatory authorities.

The investigator must keep the study documents for 15 years or until the corresponding notification by the Sponsor that such documents are no longer required (whichever occurs first). Medical records of the volunteers and other documents (including copies of protocols, CRFs, original laboratory records, vaccine accountability logs, copies of signed informed consent forms and other documents) must be kept for the longest period applicable for the medical institution. Study documents can be destructed only after a written agreement between the Sponsor and the investigator. If the investigator wishes to transfer the authority for keeping the archive to a third party or change the archive location, he must obtain written permission from the Sponsor.

#### 12.2 Publications

Exclusive rights for study results belong to the Sponsor (NPO Petrovax Pharm LLC). The investigators can publish any study results only after obtaining the Sponsor's permission. Before that, the investigator should provide the representative of NPO Petrovax Pharm LLC with a copy of the planned publication. Any information obtained during the study is confidential.

### APPENDIX 1. GRADING OF ADVERSE EVENT SEVERITY

Tables below should be used to assess the severity of clinical and laboratory abnormalities according to the FDA Guidance on Toxicity Grading Scale for Healthy Adult and Adolescent Volunteers Enrolled in Preventive Vaccine Clinical Trials (September 2007) https://www.fda.gov/media/73679/download

Temperature responses and the severity of AEs not listed in the below tables should be assessed according to the categories provided in section 8.4.2.8.2.

**Table 18 Clinical Abnormalities**

| Local reaction at the injection site | Mild (Grade 1) | Moderate (Grade 2) | Severe (Grade 3) | Life-threatening (Grade 4) |
|---|---|---|---|---|
| Pain | Does not interfere with activity | Repeated use of nonnarcotic pain reliever > 24 hours or interferes with activity | Any use of narcotic pain reliever or prevents daily activity | Emergency room (ER) visit or hospitalisation |
| Tenderness | Mild discomfort to touch | Discomfort with movement | Significant discomfort at rest | Emergency room (ER) visit or hospitalisation |
| Erythema/Redness* | 2.5–5 cm | 5.1–10 cm | > 10 cm | Necrosis or exfoliative dermatitis |
| Induration/Swelling** | 2.5–5 cm and does not interfere with activity | 5.1–10 cm or interferes with activity | > 10 cm or prevents daily activity | Necrosis |

*In addition to grading the measured local reaction at the greatest single diameter, the measurement should be recorded as a continuous variable.

**Induration/Swelling should be evaluated and graded using the functional scale as well as the actual measurement.

| Systemic (General) | Mild (Grade 1) | Moderate (Grade 2) | Severe (Grade 3) | Life-threatening (Grade 4) |
|---|---|---|---|---|
| Nausea/vomiting | No interference with activity or 1–2 episodes/24 hours | Some interference with activity or > 2 episodes/24 hours | Prevents daily activity, requires outpatient IV hydration | ER visit or hospitalisation for hypotensive shock |
| Diarrhoea | 2–3 loose stools or < 400 g/24 hours | 4–5 stools or 400–800 g/24 hours | 6 or more watery stools or > 800 g/24 hours or requires outpatient IV hydration | Emergency room (ER) visit or hospitalisation |
| Headache | Does not interfere with activity | Repeated use of nonnarcotic pain reliever > 24 hours or some interference with activity | Significant; any use of narcotic pain reliever or prevents daily activity | Emergency room (ER) visit or hospitalisation |
| Fatigue | Does not interfere with activity | Some interference with activity | Significant; prevents daily activity | Emergency room (ER) visit or hospitalisation |
| Myalgia | Does not interfere with activity | Some interference with activity | Significant; prevents daily activity | Emergency room (ER) visit or hospitalisation |
| Illness or clinical adverse event | Does not interfere with activity | Some interference with activity not requiring medical intervention | Prevents daily activity and requires medical intervention | Emergency room (ER) visit or hospitalisation |

**Table 19 Laboratory Abnormalities**

The laboratory values provided in the tables below serve as guidelines and are dependent upon institutional normal parameters. Institutional normal reference ranges should be provided to demonstrate that they are appropriate.

| Serum* | Mild (Grade 1) | Moderate (Grade 2) | Severe (Grade 3) | Life-threatening (Grade 4)** |
|---|---|---|---|---|
| Sodium – Hyponatremia mEq/L | 132–134 | 130–131 | 125–129 | < 125 |
| Sodium – Hypernatremia mEq/L | 144–145 | 146–147 | 148–150 | > 150 |
| Potassium – Hyperkalaemia mEq/L | 5.1–5.2 | 5.3–5.4 | 5.5–5.6 | > 5.6 |
| Potassium – Hypokalaemia mEq/L | 3.5–3.6 | 3.3–3.4 | 3.1–3.2 | < 3.1 |
| Glucose – Hypoglycaemia mg/dL | 65–69 | 55–64 | 45–54 | < 45 |
| Glucose – Hyperglycaemia□<br>Fasting – mg/dL□<br>Random – mg/dL | 100–110<br>110–125 | 111–125<br>126–200 | > 125<br>> 200 | Insulin requirements or hyperosmolar coma |
| Blood Urea Nitrogen□<br>BUN mg/dL | 23–26 | 27–31 | > 31 | Requires dialysis |
| Creatinine – mg/dL | 1.5–1.7 | 1.8–2.0 | 2.1–2.5 | > 2.5 or requires dialysis |
| Calcium – hypocalcaemia mg/dL | 8.0–8.4 | 7.5–7.9 | 7.0–7.4 | < 7.0 |
| Calcium – hypercalcemia mg/dL | 10.5–11.0 | 11.1–11.5 | 11.6–12.0 | > 12.0 |
| Magnesium – hypomagnesemia mg/dL | 1.3–1.5 | 1.1–1.2 | 0.9–1.0 | < 0.9 |
| Phosphorous – hypophosphatemia mg/dL | 2.3–2.5 | 2.0–2.2 | 1.6–1.9 | < 1.6 |
| CPK – mg/dL | 1.25–1.5 x ULN*** | 1.6–3.0 x ULN | 3.1–10 x ULN | > 10 x ULN |
| Albumin – Hypoalbuminemia g/dL | 2.8–3.1 | 2.5–2.7 | < 2.5 | -- |
| Total Protein – Hypoproteinaemia g/dL | 5.5–6.0 | 5.0–5.4 | < 5.0 | -- |
| Alkaline phosphate – increase by factor | 1.1–2.0 x ULN | 2.1–3.0 x ULN | 3.1–10 x ULN | > 10 x ULN |
| Liver Function Tests – ALT, AST increase by factor | 1.1–2.5 x ULN | 2.6–5.0 x ULN | 5.1–10 x ULN | > 10 x ULN |
| Bilirubin – when accompanied by any increase in Liver Function Test increase by factor | 1.1–1.25 x ULN | 1.26–1.5 x ULN | 1.51–1.75 x ULN | > 1.75 x ULN |
| Bilirubin – when Liver Function Test is normal; increase by factor | 1.1–1.5 x ULN | 1.6–2.0 x ULN | 2.0–3.0 x ULN | > 3.0 x ULN |
| Cholesterol | 201–210 | 211–225 | > 226 | --- |
| Pancreatic enzymes – amylase, lipase | 1.1–1.5 x ULN | 1.6–2.0 x ULN | 2.1–5.0 x ULN | > 5.0 x ULN |

*The laboratory values provided in the tables serve as guidelines and are dependent upon institutional normal parameters. Institutional normal reference ranges should be provided to demonstrate that they are appropriate.

**The clinical signs or symptoms associated with laboratory abnormalities might result in the characterisation of the laboratory abnormalities as Potentially Life-Threatening (Grade 4). For example. a low sodium value that falls within a grade 3 parameter (125–129 mE/L) should be recorded as a grade 4 hyponatremia event if the subject had a new seizure associated with the low sodium value.

***ULN is the upper limit of the normal range.

| **Haematology*** | **Mild (Grade 1)** | **Moderate (Grade 2)** | **Severe (Grade 3)** | **Life-threatening (Grade 4)** |
|---|---|---|---|---|
| Haemoglobin (Female) – gm/dL | 11.0–12.0 | 9.5–10.9 | 8.0–9.4 | < 8.0 |
| Haemoglobin (Female) change from base-line value – gm/dL | Any decrease – 1.5 | 1.6–2.0 | 2.1–5.0 | > 5.0 |
| Haemoglobin (Male) – gm/dL | 12.5–13.5 | 10.5–12.4 | 8.5–10.4 | < 8.5 |
| Haemoglobin (Male) change from base-line value – gm/dL | Any decrease – 1.5 | 1.6–2.0 | 2.1–5.0 | > 5.0 |
| WBC Increase – cell/mm$^3$ | 10,800–15,000 | 15,001–20,000 | 20,001–25,000 | > 25,000 |
| WBC Decrease – cell/mm$^3$ | 2500–3500 | 1500–2499 | 1000–1499 | < 1000 |
| Lymphocytes Decrease – cell/mm$^3$ | 750–1000 | 500–749 | 250–499 | < 250 |
| Neutrophils Decrease – cell/mm$^3$ | 1500–2000 | 1000–1499 | 500–999 | < 500 |
| Eosinophils – cell/mm$^3$ | 650–1500 | 1501–5000 | > 5000 | Hypereosinophilic |
| Platelets Decreased – cell/mm$^3$ | 125,000–140,000 | 100,000–124,000 | 25,000–99,000 | < 25,000 |
| PT – increase by factor (prothrombin time) | 1.1–1.10 x ULN** | 1.11–1.20 x ULN | 1.21–1.25 x ULN | > 1.25 x ULN |
| PTT – increase by factor (partial thromboplastin time) | 1.0–1.2 x ULN | 1.21–1.4 x ULN | 1.41–1.5 x ULN | > 1.5 x ULN |
| Fibrinogen increase – mg/dL | 400–500 | 501–600 | > 600 | -- |
| Fibrinogen decrease – mg/dL | 150–200 | 125–149 | 100–124 | < 100 or associated with gross bleeding or disseminated intravascular coagulation (DIC) |

*The laboratory values provided in the tables serve as guidelines and are dependent upon institutional normal parameters. Institutional normal reference ranges should be provided to demonstrate that they are appropriate.
**ULN is the upper limit of the normal range.

| **Urine*** | **Mild (Grade 1)** | **Moderate (Grade 2)** | **Severe (Grade 3)** | **Life-threatening (Grade 4)** |
|---|---|---|---|---|
| Protein | Trace | 1+ | 2+ | Hospitalisation or dialysis |
| Glucose | Trace | 1+ | 2+ | Hospitalisation or dialysis |
| Blood (microscopic) – red blood cells per high power field (rbc/hpf) | 1–10 | 11–50 | > 50 and/or gross blood | Hospitalisation or packed red blood cells (PRBC) transfusion |

**The laboratory values provided in the tables serve as guidelines and are dependent upon institutional normal parameters. Institutional normal reference ranges should be provided to demonstrate that they are appropriate.
*Visit 1 includes volunteer's stay at the trial site for 2 hours after vaccination and phone call with the study doctor in 5 hours after vaccination.
