## Supplementary material for "Immunogenicity and safety of a recombinant adenovirus type-5 COVID-19 vaccine in adults: data from a randomised, double-blind, placebo-controlled, single-dose, phase 3 trial in Russia": S1 Table

**S1 Table. Systemic (general) post-vaccination reactions (Safety population).**

| **System Organ Class**  Preferred Term | **Ad5-nCoV**  **N=372**  **n (%)** | **Placebo**  **N=124**  **n (%)** | **Total**  **N=496**  **n (%)** |
| --- | --- | --- | --- |
| **At least one AE (n[%])** | **100 (26.9)** | **2 (1.6)** | **113 (2.8)** |
| **Gastrointestinal disorders** | **4 (1.1)** | **1 (0.8)** | **5 (1.0)** |
| Diarrhoea | 1 (0.3) | 1 (0.8) | 2 (0.4) |
| Nausea | 2 (0.5) | 0 | 2 (0.4) |
| Vomiting | 1 (0.3) | 0 | 1 (0.2) |
| **General disorders and administration site conditions** | **23 (6.2)** | **4 (3.2)** | **27 (5.4)** |
| Shiver | 1 (0.3) | 0 | 1 (0.2) |
| Fatigue | 20 (5.4) | 4 (3.2) | 24 (4.8) |
| Fever sensation | 2 (0.5) | 0 | 2 (0.4) |
| **Investigations** | **76 (20.4)** | **8 (6.5)** | **84 (16.9)** |
| Body temperature increase | 75 (20.2) | 8 (6.5) | 83 (16.7) |
| C-reactive protein increase | 1 (0.3) | 0 | 1 (0.2) |
| **Musculoskeletal and connective tissue disorders** | **25 (6.7)** | **0** | **25 (5.0)** |
| Arthralgia | 7 (1.9) | 0 | 7 (1.4) |
| Back pain | 1 (0.3) | 0 | 1 (0.2) |
| Myalgia | 18 (4.8) | 0 | 18 (3.6) |
| **Nervous system disorders** | **23 (6.2)** | **6 (4.8)** | **29 (5.8)** |
| Headache | 22 (5.9) | 6 (4.8) | 28 (5.6) |
| Hypoaesthesia | 1 (0.3) | 0 | 1 (0.2) |
| **Respiratory, thoracic and mediastinal disorders** | **1 (0.3)** | **0** | **1 (0.2)** |
| Panting | 1 (0.3) | 0 | 1 (0.2) |
| **Disorders of the skin and subcutaneous** | **2 (0.5)** | **0** | **2 (0.4)** |
| Hyperhidrosis | 2 (0.5) | 0 | 2 (0.4) |

Ad5-nCoV, adenovirus type-5 vectored COVID-19 vaccine; AE, adverse event; N, total number of patients; n , number of patients that experienced an AE.
