## Supplementary material for "Immunogenicity and safety of a recombinant adenovirus type-5 COVID-19 vaccine in adults: data from a randomised, double-blind, placebo-controlled, single-dose, phase 3 trial in Russia": S2 Table

**S2 Table. Systemic (general) post-vaccination reactions by severity (Safety population).**

| **Preferred Term** | **Ad5-nCoV**  **N=372**  **n (%)** | **Placebo**  **N=124**  **n (%)** | **Total**  **N=496**  **n (%)** |
| --- | --- | --- | --- |
| **Grade 1** | **78 (21.0)** | **10 (8.1)** | **88 (17.7)** |
| **Grade 2** | **17 (4.6)** | **3 (2.4)** | **20 (4.0)** |
| **Grade 3** | **5 (1.3)** | **0** | **5 (1.0)** |
| **Diarrhoea** |  |  |  |
| Grade 1 | 1 (0.3) | 0 | 1 (0.2) |
| Grade 2 | 0 | 1 (0.8) | 1 (0.2) |
| **Nausea** |  |  |  |
| Grade 1 | 2 (0.5) | 0 | 2 (0.4) |
| **Vomiting** |  |  |  |
| Grade 1 | 1 (0.3) | 0 | 1 (0.2) |
| **Shiver** |  |  |  |
| Grade 1 | 1 (0.3) | 0 | 1 (0.2) |
| **Fatigue** |  |  |  |
| Grade 1 | 18 (4.8) | 3 (2.4) | 21 (4.2) |
| Grade 2 | 2 (0.5) | 1 (0.8) | 3 (0.6) |
| **Fever sensation** |  |  |  |
| Grade 1 | 2 (0.5) | 0 | 2 (0.4) |
| **Body temperature increase** |  |  |  |
| Grade 1 | 58/64 (15.6) | 7/8 (5.6) | 65/72 (13.1) |
| Grade 2 | 13/14 (3.5) | 1 (0.8) | 14/15 (2.8) |
| Grade 3 | 4 (1.1) | 0 | 4 (0.8) |
| **C-reactive protein increase** |  |  |  |
| Grade 1 | 1 (0.3) | 0 | 1 (0.2) |
| **Arthralgia** |  |  |  |
| Grade 1 | 6 (1.6) | 0 | 6 (1.2) |
| Grade 2 | 1 (0.3) | 0 | 1 (0.2) |
| **Back pain** |  |  |  |
| Grade 1 | 1 (0.3) | 0 | 1 (0.2) |
| **Myalgia** |  |  |  |
| Grade 1 | 12 (3.2) | 0 | 12 (2.4) |
| Grade 2 | 5 (1.3) | 0 | 5 (1.0) |
| Grade 3 | 1 (0.3) | 0 | 1 (0.2) |
| **Headache** |  |  |  |
| Grade 1 | 19 (5.1) | 5 (4.0) | 24 (4.8) |
| Grade 2 | 3 (0.8) | 1 (0.8) | 4 (0.8) |
| **Hypoaesthesia** |  |  |  |
| Grade 1 | 1 (0.3) | 0 | 1 (0.2) |
| **Panting** |  |  |  |
| Grade 2 | 1 (0.3) | 0 | 1 (0.2) |
| **Hyperhidrosis** |  |  |  |
| Grade 1 | 2 (0.5) | 0 | 2 (0.4) |

Ad5-nCoV, adenovirus type-5 vectored COVID-19 vaccine; AE, adverse event; N, total number of patients; n , number of patients that experienced an AE.
