## Supplementary material for "Immunogenicity and safety of a recombinant adenovirus type-5 COVID-19 vaccine in adults: data from a randomised, double-blind, placebo-controlled, single-dose, phase 3 trial in Russia": S3 Table

**S3 Table. Summary of all serious adverse events (SAEs) that led to hospitalisation or prolonged hospitalisation.**

| **Subject** | **Treatment** | **Description** | **SOC** | **Hospital Stay** | **Outcome** | **Related to Vaccine** |
| --- | --- | --- | --- | --- | --- | --- |
| 01 | Placebo | Acute pyelonephritis | Infectious and parasitic diseases | 11 days | RECOVERED/  RESOLVED | NOT RELATED |
| 02 | Placebo | Acute haemorrhagic cystitis | Disorders of the kidneys and urinary tract | 3 days | RECOVERED/  RESOLVED | UNLIKELY |
| 03 | Placebo | COVID 19  Due to the deterioration of this condition, he sought emergency medical care and was hospitalised. | Infectious and parasitic diseases | 11 days | RECOVERED/  RESOLVED | NOT RELATED |
| 04 | Placebo | Acute obstruction calculous cholecystitis | Disorders of the hepatobiliary system | 6 days | RECOVERED/  RESOLVED | NOT RELATED |
| 05 | Placebo | COVID-19.  On Jan 09, results from a CT scan indicated bilateral polysegmental pneumonia of viral etiology, i.e., COVID-19. | Infectious and parasitic diseases | 16 days | RECOVERED/  RESOLVED | NOT RELATED |
| 06 | Ad5-nCoV | Inflammatory neoplasm in the lower lobe of the left lung | Benign, malignant and unspecified neoplasms | 147 days | RECOVERED/  RESOLVED | NOT RELATED |

Ad5-nCoV, adenovirus type-5 vectored COVID-19 vaccine; COVID-19, coronavirus 2019; CT, computed tomography; SOC, system organ class.
