## Supplementary material for "Immunogenicity and safety of a recombinant adenovirus type-5 COVID-19 vaccine in adults: data from a randomised, double-blind, placebo-controlled, single-dose, phase 3 trial in Russia": S4 Table

**S4 Table. Local post-vaccination reactions (Safety population).**

| **System Organ Class**  Preferred Term | **Ad5-nCoV**  **N=372**  **n (%)** | **Placebo**  **N=124**  **n (%)** | **Total**  **N=496**  **n (%)** |
| --- | --- | --- | --- |
| **At least one AE (n[%])** | **106 (28.5)** | **2 (1.6)** | **108 (21.8)** |
| **General disorders and administration site conditions** | **105 (28.2)** | **2 (1.6)** | **107 (21.6)** |
| Erythema | 55 (14.8) | 0 | 55 (11.1) |
| Haematoma | 1 (0.3) | 0 | 1 (0.2) |
| Induration | 14 (3.8) | 1 (0.8) | 15 (3.0) |
| Pain | 63 (16.9) | 1 (0.8) | 64 (12.9) |
| Itching | 7 (1.9) | 0 | 7 (1.4) |
| Swelling | 18 (4.8) | 2 (1.6) | 20 (4.0) |
| **Musculoskeletal and connective tissue disorders** | **2 (0.5)** | **0** | **2 (0.4)** |
| Muscle weakness | 1 (0.3) | 0 | 1 (0.2) |
| Arm pain | 1 (0.3) | 0 | 1 (0.2) |
| **Nervous system disorders** | **1 (0.3)** | **0** | **1 (0.2)** |
| Hyperhidrosis | 1 (0.3) | 0 | 1 (0.2) |

Ad5-nCoV, adenovirus type-5 vectored COVID-19 vaccine; AE, adverse event; N, total number of patients; n , number of patients that experienced an AE.
