## Supplementary material for "Immunogenicity and safety of a recombinant adenovirus type-5 COVID-19 vaccine in adults: data from a randomised, double-blind, placebo-controlled, single-dose, phase 3 trial in Russia": S5 Table

**S5 Table. Local post-vaccination reactions by severity (Safety population).**

| **Preferred Term** | **Ad5-nCoV**  **N=372**  **n (%)** | **Placebo**  **N=124**  **n (%)** | **Total**  **N=496**  **n (%)** |
| --- | --- | --- | --- |
| **Grade 1** | **91 (24.5)** | **2 (1.6)** | **93 (18.8)** |
| **Grade 2** | **15 (4.0)** | **0** | **15 (3.0)** |
| **Erythema** |  |  |  |
| Grade 1 | 47 (12.6) | 0 | 47 (9.5) |
| Grade 2 | 8 (2.2) | 0 | 8 (1.6) |
| **Haematoma** |  |  |  |
| Grade 1 | 1 (0.3) | 0 | 1 (0.2) |
| **Induration** |  |  |  |
| Grade 1 | 13 (3.5) | 1 (0.8) | 14 (2.8) |
| Grade 2 | 1 (0.3) | 0 | 1 (0.2) |
| **Pain** |  |  |  |
| Grade 1 | 57 (15.3) | 1 (0.8) | 58 (11.7) |
| Grade 2 | 6 (1.6) | 0 | 6 (1.2) |
| **Itching** |  |  |  |
| Grade 1 | 7 (1.9) | 0 | 7 (1.4) |
| **Swelling** |  |  |  |
| Grade 1 | 13 (3.5) | 2 (1.6) | 15 (3.0) |
| Grade 2 | 5 (1.3) | 0 | 5 (1.0) |
| **Muscle weakness** |  |  |  |
| Grade 1 | 1 (0.3) | 0 | 1 (0.2) |
| **Arm pain** |  |  |  |
| Grade 1 | 1 (0.3) | 0 | 1 (0.2) |
| **Hypoaesthesia** |  |  |  |
| Grade 1 | 1 (0.3) | 0 | 1 (0.2) |

AE, adverse event; N, total number of patients; n , number of patients that experienced an AE.
