## Supplementary material for "Immunogenicity and safety of a recombinant adenovirus type-5 COVID-19 vaccine in adults: data from a randomised, double-blind, placebo-controlled, single-dose, phase 3 trial in Russia": S6 Table

**S6 Table. Other adverse events unrelated to vaccination reactions (Safety population).**

| **Description** | **Ad5-nCoV**  **N=372**  **n (%)** | **Placebo**  **N=124**  **n (%)** | **Total**  **N=496**  **n (%)** |
| --- | --- | --- | --- |
| **Out-of-range laboratory measurements (n[%])** | **128 (34.4)** | **21 (16.9)** | **149 (30.0)** |
| Elevated C-reactive protein | 67 (18.0) | 3 (2.4) | 70 (14.1) |
| Increase in the number of monocytes | 14 (4.0) | 0 (0.0) | 14 (3.0) |
| Increase in aspartate aminotransferase | 9 (2.4) | 2 (1.6) | 11 (2.2) |
| Increase in blood fibrinogen | 4 (1.1) | 0 (0.0) | 4 (0.8) |
| Decrease in blood fibrinogen | 2 (0.5) | 1 (0.8) | 3 (0.6) |
| Reduction in the number of neutrophils | 24 (6.5) | 3 (2.4) | 27 (5.4) |

Ad5-nCoV, adenovirus type-5 vectored COVID-19 vaccine; N, total number of patients;
n , number of patients that experienced an AE.

Note: Other laboratory parameters changed in rare cases. Information available upon request.
